# Quantifying SARS-CoV-2 spread in Switzerland based on genomic sequencing data

**DOI:** 10.1101/2020.10.14.20212621

**Authors:** Sarah Nadeau, Christiane Beckmann, Ivan Topolsky, Timothy Vaughan, Emma Hodcroft, Tobias Schär, Ina Nissen, Natascha Santacroce, Elodie Burcklen, Pedro Ferreira, Kim Philipp Jablonski, Susana Posada-Céspedes, Vincenzo Capece, Sophie Seidel, Noemi Santamaria de Souza, Julia M. Martinez-Gomez, Phil Cheng, Philipp P. Bosshard, Mitchell P. Levesque, Verena Kufner, Stefan Schmutz, Maryam Zaheri, Michael Huber, Alexandra Trkola, Samuel Cordey, Florian Laubscher, Ana Rita Gonçalves, Karoline Leuzinger, Madlen Stange, Alfredo Mari, Tim Roloff, Helena Seth-Smith, Hans H. Hirsch, Adrian Egli, Maurice Redondo, Olivier Kobel, Christoph Noppen, Niko Beerenwinkel, Richard A. Neher, Christian Beisel, Tanja Stadler

## Abstract

Pathogen genomes provide insights into their evolution and epidemic spread. We sequenced 1,439 SARS-CoV-2 genomes from Switzerland, representing 3-7% of all confirmed cases per week. Using these data, we demonstrate that no one lineage became dominant, pointing against evolution towards general lower virulence. On an epidemiological level, we report no evidence of cryptic transmission before the first confirmed case. We find many early viral introductions from Germany, France, and Italy and many recent introductions from Germany and France. Over the summer, we quantify the number of non-traceable infections stemming from introductions, quantify the effective reproductive number, and estimate the degree of undersampling. Our framework can be applied to quantify evolution and epidemiology in other locations or for other pathogens based on genomic data.

**One Sentence Summary:** We quantify SARS-CoV-2 spread in Switzerland based on genome sequences from our nation-wide sequencing effort.

## Main Text

SARS-CoV-2 originated in late 2019 in Wuhan, China and has been spreading rapidly around the globe in 2020. After the initial outbreak in China, Europe soon became the epicenter of the pandemic. As a counter measure, European countries implemented a variety of non-pharmaceutical interventions (NPIs), ranging from border closures to social distancing regulations (*1*). Case numbers declined to a low in early summer but are now increasing again across Europe (*2*). Switzerland is centrally located in Europe and is well-connected with other European countries. Switzerland also implemented some of the same NPIs as other European countries. Thus, we hypothesize that viral introduction and epidemiological dynamics within Switzerland are exemplary for a European country.

We characterize the SARS-CoV-2 epidemic in Switzerland from the first confirmed case on Feb. 24^th^ until Aug. 31^st^ based on SARS-CoV-2 genome sequences. We define an “early epidemic” from Feb. 24^th^ until the low point in daily cases around Jun. 1^st^ and a “summer epidemic” from Jun. 1^st^ until the end of our sampling period on Aug. 31^st^. To characterize the epidemic over the complete period, we generate 1,439 sequences from our nation-wide sequencing project, representing 3-7% of all laboratory-confirmed cases per week across Switzerland. We couple these data with 675 additional sequences from the early epidemic collected by labs in Basel (*3*), Geneva and Zurich to represent in total 5% of all laboratory-confirmed cases in Switzerland until Aug. 31^st^. We combine this Swiss dataset with additional sequences from across the globe to address five main goals:

i. to assess the change in viral genomic diversity within Switzerland,
ii. to estimate the timing of introduction events and longevity of transmission chains within Switzerland,
iii. to estimate the geographic origin of introductions and geographic spread of transmission chains within Switzerland,
iv. to quantify the relative importance of introductions versus local transmission through time, and
v. to quantify the effective reproductive number and case underreporting in Switzerland.

To meet these goals, we perform a phylogenetic analysis of the sequence dataset as detailed in the Materials and Methods. Briefly, we pair Swiss SARS-CoV-2 sequences with sequences from abroad that are either genetically similar to Swiss sequences (“similarity dataset”) or that stem from countries with lots of travel connections to Switzerland (“context dataset”). We then construct a dated maximum-likelihood phylogenetic tree using all three datasets that approximates the transmission history of the considered cases.

We define Swiss transmission chains based on this phylogenetic tree. Essentially, Swiss sequences that are more similar to each other than to sequences from abroad are proposed to belong to the same transmission chain. We apply two different extreme criteria to define transmission chains, namely, a “maximum introductions, minimum local transmission” assumption and a “minimum introductions, maximum local transmission” assumption (see Materials and Methods for details). In this way, we refrain from arbitrarily resolving phylogenetic uncertainty. Instead, we provide upper- and lower-bound estimates for epidemiological parameters of the Swiss epidemic based on these two extreme assumptions. All Swiss sequences that are not linked to a transmission chain are coined singletons. Importantly, each Swiss transmission chain and each singleton is assumed to result from an independent introduction event into Switzerland.

After identifying Swiss introductions, we estimate their geographic origins using a parsimony-based approach. For this, we consider only sequences in the context dataset. Since the context dataset is constructed based on travel connections with Switzerland and not foreign sequencing effort, we aim to reconstruct the true source location of introductions rather than reflect biased sampling.

We extract answers for goals (i-iv) directly from the phylogenetic tree and the corresponding introductions and transmission chains. Finally, we perform additional Bayesian phylodynamic analyses of the identified Swiss transmission chains and singletons to achieve goal (v). Goals (i, iii, iv, v) require a sequence set which is representative for the Swiss epidemic and thus we use only Swiss sequences from our nation-wide SARS-CoV-2 sequencing project. For goal (ii) we use all available Swiss sequences.

## Results

### (i) Change in viral genomic diversity

First, we assess how the SARS-CoV-2 population changed through time within Switzerland. By sequencing 3-7% of confirmed cases each week (Fig. S1) from across the country (Fig. S2), we argue that we obtain a representative picture of SARS-CoV-2 genetic diversity in Switzerland through time. Here we use the Nextstrain nomenclature standard to describe genetic diversity.

Fig. 1a shows that in Switzerland 20A strains dominated until early summer, then 20B increased in frequency throughout the summer, and finally 20A again became dominant in late August. Since 20B strains increased in frequency while case numbers in Switzerland dropped, and later numbers of new hospitalizations remained low despite rising case numbers (*2*), one may hypothesize that a single 20B strain evolved lower virulence and/or lower pathogenicity before becoming dominant during the summer epidemic. However, on the Swiss phylogenetic tree many lineages cross from the early epidemic into the summer epidemic. Taking June 1 to be the start of the summer epidemic, 114 lineages give rise to 721 sequences from the summer epidemic, with the 8 most fecund lineages producing 60% of sequences from the summer epidemic (Fig. 1b). Similar results are obtained using different date cutoffs between the early and summer epidemic (Fig. S3).

**Fig. 1.**
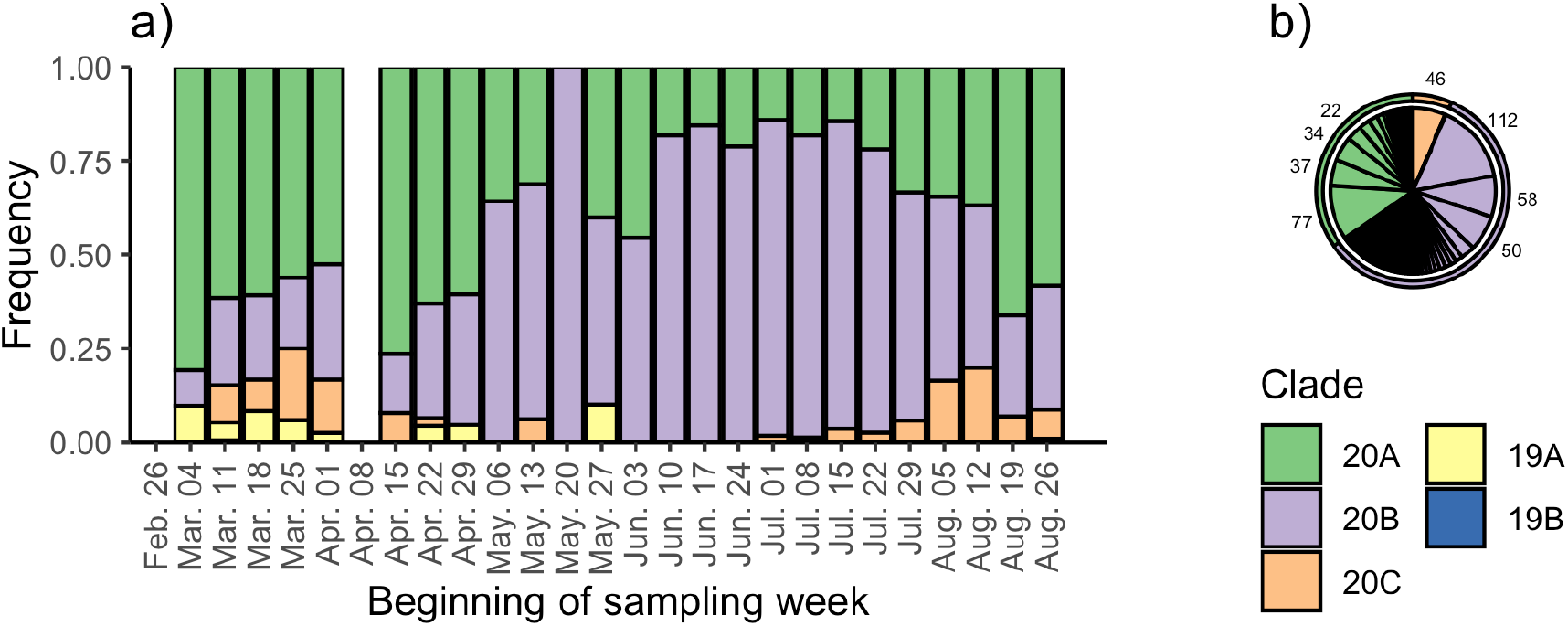
Genomic diversity of the Swiss epidemic over time based on sequences from our nation-wide sequencing project. **(a)** shows the frequency of different clades over time. **(b)** shows the number of samples from the summer epidemic generated by different lineages from the early epidemic. Each slice represents a unique lineage present on Jun. 1 2020, where the width of the slice is proportional to the number of sampled descendants after Jun. 1. In total, 114 lineages present on Jun. 1 gave rise to 721 samples from the summer epidemic. Combined, the 8 largest lineages (sample numbers shown) gave rise to 60% of the samples from the summer epidemic.

In summary, we show that although the dominant clade in the Swiss epidemic has changed several times, it is implausible that any single strain evolved lower virulence (or any other remarkable phenotype) and then became dominant.

### (ii) Timing of introduction events and longevity of transmission chains

Next, we estimated the timing of viral introductions into Switzerland by focusing on the identified introductions. We note that phylogenetic results need to be interpreted carefully due to incomplete sampling and phylogenetic uncertainty. In fact, in Fig. S4 we show that our sampling is not saturated, meaning that if we were to sequence more genomes, we would find more introductions. As mentioned, we account for phylogenetic uncertainty by presenting results robust to two different assumptions about the degree of local transmission vs. introductions.

Our insights on the introduction times of SARS-CoV-2 lineages into Switzerland rely on the fact that a lineage could have been introduced into Switzerland anytime between the date of the most recent common ancestor (MRCA) of a Swiss transmission chain and the date the MRCA attaches to the rest of the tree. For a singleton, the introduction could have occurred anytime between the attachment point and the sample date. Fig. 2 shows that for the early epidemic we cannot pinpoint transmission chain introduction dates very precisely. Fig. S5 shows that the same holds for singletons. As others have noted, there are not enough informative mutations to resolve the ordering of many transmission events (*4*) or to confidently estimate MRCA dates (*5*). However, with more samples, the MRCAs may become older and the attachment times may become younger, reducing this uncertainty.

**Fig. 2.**
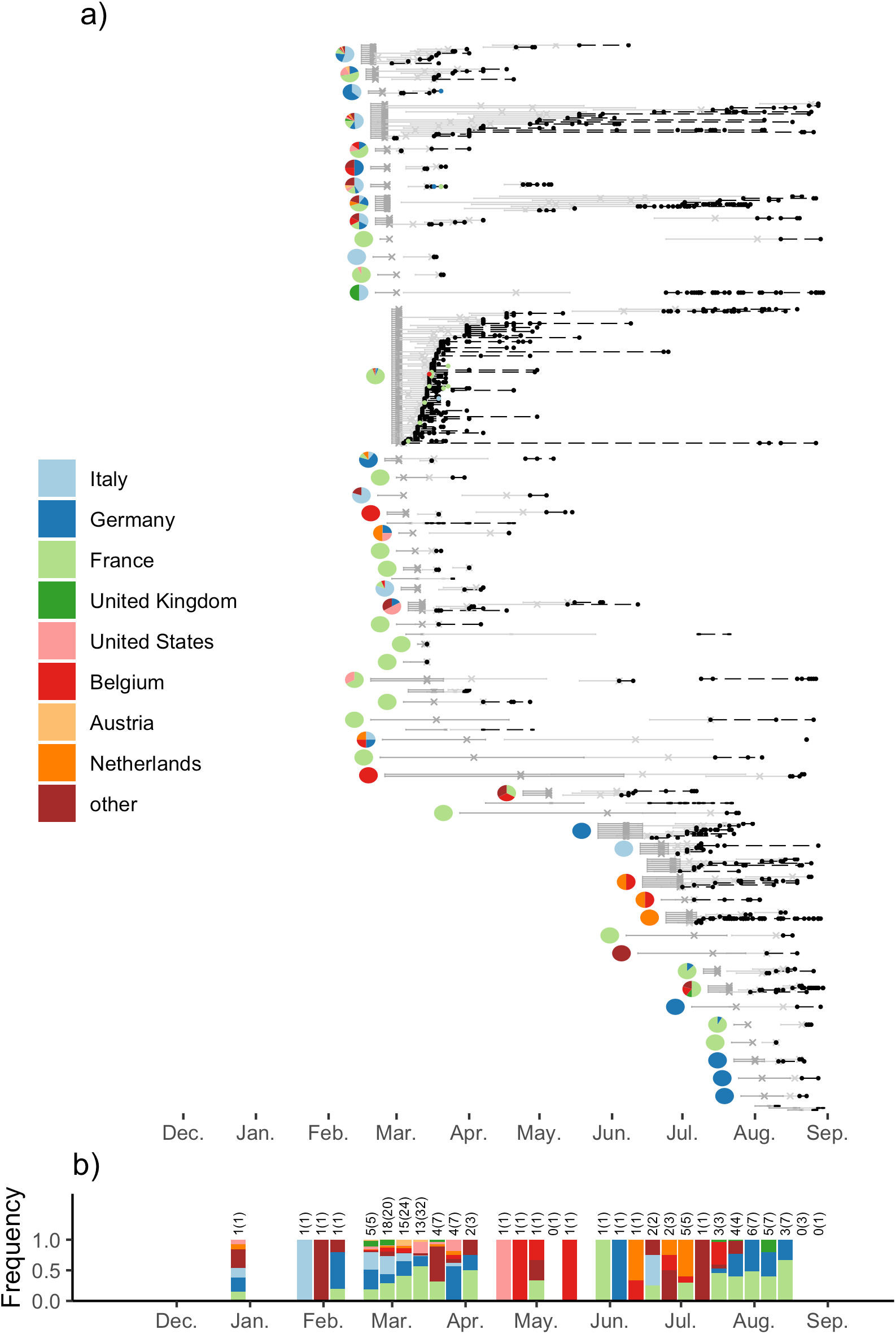
**(a)** Swiss transmission chains based on all publicly available Swiss sequences. Dashed lines connect samples from the same transmission chain. Samples with known travel information are colored accordingly. The dark grey intervals are estimates for the attachment time of each transmission chain to the rest of the tree and the light grey intervals are estimates for each transmission chain’s MRCA, where crosses are the least-square estimates for the respective date. The pie charts show estimates for the location of each attachment point. We do not infer geographic origins for transmission chains that cluster with only sequences from the “similarity” dataset. Fig. S4 shows the corresponding plot for singletons. **(b)** The estimated contribution of each country towards introductions (transmission chains and singletons) into Switzerland over time. The bars summarize how frequent each location is across the least-square attachment times (dark grey crosses) dated during each week. The numbers above the bars give the number of attachment points with location information falling in that week; values in parentheses are the total number of attachment points. We note that the bar in late December corresponds to a polytomy with Swiss singleton descendants (Fig. S5). The start of local spread within Switzerland is very likely to have been much later (Fig. S6).

Given that transmission chain MRCAs are the earliest evidence of local transmission, we do not find evidence for cryptic spread in Switzerland prior to detection of the first case on Feb. 24^th^. The oldest estimated MRCA is Feb. 22^nd^ (90^th^ percentile confidence interval Feb. 17^th^ – Mar. 7^th^). In fact, the majority of lineages which generated Swiss cases during the early epidemic were still circulating outside of Switzerland in the weeks Feb. 26^th^ – Mar. 10^th^ (Fig. S6a). In comparison, only a few were still circulating outside of Switzerland after the lockdown and border closures were implemented in Switzerland on Mar. 17^th^ (Fig. S6a).

About a third of all sampled Swiss transmission chains and more than half of the transmission chains of the early epidemic (here defined as starting spreading prior to June 1) began spreading locally around Mar. 4 – 24^th^ (Fig. S6b). Three of these transmission chains persisted from March until August and due to incomplete sampling, we may underestimate the longevity of some other transmission chains. Finally, transmission chains in the summer epidemic largely started spreading locally after Jun. 15^th^, the date Switzerland lifted its partial border closure.

These results highlight that there is no evidence of cryptic transmission prior to the first confirmed case in Switzerland, that many introduction events occurred just before Swiss border closures, and that new introduction events occurred just after borders were re-opened.

### (iii) Geographic origin and geographic spread of transmission chains

We asked whether the source of Swiss introductions changed over the course of the epidemic thus far. To do this, we estimated the geographic location of attachment points of Swiss introductions. We used only our context dataset of foreign sequences to generate these estimates, which should reduce the effect of uneven foreign sampling efforts. However, we note that we may underestimate the contributions of Italy, France, and Germany during some summer months as we lack sequences from some of these countries (Fig. S7).

In Fig. 2 we show that while initially the origin of introduced strains was from our neighboring countries France, Italy, and Germany, the source location shifted more towards Belgium and the Netherlands in May – July and then, despite under-sampling, to Germany and France more recently. These results were roughly consistent across three different random samplings of context sequences (Fig. S8 – S10). Some of our source location estimates can be confirmed with independent information on case travel history. Where this information is available, our estimates for transmission chain origins largely agree (Fig. 2a).

To investigate epidemic spread between Swiss cantons, we tallied how often samples from the same transmission chain were found in different Swiss cantons (Fig. S11). We observe significantly more mixing than expected by chance between the neighboring cantons of Basel-Stadt and Basel-Land, with 16 transmission chains containing samples from both cantons. None of the cantons mixed less than expected based on the available data.

We conclude that in general, SARS-CoV-2 introduction dynamics into Switzerland shifted from neighboring countries early in the epidemic towards more non-neighboring countries as border restrictions eased and then finally back to neighboring countries near the end of the summer holidays. Within Switzerland, there is evidence that two neighboring cantons share a single intermixed epidemic.

### (iv) Relative importance of introductions versus local transmission

Next, we assessed the relative importance of introductions compared to local transmissions over time. Importantly, the chance of sampling a local transmission compared to an introduction increases with increased sampling. Thus, local transmission may be even more significant than we report here.

Fig. 3a shows that we sample many new introductions in March (up to 28-161 in the most extreme week) compared to almost none from mid-May to mid-June. These trends coincide with closed borders in Switzerland from Mar. 17^th^ – Jun. 15^th^. We then had a stable period with 1-28 introductions per week until the end of July. In August, towards the end of the summer holidays, we see a steady increase in introductions. The probability for a new case to arise from a local transmission (as opposed to an introduction) rose throughout the epidemic to above 80% in July. In August, this probability dropped to around 40%-90% as introductions again increased (Fig. 3a).

**Fig. 3.**
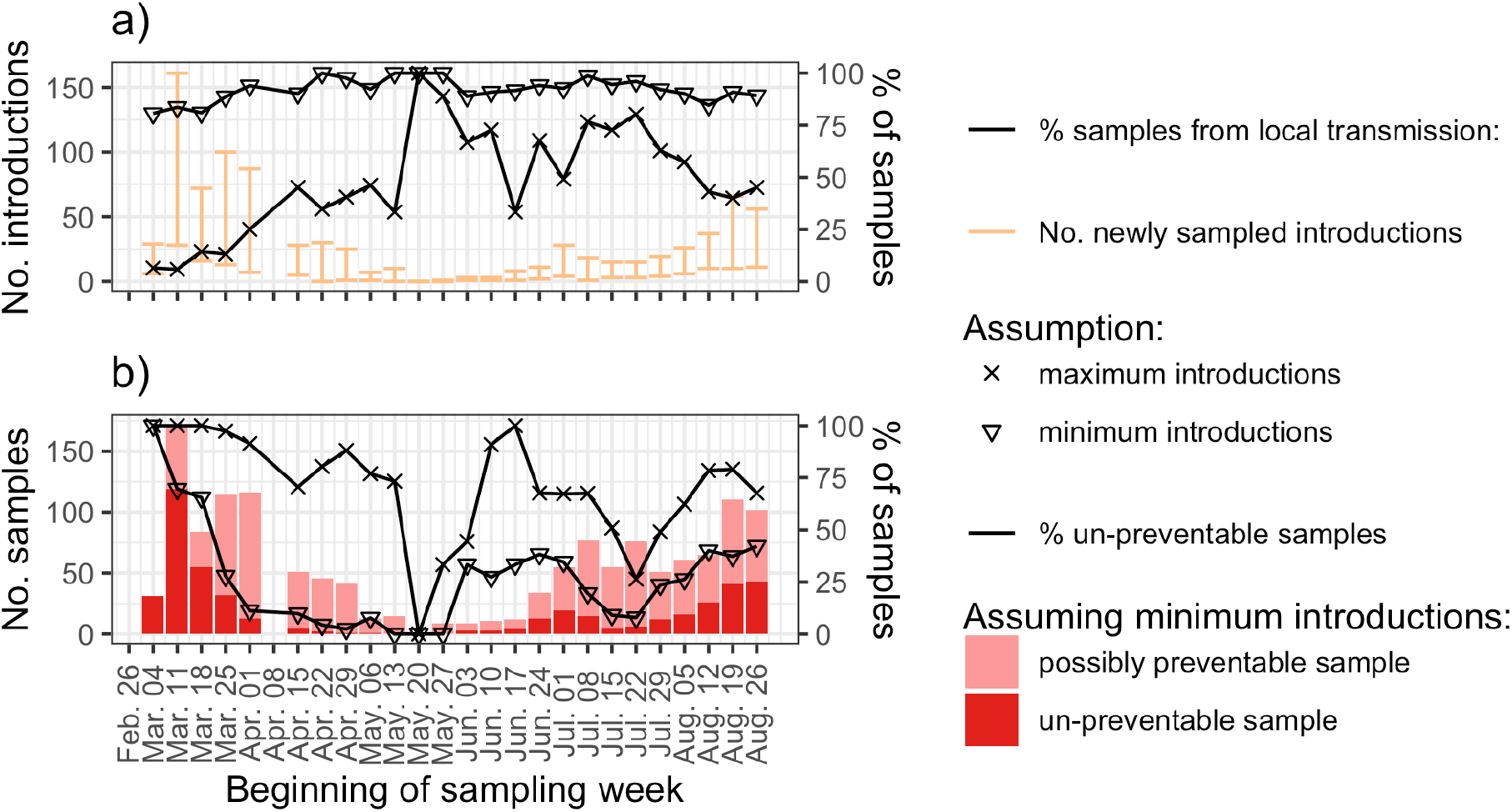
Importance of local transmission versus introductions through time based on sequences from our nation-wide sequencing project. Here we label the first sample from each introduction the index sample. We then count the number of index samples (i.e. newly sampled introductions) and non-index samples (i.e., samples from local transmission) each week. **(a)** shows the number of newly sampled introductions (transmission chains and singletons) each week in orange and the percentage of samples from local transmission each week in black. **(b)** shows the lower bound number (dark red) and percentage (black) of samples that are un-avoidable even given a perfect test, trace, quarantine, and isolate strategy (see also Supplementary Text). Importantly, un-preventable cases result from transmission chains caused by introductions prior to these introduced transmission chains being traceable.

To quantify the importance of these trends, we estimate the number of samples that are un-preventable vs. preventable via perfect contact tracing. To do this, we rely on the insight that cases generated between an introduction index case’s infection date and case confirmation (we assume a 10-day period (*6*)) cannot be prevented because contact tracers are not yet aware of the transmission chain. We estimate that there were >100 such un-preventable secondary cases in the week prior to the lockdown. This number decreased to <10 per week in late spring, and more recently increased to around 50 per week. On the flipside, Fig. 3b shows that over the summer, 25% or more of the new infections each week could have been prevented under the optimistic assumption that contact tracing could have prevented all cases confirmed more than 10 days after the index case.

These results highlight the fact that transmission by introductions is harder to contain than local transmission as we cannot identify them quickly through contact tracing. As local transmission increases, contact tracing becomes a very effective strategy to prevent onward transmission.

### (v) Effective reproductive number

Finally, we quantify the effective reproductive number (R_e_) and under-sampling in Switzerland by fitting a phylodynamic model to the identified Swiss transmission chains and singletons. We only present estimates after May 1^st^ because prior to this time, low viral diversity contributes to high uncertainty in the size and number of transmission chains (i.e. Fig. 3a shows that the estimated number of introductions varies widely depending on the assumptions used).

Beginning May 1^st^, we estimate that R_e_ was significantly below 1 (Fig. 4a). Coincident with the opening of shops and restaurants on May 11^th^, R_e_ increased and our estimates include the threshold of 1 throughout most of the summer. The estimates of R_e_ based on confirmed case data are contained within our estimated uncertainty intervals for most of the time.

**Figure 4.**
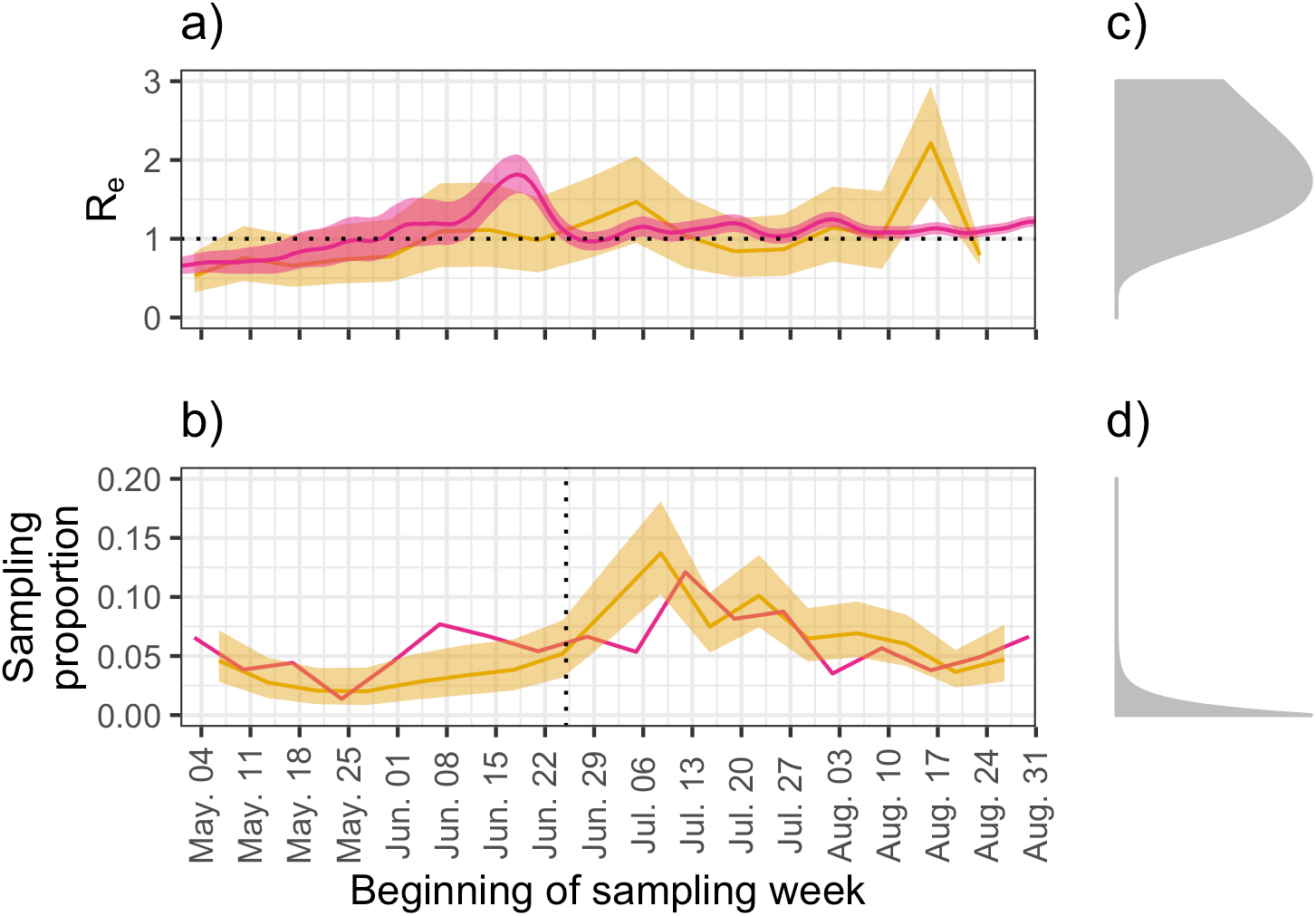
**(a)** Estimates of the effective reproductive number (R_e_) and **(b)** the sampling proportion in Switzerland through time based on genome sequences from our nation-wide sequencing project (yellow) compared with estimates from line-line data (pink, (*2,31*)). **(c, d)** The prior distributions applied to R_e_ and the sampling proportion. The week-to-week smoothing prior applied to both parameters (see Supplementary Text) is not shown.

Our phylodynamic analysis co-infers the sampling proportion, which is the proportion of total cases included in the analysis, over time. Fig. 4b shows that the inferred sampling proportion closely matches the fraction of total confirmed cases included in the analysis. Therefore, we do not find evidence of significant undersampling during the summer epidemic in Switzerland.

Overall, genome-based estimates of R_e_ match independent estimates based on confirmed case data. Pending confirmation with seroprevalence information, genome-based estimates of the sampling proportion suggest most infections over the summer were successfully identified.

## Discussion

By analyzing a time-homogenous dataset of Swiss SARS-CoV-2 genome sequences, we generated evolutionary and epidemiological insights on SARS-CoV-2 spread for a centrally located and well-connected European country. The insights that we obtained regarding our goals (i-v) can be directly relevant for public health policies.

First, knowing that no one lineage evolved lower virulence and then became dominant means that the current low numbers of hospitalizations and deaths relative to case numbers in Switzerland (*2*) cannot be explained by viral evolution. In fact, the shift in case age distribution towards younger people (*7*) may well explain the low numbers.

On an epidemiological level, we observe no cryptic spread prior to the first confirmed case in Switzerland, suggesting that the Swiss early surveillance was well-functioning. As recent studies have highlighted (*8, 9*), early identification is crucial in order to buy time for preparing a pandemic response plan. With currently increasing case numbers across Europe, the usefulness of maintaining and expanding quarantine rules is becoming a widely discussed topic. In the summer epidemic in Switzerland, we identify first Belgium and the Netherlands as important sources for introductions and then more recently, France and Germany. Of these countries, only Belgium was put on the Swiss quarantine list (from Aug. 20) shortly before the end of our study period on Aug. 31. Since then, quarantine has been made mandatory for travelers returning from Belgium, the Netherlands, and many regions of France and Germany. Given our baseline estimates for introductions in the absence of mandatory quarantine, it will be interesting to assess the impact of quarantine rules going forward.

Finally, we estimate that, in comparison to the early epidemic where a 5-10 time underreporting was observed based on seroprevalence studies (*10*), confirmed case counts over the summer closely match genome-based estimates of the total number of infected individuals. This suggests that the SARS-CoV-2 epidemic in Switzerland can in fact be well-monitored. However, this estimate should be assessed for plausibility through additional seroprevalence studies.

We will continue to update our genome-based results on SARS-CoV-2 evolution and epidemiology as the epidemic continues and plan to publish them on the Swiss National COVID-19 Scientific Task Force website and a Swiss Nextstrain site (*11*). Going forward, we call for linking epidemiological metadata, contact tracing information, clinical data, and genomic data in order to extract the maximum amount of information about epidemiological dynamics. Such information is crucial in order to understand where infections happen and thus to design specific and efficient public health interventions.

## Data Availability

All sequence data used is available on https://www.gisaid.org/.

## Notes

### Competing Interest Statement

The authors have declared no competing interest.

### Funding Statement

SN and TS are supported by the Swiss National Science Foundation (grant number 31CA30_196267).

