## Supplemental Information for "Quantifying SARS-CoV-2 spread in Switzerland based on genomic sequencing data"

**This PDF file includes:**

Materials and Methods
Supplementary Text
Figs. S1 to S13
Tables S1 to S3

### Materials and Methods

Genome sequencing

Most of the Swiss SARS-CoV-2 genome samples analyzed were provided by Viollier AG, a Swiss medical diagnostics company. RNA was extracted from patient naso- or oral-pharangeal swabs using either the Abbott m2000sp or Seegene STARMag 96x4 Universal Cartridge RNA extraction kit. One aliquot of RNA extract was used for a qPCR test by Viollier and the remaining material from PCR-positive samples was transferred to the Genomics Facility Basel for whole-genome sequencing. Amplicon sequencing was performed using the ARCTIC v3 primer scheme, which yields tiled amplicons of approximately 400bp (*12*). Libraries were sequenced on an Illumina MiSeq machine, resulting in 2 x 251 base reads. We used V-pipe (*13*) for raw read quality control, mapping to the reference genome MN908947.3, and consensus base calling. Positions with <5x coverage were not called and positions with >5% minor base composition were assigned the appropriate ambiguity code. We rejected samples where <90% of quality-controlled reads did not map to the reference or with < 20,000 bases called. The consensus sequences we generated have been made publicly available on both GISAID (Accession IDs are given in Table S1) and ENA (samples are registered under study PRJEB38472).

Genome quality control

We supplemented our Swiss data with other Swiss sequences and foreign sequences available via the "nextfasta" downloaded resource on GISAID (accessed 10. October 2020). We removed non-human samples, duplicates, samples lacking a complete date specification, and samples with sequencing issues flagged by the Nextstrain team (*14*). We aligned the sequences to the reference genome MN908947.3 using MAFFT. We retained all Swiss sequences ≥20 kbases long and foreign sequences ≥27 kbases long. We then masked sites with high homoplasy or other known problems (*15*, *16*). Finally, we used the Nextstrain diagnostic script to additionally exclude sequences with unexpectedly high divergence from the reference, clusters of SNPs in close proximity, or >3000 missing bases (criterion relaxed to >10000 missing bases for Swiss sequences). Sequence counts after each quality-control step are given in Table S2.

Subsampling

For analysis, we considered all quality control-passed sequences from Switzerland ("Swiss dataset") along with the most genetically similar sequences from abroad ("similarity dataset") and a subset of context sequences from countries likely to have exported cases to Switzerland ("context dataset").

The similarity dataset (selected using the Nextstrain priority protocol) was used to identify Swiss transmission chains. Our rationale is that diversity sampled primarily within Switzerland and not abroad is likely the result of within-Switzerland transmission.

The context dataset was additionally used for parsimony-based inference of the origin locations of Swiss introductions. For this purpose, we wanted to include a random sample of sequences from the pool of cases abroad who might seed Swiss transmission chains.

To determine how many sequences to take from each country and each month for the context dataset, we estimated the expected number of infected individuals arriving in Switzerland over time (Fig. S12). We calculated this to be the number of arrivals from each country in a month multiplied by the average proportion of the source population infectious on any given day in that month. We estimated the number of arrivals to be the number of hotel arrivals from a country plus the number of cross-border commuter permits held by its residents (*17*, *18*). We estimated the number of infectious individuals each day by assuming individuals were infectious for 10 days prior to case confirmation. Confirmed case and national population data were taken from the ECDC (*19*). Our estimates for the number of infected individuals arriving in Switzerland each month are intended only to be proportional to the expected number of arrivals, subject to several simplifying assumptions. Namely, the propensity of an infected individual to travel does not vary by country, travel between Switzerland and other countries is reciprocal (since we do not have data on the number of Swiss travelling abroad who then re-enter the country), and finally that the number of confirmed cases in each country is a constant proportion of the true number of cases over time.

Once we identified the number of context sequences to take from each country in each month we randomly selected sequences publicly available on GISAID. For months without enough sequences, we took extra sequences from the following month if possible and the preceding month if necessary. The numbers of sequences included from each country in each month is shown in Fig. S4. We applied this sub-sampling protocol three times with different random seeds. As an additional sensitivity check, we also padded the estimated imports per country and month by 1 before determining the number of context sequences to take. Based on the padded numbers, we randomly chose another set of context sequences. In all, we analyzed three replicate context datasets and one context dataset based on padded imports. In the main text, we discuss results based on only one of the replicate datasets; results from the other datasets are qualitatively similar (Fig. S8 – S10).

The final dataset includes 2,114 Swiss sequences, 1,000 genetically similar sequences, and 1,099 context sequences (approximately 4,190 sequences in all depending on the context dataset, since some context sequences were also selected as genetically similar sequences).

Clade assignment

We assigned the genome sequences from our nation-wide genome sequencing project to Nextstrain clades using the Nextclade tool (*20*). We classified each lineage in the Swiss tree that crossed our date threshold from the early to the summer epidemic as belonging to the same clade as the majority of the tips descending from it. These assignments were unambiguous with the exception of 3-6 lineages, depending on the date threshold used.

Tree building

We built a maximum-likelihood tree using IQ-TREE (*21*) and then estimated branch lengths for the maximum-likelihood topology using least-squares dating (*22*). For the initial maximum-likelihood tree inference, we used an HKY substitution model with empirical base frequencies and 4 gamma rate categories. To root the tree, we specified an outgroup defined by the most recent common ancestor (MRCA) of two early sequences from Nextstrain clades 19A and 19B (EPI_ISL_402125 and EPI_ISL_406798). This placed the root of the tree roughly between the 19A/B clades and ingroup clade 20A (Nextstrain clade notation), or the L, S, O, V clades and ingroup clades G, GR, and GH (GISAID clade notation). For least-squares dating, we assumed a strict molecular clock with a minimum mutation rate of 0.0008 substitutions/site/year. We additionally assumed the root date to be between 15. Nov. and 24. Dec. 2019 (roughly in line with estimates provided by (*23*)) and set the minimum branch length to zero. Sequences that violated the strict clock assumption (z-score threshold > 3) were removed and near-zero branches ($\leq 1.7 \times 10^{-5}$ substitutions/site/year) were collapsed into polytomies, reflecting the fact that the sequence data is not sufficient to resolve the ordering of these transmission events. Given the root date constraints, the mutation rate conformed to the lower bound of 0.0008 with extremely narrow confidence intervals. Confidence intervals for node dates were generated in LSD by re-sampling branch lengths 100 times under a lognormal relaxed clock model with standard deviation 0.4.

Identifying SARS-CoV-2 introductions into Switzerland

In the tree based on sequences from the Swiss, similarity, and context datasets we define Swiss *transmission chains* as maximal sets of at least 2 Swiss sequences satisfying the following criteria: the Swiss sequences are part of a clade in the tree and the subtree spanned by these Swiss sequences is monophyletic upon removing (a) up to 3 export events where (b) only one export event may occur along each internal branch. Exports are defined to be clades containing non-Swiss sequences. We chose a conservative value for criterion (b) while still allowing some export events and note that the number of inferred transmission chains is robust to different values for criterion (a) given criterion (b) (Fig. S13).

We repeated our analysis interpreting polytomies in two ways. Once, we split Swiss clades descending from polytomies unless the polytomy only had a single non-Swiss descendent (see criterion (b)). Alternatively, we aggregated all Swiss clades descending from polytomies into a single transmission chain. These procedures represent two plausible extremes, the first being maximum introductions and minimum local transmission and the second being minimum introductions and maximum local transmission.

We refer to any Swiss sequence not falling into a Swiss transmission chain as a Swiss *singleton*. We assume each singleton and each transmission chain represent an independent introduction of SARS-CoV-2 into Switzerland; together these are called Swiss *introductions*.

We inferred the source location of each introduction with a parsimony-based approach. For this, we used only tips in the context dataset and ignored tips in the similarity dataset. Otherwise, over-sequenced locations would show up too often as sources (*24*). Our procedure is as follows: we begin by labelling the MRCAs of Swiss transmission chains and all subtending nodes in the tree (excepting exported clades) as Swiss. Switzerland was excluded as a possible location for all other nodes. Given these constraints, we calculate the parsimony score for each possible location at each remaining node. Among the possible locations, we also include a "dummy" location to record the maximum parsimony score. So, in a first step we calculate parsimony scores up the tree. In a second step, we covert the parsimony scores at each node to location weights by taking the difference to the "dummy" score. Thus, locations with no support in the subtending tree are given a weight 0 and supported locations are weighted by the number of location changes they prevent. Importantly, these weights are determined through the parsimony score of the subtree descending the considered node and not of the full tree. Finally, we normalized the scores to 1 for each node. In summary, locations that would require more state changes in the subtending tree are down-weighted. For the summary figure Fig. 2b, we conservatively count each polytomy with Swiss descendants as a single introduction.

Phylodynamic analysis

Finally, after grouping sequences into introductions using our clustering procedure, we jointly infer the effective reproductive number and the sampling proportion from all clusters as in (*25*). We perform inference using the BDSKY model (*26*) in BEAST2 (*27*), which assumes a birth-death population dynamical model. Each cluster was assumed to result from an independent birth-death process having its own start time, but sharing all other parameters with the processes associated with the other clusters. We fixed the expected become-uninfectious rate to 36.5 per year, which corresponds to an average time to becoming uninfectious of 10 days. We assumed an HKY (*28*) nucleotide substitution model with 4 gamma rate categories to account for site-to-site rate heterogeneity (*29*). We assumed a strict clock with a rate fixed to $8x10^{-4}$ substitutions per site per year. The effective reproductive number (Fig. 4a) and the sampling proportion (Fig. 4b) were allowed to vary in a piecewise-constant fashion in intervals of 1 week. An independent smoothing prior was applied to the natural logarithm of each of these time-varying parameters (*30*). So as to constrain both the relative sizes of the change between weeks and the absolute reproductive number and sampling proportion values, we used smoothing priors formulated as the product between a) a Gaussian penalty on the differences between the logarithm of values for adjacent weeks and b) a function of the value taken in each week. For this function, we used the probability density of a LogNormal(0.8, 0.5) distribution in the case of the Re values, and the density of a Beta(1, 99) distribution in the case of the sampling proportion values. An Exp(1) hyperprior was applied to the variance of the Gaussian component of each smoothing prior.

### Supplementary Text

Sampling saturation

To evaluate the completeness of our sample set, we ask whether we could expect to detect even more introductions with additional sampling. We first consider all Swiss sequences and record the number of Swiss transmission chains and Swiss singletons inferred by our clustering procedure. Then we randomly subsample 90%, 80%...10% of the Swiss sequences and record how many of the Swiss transmission chains and Swiss singletons were sampled. From Fig. S2a, we observe that under the “minimum introductions, maximum Swiss transmission” assumption, the number of Swiss introductions does not saturate as we sample more sequences, thus we speculate that we did not sample the majority of imports. This conclusion is even stronger under the “maximum introductions, minimum Swiss transmission” assumption (Fig. S2b).

Evaluation of Test, Trace, quarantine, isolate program

To evaluate how well Swiss introductions were contained after detection, we counted the number of secondary cases arising more than 10 days after the index case was tested positive. We assume that it takes 10 days from infection to case confirmation, so all confirmed cases testing positive within 10 days of a positive test of the index case were infected before the transmission chain was detected (i.e. before the positive test of the index case). Importantly, these infections stemming from imported outbreaks cannot be avoided by contact tracing. On the other hand, in an ideal test, trace, and isolate scenario in which we could immediately identify and isolate all contacts of the index case (and contacts of contacts, as necessary), we would avoid all cases which are confirmed more than 10 days after the index case testing positive. Of course, these contact tracing demands are unrealistic, particularly if there was a large amount of spread before the index case was tested.

Assessing epidemic spread between Swiss cantons

To test whether sequences from the different Swiss cantons clustered together in the same transmission chains more or less often than we would expect by chance, we performed two different permutation tests. To test for more-than-expected-mixing, we permuted canton labels under a model in which the maximum number of mixing events were allowed, meaning we assumed each individual in a mixed-canton transmission chain was equally likely to have been infected from another canton. To test for less-than-expected mixing, we permuted under a model in which the minimum number of mixing events was allowed, meaning we assumed that only one individual per canton in mixed-canton transmission chains was infected from another canton. Practically, this means we ran one test in which we permuted individuals and one test in which we collapsed individuals from the same canton in the same transmission chain to a single unit before permuting. We permuted canton labels across only transmission chains consisting of at least two cantons and keep the permutation if all transmission chains are again composed of at least two cantons.

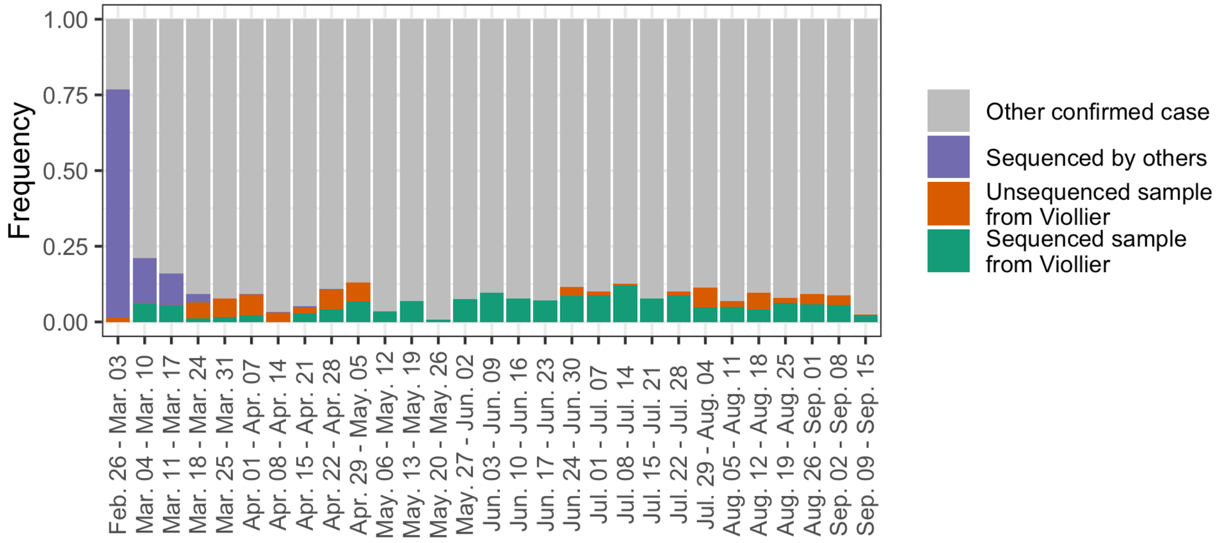


**Fig. S1.**

Summary of Swiss sequencing efforts. We tally the number of sequences generated each week from our nation-wide genome sequencing project, the number of not-yet sequenced samples we received from the diagnostics company Viollier, and the number of sequences generated by others and submitted to GISAID. These tallies are displayed as a fraction of the number of confirmed cases each week.

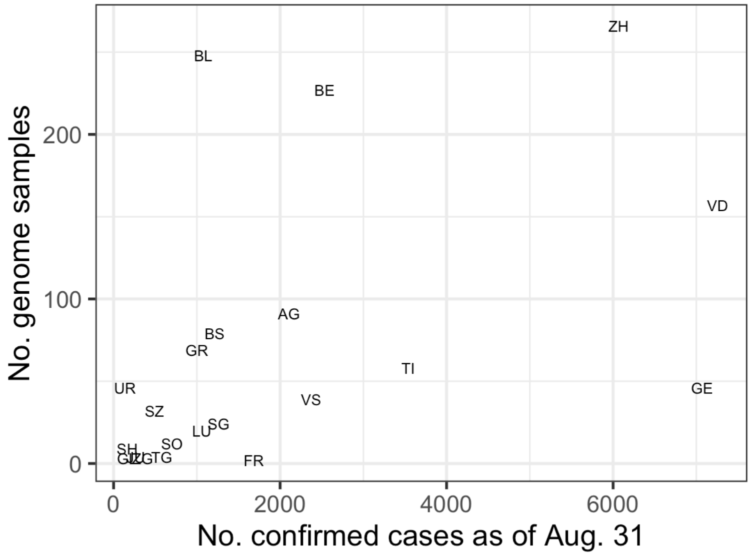


**Fig. S2.**

Representativeness of our nation-wide genome sequencing project at the cantonal level. The number of genome sequences we generated (y-axis) is plotted against the cumulative number of confirmed cases from each canton as of Aug. 31 (x-axis). Points are labelled with standard abbreviations of the canton names.

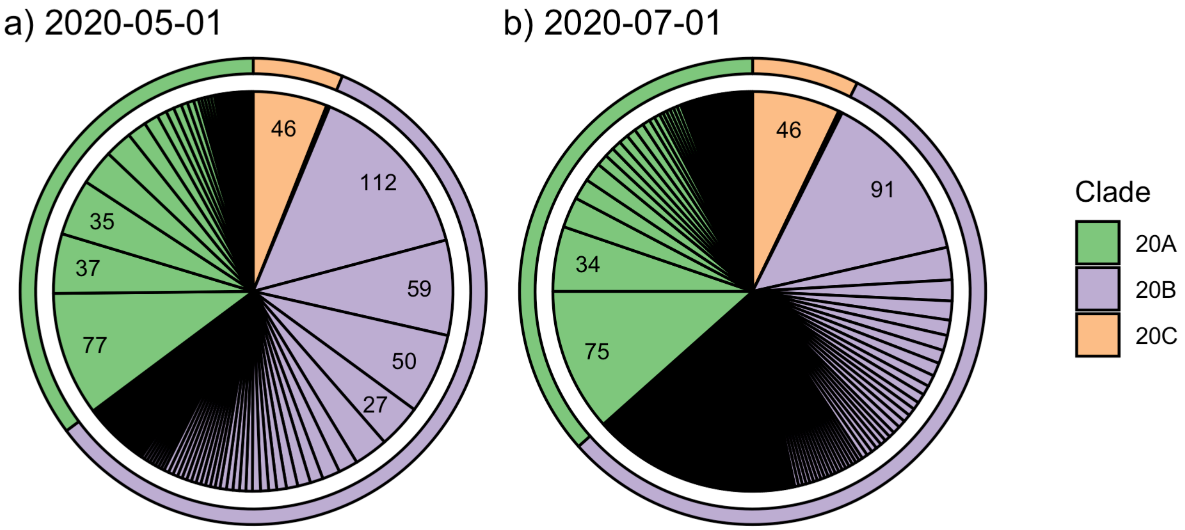


**Fig. S3.**
The number of Swiss genome sequences sampled during the summer epidemic that were generated by different lineages from the early epidemic. These plots are equivalent to Fig. 1b but use **(a)** May. 1 and **(b)** Jul. 1 instead of Jun. 1 as a cut-off between the early and summer epidemic.

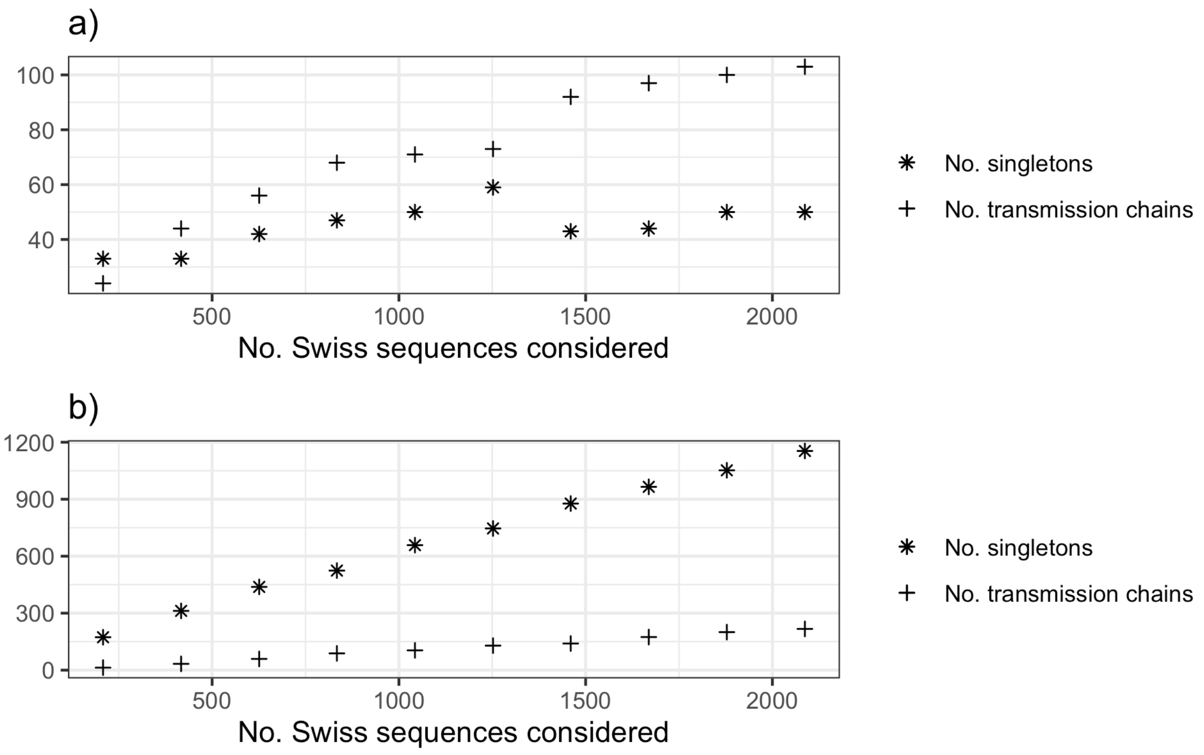


**Fig. S4.**
The number of Swiss introductions (singletons and transmission chains) identified on a fixed tree with increasing numbers of randomly selected Swiss sequences considered. **(a)** shows introductions inferred under the "minimum introductions, maximum local transmission" assumption while **(b)** shows introductions inferred under the "maximum introductions, minimum local transmission" assumption. See Materials and Methods for further details.

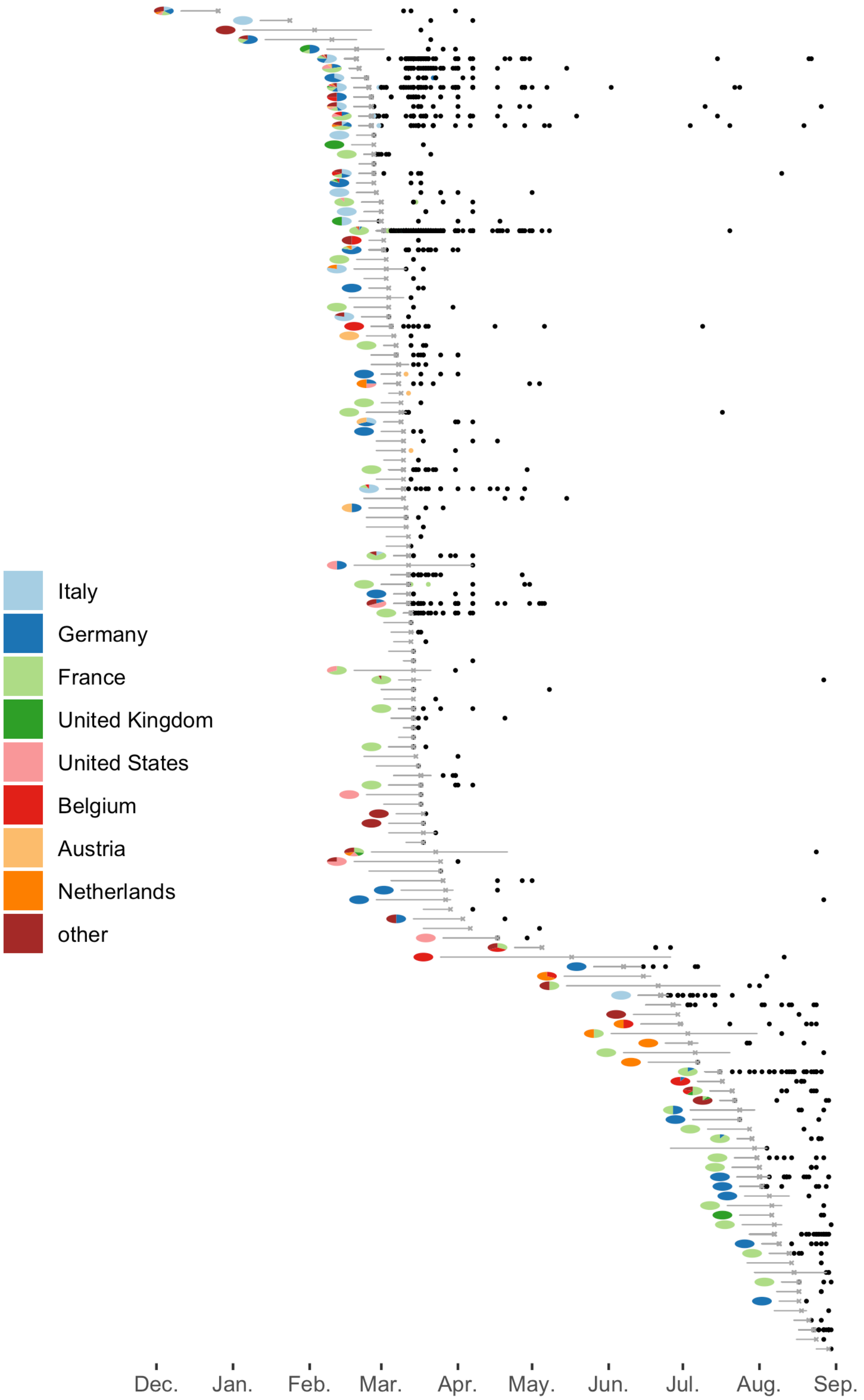


**Fig. S5.**
The analog to main text Fig. 2a for Swiss singletons. Here, the black points aligned horizontally are singletons descending from the same attachment point.

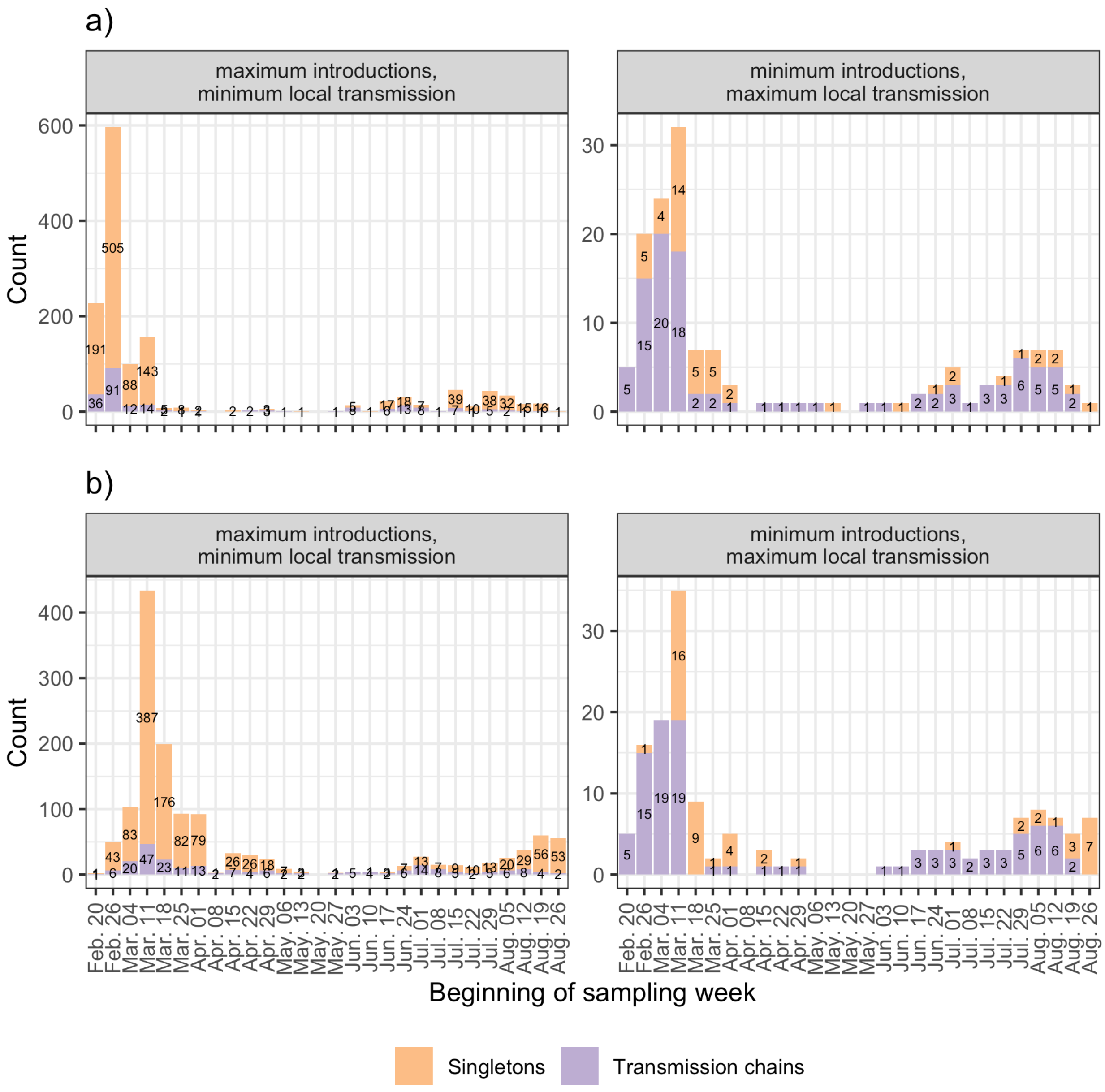


**Fig. S6**

Least-square estimates for **(a)** the attachment times of Swiss introductions to the rest of the tree and **(b)** Swiss transmission chain MRCAs and Swiss singleton sample dates. The two facets **c**orrespond to two different assumptions used to define Swiss transmission chains. The numbers give the number of introductions and transmission chains, respectively.

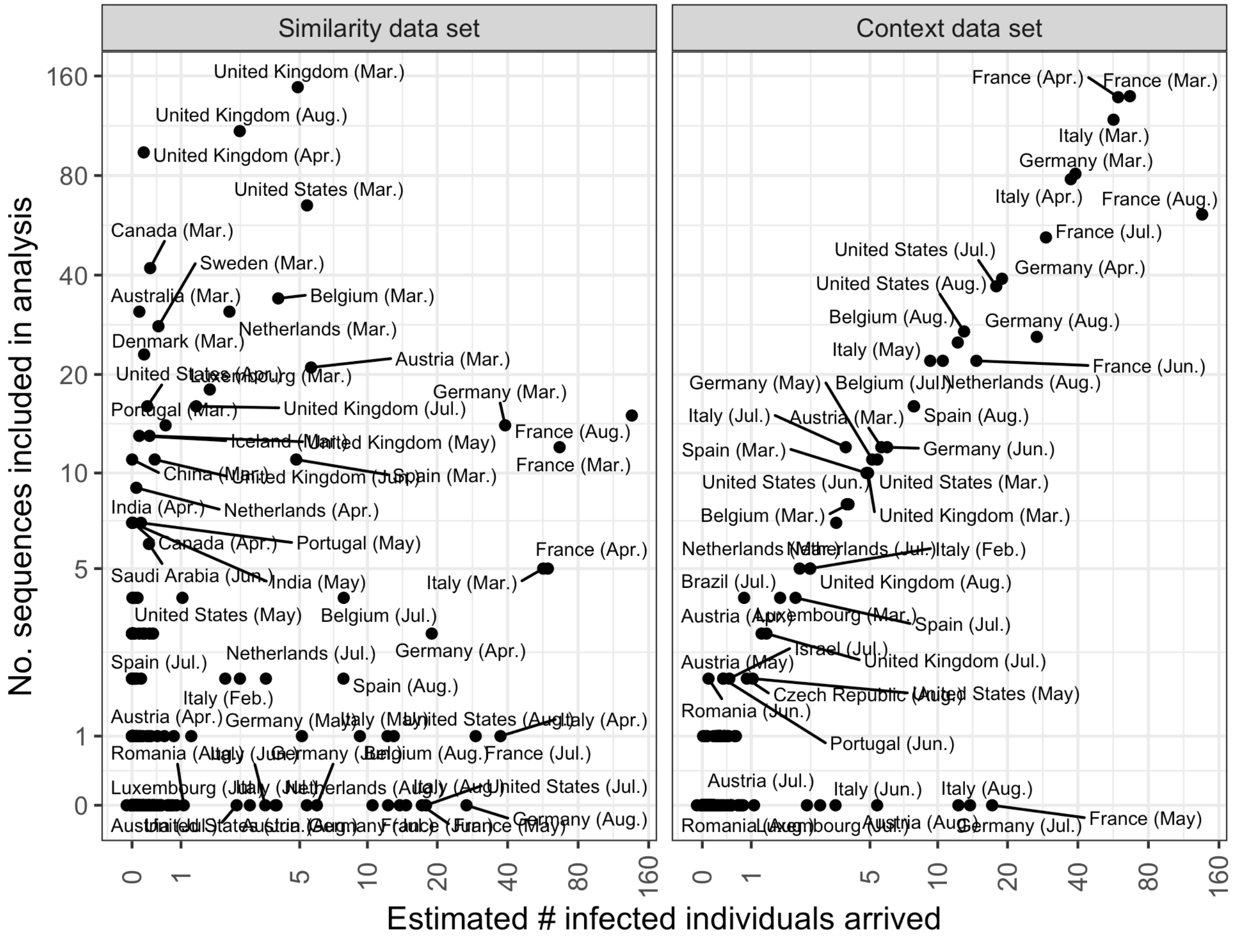


**Fig. S7.**

The number of genome sequences from each dataset included in our analysis plotted against the estimated number of infected individuals arriving in Switzerland from each source country until Aug. 2020. The left panel shows sequences included for being genetically similar to a Swiss sequence and the right panel shows sequences selected based on the estimated number of arrivals of infected individuals into Switzerland each month. Both the “similarity” and “context” datasets were used to identify Swiss introductions. Only the “context” dataset was used to estimate the source location of introductions. Since we lack genome sequences from some of Switzerland’s neighboring countries during a few summer months, we may underestimate the contributions of these countries around this time.

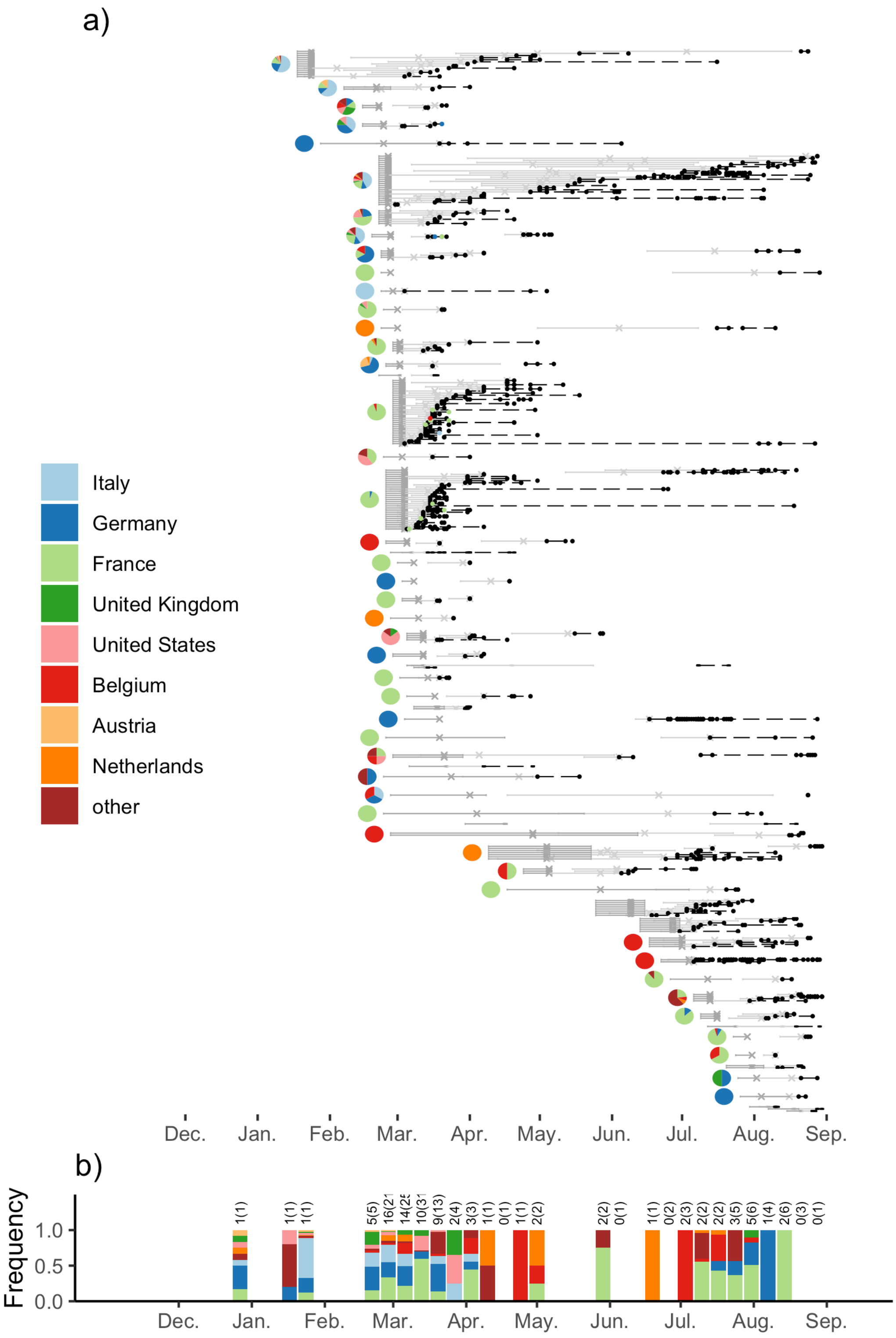


**Fig. S8.**

This plot is equivalent to Fig. 2 but uses replicate 2 of context sequences. See Methods and Materials for further details.

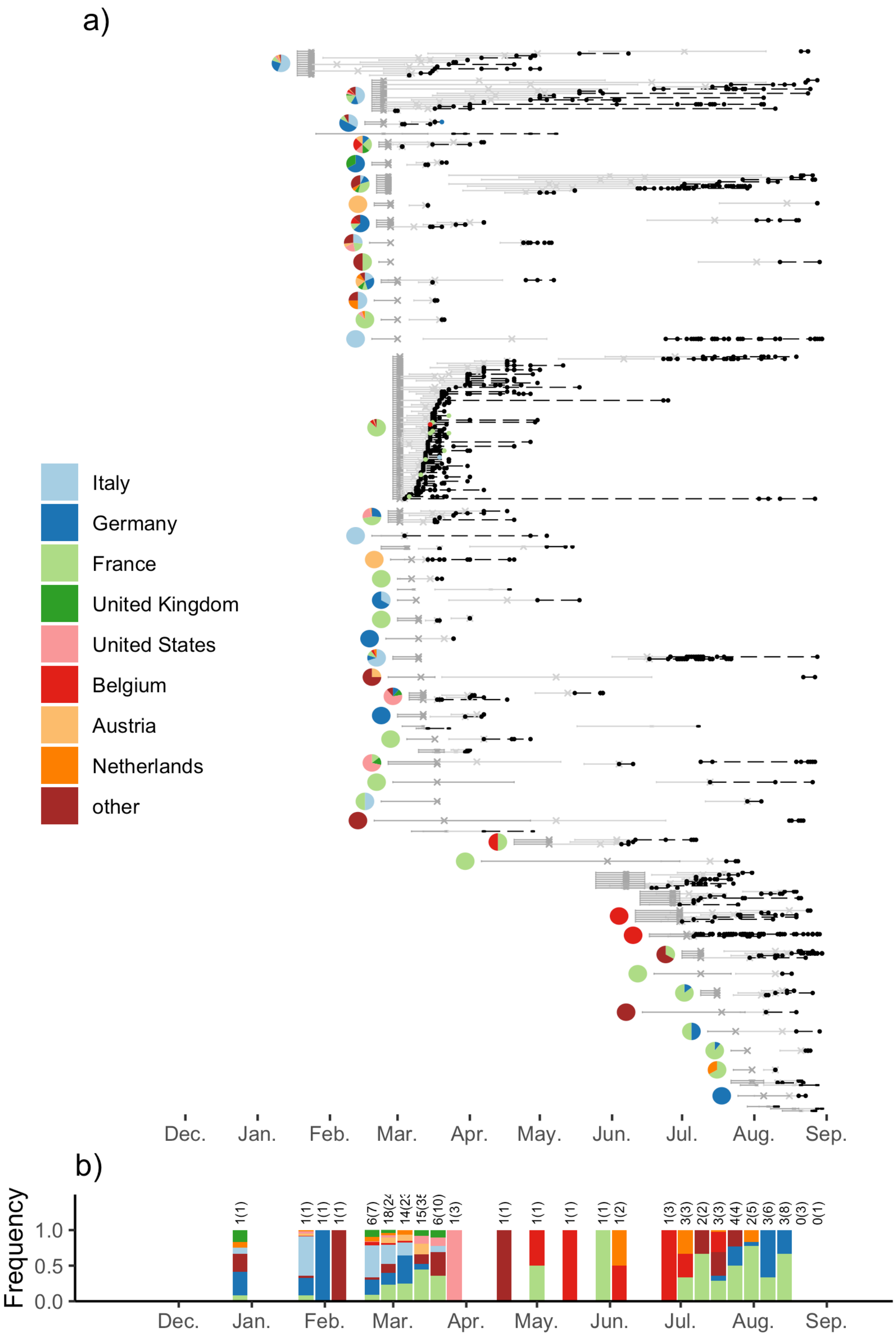


**Fig. S9.**

This plot is equivalent to Fig. 2 but uses replicate 3 of context sequences. See Methods and Materials for further details.

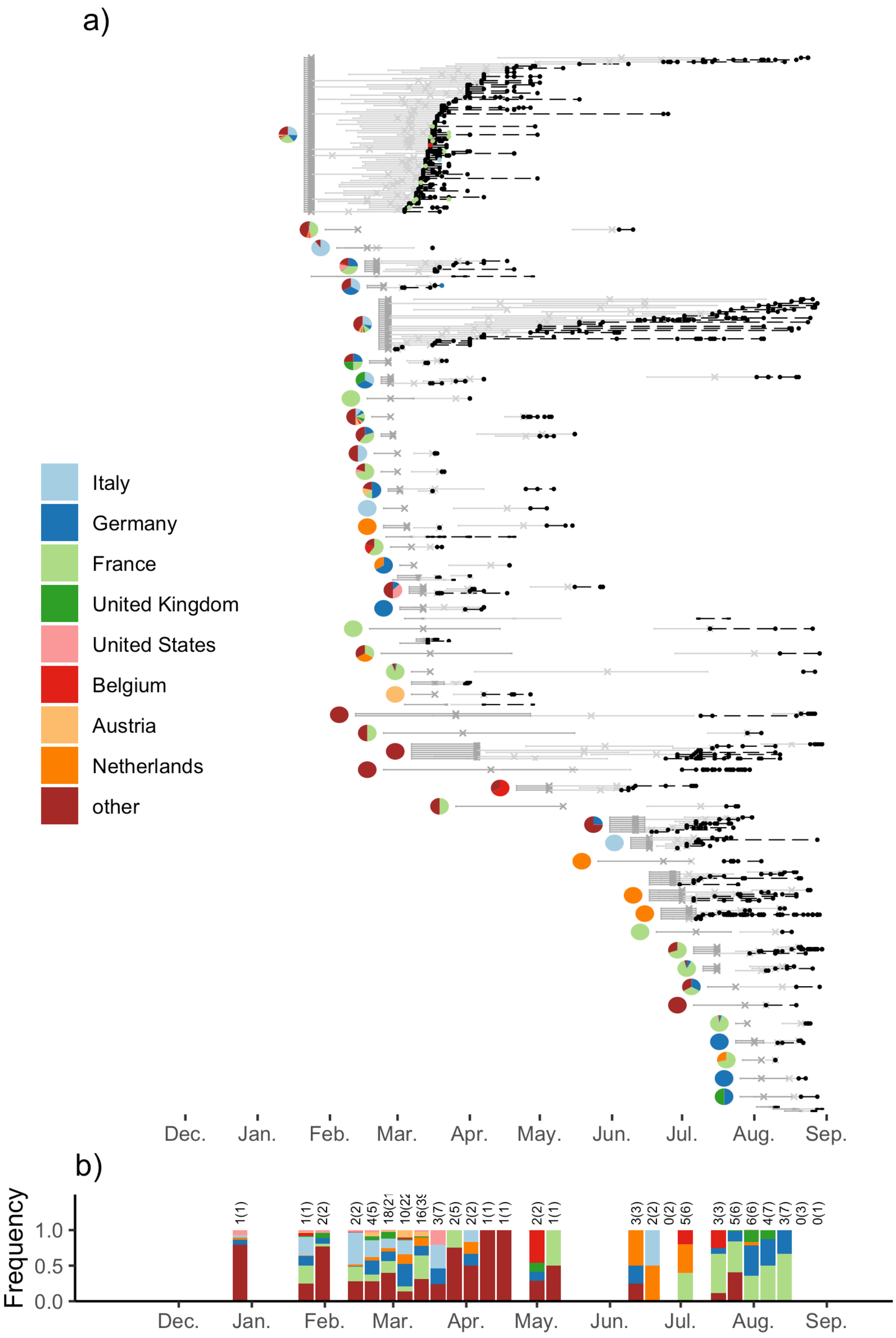


**Fig. S10.**

This plot is equivalent to Fig. 2 but uses context sequences based on padding estimated imports. See Methods and Materials for further details.

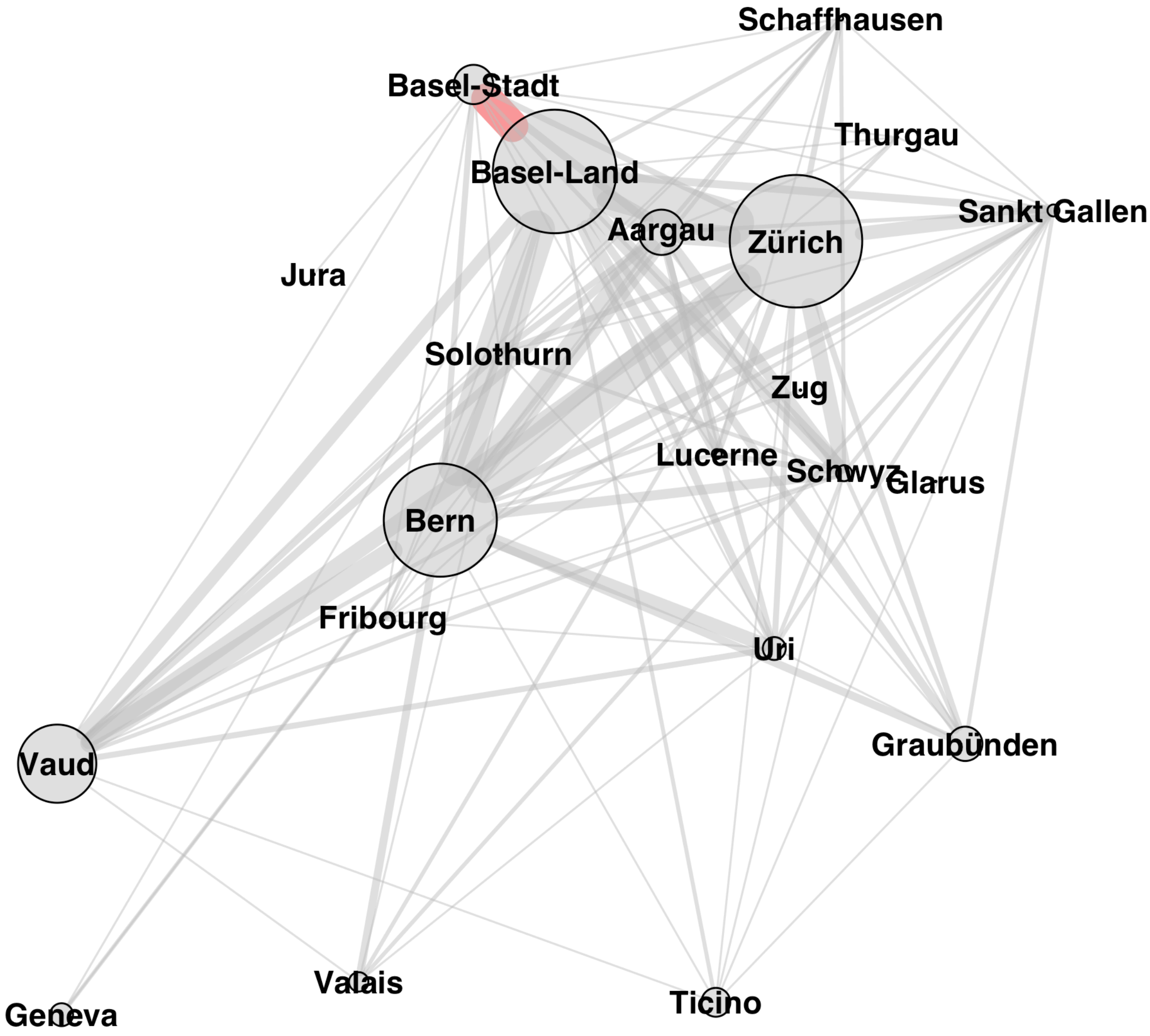


**Fig. S11.**

Network depicting the number of times cantons cluster together in the same transmission chain. This plot is based on sequences from our nation-wide genome sequencing project. Edge weights are proportional to the number of transmission chains containing sequences from both cantons. Edge colors indicate significantly more or less mixing than expected based on the permutation tests described in the Materials and Methods. The red edge shows that the neighboring cantons of Basel-Stadt and Basel-Land mixed significantly more than expected based on chance (these cantons are found together in 16 transmission chains, which is more often than in 90% of our permutations assuming maximal mixing). None of the cantons mixed less than expected.

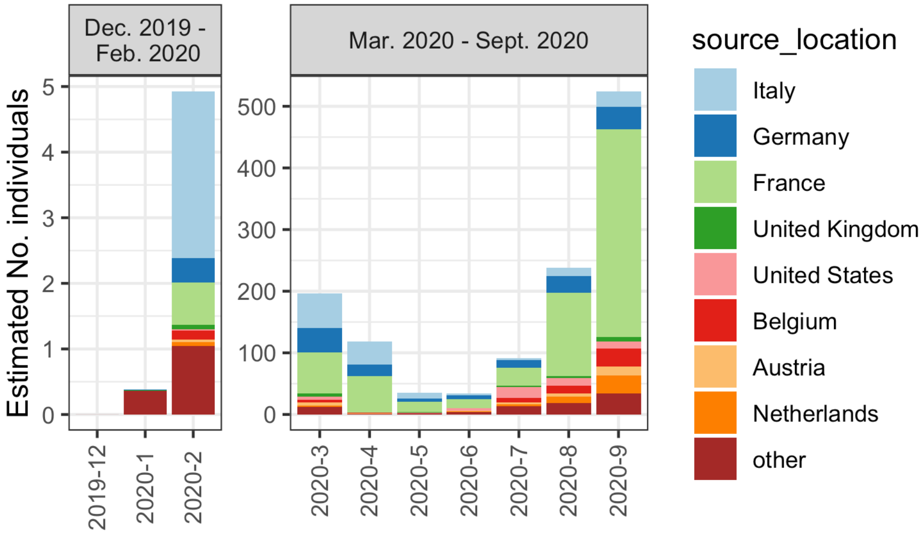


**Fig. S12.**
The estimated number of infected individuals arriving in Switzerland by month and source country. These estimates are generated by multiplying the number of arrivals from each country in a month by the average proportion of the source population infectious on any given day in that month. The values are intended only to be proportional to the expected number of arrivals.

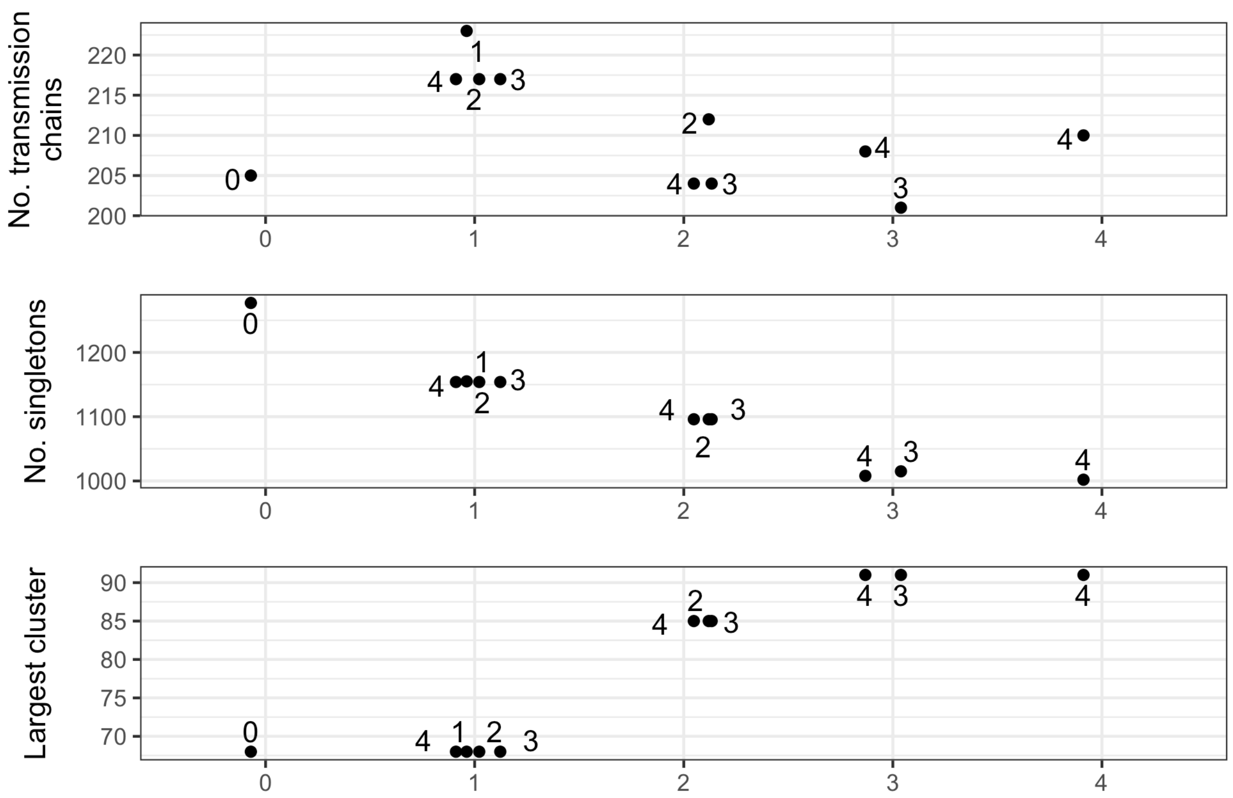


**Fig. S13.**

Summary statistics of inferred introductions with varying clustering criteria. The column represents the maximal number of exports along a single Swiss branch (criterium (b) in cluster definition). The numeric label corresponds to the maximal number of exports allowed when defining a Swiss transmission chain (criterium (a) in cluster definition).

**Table S1.**

Details of genome sequence quality control filtering.

| Criteria | No. sequences |
|---|---|
| Total sequences downloaded and aligned | 136801 |
| Filtering of non-Swiss sequences:<br>Excluded based on non-human sample, < 27,000bp, incomplete date, or in Nextstrain exclude document | -11097 |
| Filtering of Swiss sequences:<br>Excluded based on non-human sample, < 20,000bp, incomplete date, or in Nextstrain exclude document | -228 |
| Excluded based on Nextstrain diagnostic check | -318 |
| Total sequences considered for analysis | 125158 |

**Table S2.**
GISAID accession identifiers for Swiss sequences generated as part of this study.

| Accession ID range start | Accession ID range end |
|---|---|
| EPI_ISL_451668 | EPI_ISL_451916 |
| EPI_ISL_466927 | EPI_ISL_467016 |
| EPI_ISL_468202 | EPI_ISL_468304 |
| EPI_ISL_476079 | EPI_ISL_476134 |
| EPI_ISL_483648 | EPI_ISL_483685 |
| EPI_ISL_486442 | EPI_ISL_486538 |
| EPI_ISL_489959 | EPI_ISL_489986 |
| EPI_ISL_498619 | EPI_ISL_498627 |
| EPI_ISL_500877 | EPI_ISL_500945 |
| EPI_ISL_510690 | EPI_ISL_510809 |
| EPI_ISL_511984 | EPI_ISL_512054 |
| EPI_ISL_516552 | EPI_ISL_516607 |
| EPI_ISL_523856 | EPI_ISL_523926 |
| EPI_ISL_535585 | EPI_ISL_535649 |
| EPI_ISL_539341 | EPI_ISL_539482 |
| EPI_ISL_541401 | EPI_ISL_541539 |
| EPI_ISL_560416 | EPI_ISL_560553 |

**Table S3.**
GISAID acknowledgments table.

| Originating Laboratory | Submitting Laboratory | Authors |
|---|---|---|
| Accession IDs | | |
| [Romania, Bucharest] National Institute for Infectious Diseases “Prof. Dr. Matei Balș” | [Romania, Bucharest] National Institute for Infectious Diseases “Prof. Dr. Matei Balș” | Leontina Banica et al |
| EPI_ISL_468149 | | |
| Agenzia di Tutela della Salute di Bergamo | Istitituto Zooprofilattico Sperimentale dell'Abruzzo e Molise "G.Caporale" | Lorusso A et al |
| EPI_ISL_528934, EPI_ISL_528936, EPI_ISL_528937, EPI_ISL_528938, EPI_ISL_528939, EPI_ISL_528941, EPI_ISL_528942, EPI_ISL_528943, EPI_ISL_528944, EPI_ISL_528945, EPI_ISL_528946, EPI_ISL_528947, EPI_ISL_528948, EPI_ISL_528949 | | |
| Alaska State Virology Laboratory | Alaska State Virology Laboratory | Chen J et al with Pathogenomics group Dagdag R et al |
| EPI_ISL_528515, EPI_ISL_528517, EPI_ISL_528526 | | |
| Alsafar - Khalifa University Abu Dhabi | Alsafar - Khalifa University Abu Dhabi | Andreas Henschel et al |
| EPI_ISL_528691, EPI_ISL_528694, EPI_ISL_528707 | | |
| Amedeo di savoia | Crosetto lab, Karolinska Institutet, SciLifeLab | Michele Simonetti et al |
| EPI_ISL_569868, EPI_ISL_569871, EPI_ISL_569875, EPI_ISL_569876, EPI_ISL_569877, EPI_ISL_569878, EPI_ISL_569879, EPI_ISL_569880, EPI_ISL_569881, EPI_ISL_569882, EPI_ISL_569883, EPI_ISL_569865, EPI_ISL_569884, EPI_ISL_569885, EPI_ISL_569886 | | |
| AR Dept. of Health-Public Health Lab | Pathogen Discovery, Respiratory Viruses Branch, Division of Viral Diseases, Centers for Disease Control and Prevention | Yan Li et al |
| EPI_ISL_535651 | | |
| AR Dept. of Health-Public Health Lab | Pathogen Discovery, Respiratory Viruses Branch, Division of Viral Diseases, Centers for Disease Control and Prevention | Ying Tao et al |

| | | |
|---|---|---|
| EPI_ISL_527632 | | |
| Area of Virology, Serology and Virology Division (SAViD), New South Wales Health Pathology Randwick | Area of Virology, Serology and Virology Division (SAViD), New South Wales Health Pathology Randwick | Rawlinson et al |
| EPI_ISL_509492, EPI_ISL_500607, EPI_ISL_500608, EPI_ISL_509493, EPI_ISL_500609, EPI_ISL_500611, EPI_ISL_527008, EPI_ISL_500630, EPI_ISL_500634, EPI_ISL_500674, EPI_ISL_500695, EPI_ISL_527015, EPI_ISL_527016 | | |
| ARS Algarve - Laboratório Laura Ayres | Instituto Nacional de Saude (INSA) | Guiomar et al et al |
| EPI_ISL_418001 | | |
| ASST GOM Niguarda | Dep. Of Oncology and Hemato-Oncology University of Milan | Claudia Alteri et al |
| EPI_ISL_542098, EPI_ISL_542099, EPI_ISL_542100, EPI_ISL_542101, EPI_ISL_542103, EPI_ISL_542105, EPI_ISL_542110, EPI_ISL_542111, EPI_ISL_542112, EPI_ISL_542115, EPI_ISL_542117, EPI_ISL_542118, EPI_ISL_542119, EPI_ISL_542122, EPI_ISL_542124, EPI_ISL_542126, EPI_ISL_542128, EPI_ISL_542129, EPI_ISL_542130, EPI_ISL_542131, EPI_ISL_542132, EPI_ISL_542133, EPI_ISL_542134, EPI_ISL_542135, EPI_ISL_542137, EPI_ISL_542140, EPI_ISL_542141, EPI_ISL_542142, EPI_ISL_542144, EPI_ISL_542148, EPI_ISL_542150, EPI_ISL_542151, EPI_ISL_542153, EPI_ISL_542155, EPI_ISL_542157, EPI_ISL_542158, EPI_ISL_542160, EPI_ISL_542162, EPI_ISL_542163, EPI_ISL_542164, EPI_ISL_542166, EPI_ISL_542167, EPI_ISL_542168, EPI_ISL_542169, EPI_ISL_542173, EPI_ISL_542174, EPI_ISL_542175, EPI_ISL_542176, EPI_ISL_542180, EPI_ISL_542182, EPI_ISL_542183, EPI_ISL_542184, EPI_ISL_542186, EPI_ISL_542187, EPI_ISL_542188, EPI_ISL_542189, EPI_ISL_542190, EPI_ISL_542191, EPI_ISL_542192, EPI_ISL_542193, EPI_ISL_542195, EPI_ISL_542198, EPI_ISL_542200, EPI_ISL_542203, EPI_ISL_542205, EPI_ISL_542207, EPI_ISL_542208, EPI_ISL_542210, EPI_ISL_542211, EPI_ISL_542215, EPI_ISL_542228, EPI_ISL_542229, EPI_ISL_542236, EPI_ISL_542238, EPI_ISL_542242, EPI_ISL_542243, EPI_ISL_542245, EPI_ISL_542246, EPI_ISL_542254, EPI_ISL_542258, EPI_ISL_542264, EPI_ISL_542265, EPI_ISL_542266, EPI_ISL_542267, EPI_ISL_542268, EPI_ISL_542269, EPI_ISL_542270, EPI_ISL_542272, EPI_ISL_542274, EPI_ISL_542275, EPI_ISL_542276, EPI_ISL_542400, EPI_ISL_542402, EPI_ISL_542404, EPI_ISL_542405, EPI_ISL_542407, EPI_ISL_542413, EPI_ISL_542415, EPI_ISL_542417, EPI_ISL_542418, EPI_ISL_542420, EPI_ISL_542423, EPI_ISL_542425, EPI_ISL_542426, EPI_ISL_542428, EPI_ISL_542430, EPI_ISL_542431, EPI_ISL_542432, EPI_ISL_542434, EPI_ISL_542437, EPI_ISL_542438, EPI_ISL_542440, EPI_ISL_542443 | | |
| Austrian Agency for Health and Food Safety (AGES) | Bergthaler laboratory, CeMM Research Center for Molecular Medicine of the Austrian Academy of Sciences | Alexandra Popa et al |
| EPI_ISL_475830, EPI_ISL_475838, EPI_ISL_475839, EPI_ISL_475841, EPI_ISL_475842, EPI_ISL_475849, EPI_ISL_475855, EPI_ISL_475863, EPI_ISL_475870, EPI_ISL_475873, EPI_ISL_475886 | | |

| | | |
|---|---|---|
| Avera Mckennan Laboratory | Minnesota Department of Health, Public Health Laboratory | Matt Plumb et al |
| EPI_ISL_507963, EPI_ISL_507965 | | |
| B.J. Medical College and Civil hospital | Gujarat Biotechnology Research Centre | Afzal Ansari et al |
| EPI_ISL_461503 | | |
| B.J. Medical College and Civil hospital | Gujarat Biotechnology Research Centre | Zuber Saiyed et al |
| EPI_ISL_467048 | | |
| Bamrasnaradura Hospital | 1. Department of Medical Sciences, Ministry of Public Health, Thailand 2. Thai Red Cross Emerging Infectious Diseases - Health Science Centre 3. Department of Disease Control, Ministry of Public Health, Thailand | Pilailuk et al |
| EPI_ISL_403962 | | |
| Barts Health NHS Trust | Wellcome Sanger Institute for the COVID-19 Genomics UK Consortium | Teresa Cutino-Moguel et al |
| EPI_ISL_541778 | | |
| BCCDC Public Health Laboratory | BCCDC Public Health Laboratory | Harrigan et al |
| EPI_ISL_418849, EPI_ISL_418857 | | |
| Biology Dpt | Microbiology and Infections Diseases | Emmanuelle Billon-Denis et al |
| EPI_ISL_505003 | | |
| Biomedical Sciences and Public Health, Polytechnic University of Marche | Biomedical Sciences and Public Health, Polytechnic University of Marche | Bagnarelli et al |
| EPI_ISL_516079, EPI_ISL_516081, EPI_ISL_516082, EPI_ISL_516086 | | |
| Bülach Hospital | Institute of Medical Virology, University of Zurich | Stefan Schmutz et al |
| EPI_ISL_524476 | | |

| | | |
|---|---|---|
| Bundeswehr Institute of Microbiology | Bundeswehr Institute of Microbiology | Mathias C Walter et al |
| EPI_ISL_414521 | | |
| Cabinet médical | National Reference Center for Viruses of Respiratory Infections, Institut Pasteur, Paris | Mélanie Albert et al |
| EPI_ISL_418235 | | |
| Cabinet Médical | National Reference Center for Viruses of Respiratory Infections, Institut Pasteur, Paris | Mélanie Albert et al |
| EPI_ISL_443301, EPI_ISL_428364 | | |
| Caloundra Hospital | Public Health Virology Laboratory | Bixing Huang et al |
| EPI_ISL_447594 | | |
| Cantonal Hospital Winterthur | Institute of Medical Virology, University of Zurich | Stefan Schmutz et al |
| EPI_ISL_524478 | | |
| Capio S:t Gorans sjukhus | The Public Health Agency of Sweden | Anna-Malin Linde et al |
| EPI_ISL_534230, EPI_ISL_534231 | | |
| Cedars-Sinai Medical Center, Department of Pathology & Laboratory Medicine, Molecular Pathology Laboratory | Cedars-Sinai Medical Center, Molecular Pathology Laboratory of Department of Pathology & Laboratory Medicine and Genomic Core | Wenjuan Zhang et al |
| EPI_ISL_475601, EPI_ISL_475679 | | |
| Center for Diagnostics, Institute of Medical Microbiology, Virology and Hygiene | University Medical Center Hamburg-Eppendorf | Huang et al |
| EPI_ISL_450413 | | |
| Center for Genome Regulation (CRG) | Center for Mathematical Modeling and Center for Genome Regulation. Santiago, Chile | Gaete A et al |
| EPI_ISL_468760 | | |
| Center for Virology, Medical University of Vienna | Bergthaler laboratory, CeMM Research Center for Molecular | Alexandra Popa et al |

| | | |
|---|---|---|
| | Medicine of the Austrian Academy of Sciences | |
| EPI_ISL_419673, EPI_ISL_437997, EPI_ISL_438013, EPI_ISL_438016, EPI_ISL_438026, EPI_ISL_438030, EPI_ISL_438033, EPI_ISL_438036, EPI_ISL_438045, EPI_ISL_438060, EPI_ISL_438072, EPI_ISL_438079, EPI_ISL_438082, EPI_ISL_438087, EPI_ISL_438089, EPI_ISL_438096, EPI_ISL_438108, EPI_ISL_438115, EPI_ISL_438116, EPI_ISL_438117, EPI_ISL_475789, EPI_ISL_475798, EPI_ISL_475809, EPI_ISL_438124 | | |
| Center of Medical Microbiology, Virology, and Hospital Hygiene, University of Duesseldorf | Center of Medical Microbiology, Virology, and Hospital Hygiene, Heinrich Heine University Düsseldorf | Maximilian Damagnez et al |
| EPI_ISL_523950 | | |
| Center of Medical Microbiology, Virology, and Hospital Hygiene, University of Duesseldorf | Center of Medical Microbiology, Virology, and Hospital Hygiene, University of Duesseldorf | Maximilian Damagnez et al |
| EPI_ISL_523929, EPI_ISL_523931, EPI_ISL_523932, EPI_ISL_523934, EPI_ISL_523935, EPI_ISL_523936, EPI_ISL_523937, EPI_ISL_523938, EPI_ISL_523939, EPI_ISL_523940, EPI_ISL_523943, EPI_ISL_523944, EPI_ISL_523945, EPI_ISL_523946, EPI_ISL_523947, EPI_ISL_523948, EPI_ISL_523949 | | |
| Center of Medical Microbiology, Virology, and Hospital Hygiene, University of Duesseldorf | Center of Medical Microbiology, Virology, and Hospital Hygiene, University of Duesseldorf | Ortwin Adams et al |
| EPI_ISL_414507, EPI_ISL_414505, EPI_ISL_414509, EPI_ISL_417458, EPI_ISL_417459, EPI_ISL_417460, EPI_ISL_417461, EPI_ISL_417462, EPI_ISL_417463, EPI_ISL_417464, EPI_ISL_417465, EPI_ISL_417466, EPI_ISL_417468, EPI_ISL_419541, EPI_ISL_419542, EPI_ISL_419543, EPI_ISL_419544, EPI_ISL_419545, EPI_ISL_419546, EPI_ISL_419547, EPI_ISL_419548, EPI_ISL_419549, EPI_ISL_419550, EPI_ISL_419551, EPI_ISL_419552, EPI_ISL_425120, EPI_ISL_425122, EPI_ISL_425124, EPI_ISL_425125, EPI_ISL_425127, EPI_ISL_425128, EPI_ISL_425126, EPI_ISL_425138, EPI_ISL_425139, EPI_ISL_425129, EPI_ISL_425130, EPI_ISL_425131, EPI_ISL_425132, EPI_ISL_425133, EPI_ISL_425134, EPI_ISL_425135, EPI_ISL_425136, EPI_ISL_425137, EPI_ISL_452180, EPI_ISL_425140 | | |
| Center of Medical Microbiology, Virology, and Hospital Hygiene, University of Duesseldorf | Universitätstr.1 40225 Düsseldorf Germany | Maximilian Damagnez et al |
| EPI_ISL_523930 | | |
| Centogene AG | Centogene AG | Prof. Dr. Peter Bauer et al |
| EPI_ISL_459962, EPI_ISL_459964, EPI_ISL_459963, EPI_ISL_457750 | | |

| | | |
|---|---|---|
| Centre de SSS de la Haute-Yamaska | Laboratoire de santé publique du Québec | Sandrine Moreira et al |
| EPI_ISL_535805 | | |
| Centre for Clinical Infection and Diagnostics Research and Genomics Innovation Unit, Guy's and St. Thomas' NHS Trust | COVID-19 Genomics UK (COG-UK) Consortium | Chloe Fisher et al |
| EPI_ISL_484130, EPI_ISL_484200 | | |
| Centre for Enzyme Innovation, University of Portsmouth / Translational Research Laboratory, Portsmouth Hospitals NHS Trust | COVID-19 Genomics UK (COG-UK) Consortium | Angela Beckett et al |
| EPI_ISL_475272, EPI_ISL_475293, EPI_ISL_479179, EPI_ISL_526434, EPI_ISL_507131, EPI_ISL_499308, EPI_ISL_507143, EPI_ISL_517511, EPI_ISL_517512 | | |
| Centre for Infectious Diseases and Microbiology Public Health | NSW Health Pathology - Institute of Clinical Pathology and Medical Research; Westmead Hospital; University of Sydney | Bachmann N et al |
| EPI_ISL_427758 | | |
| Centre for Infectious Diseases and Microbiology Public Health | NSW Health Pathology - Institute of Clinical Pathology and Medical Research; Westmead Hospital; University of Sydney | O'Sullivan MV et al |
| EPI_ISL_417394 | | |
| Centre for Infectious Diseases and Microbiology Public Health | NSW Health Pathology - Institute of Clinical Pathology and Medical Research; Westmead Hospital; University of Sydney | Sadsad R et al |
| EPI_ISL_417407 | | |
| Centre Hositalier Universitaire de Rouen Laboratoire de Virologie | National Reference Center for Viruses of Respiratory Infections, Institut Pasteur, Paris | Mélnie Albert et al |
| EPI_ISL_416494 | | |
| Centre Hospitalier Alpes Leman | CNR Virus des Infections Respiratoires - France SUD | Antonin Bal et al |
| EPI_ISL_509012 | | |
| Centre hospitalier Anna-Laberge | Laboratoire de santé publique du Québec | Sandrine Moreira et al |

| | | |
|---|---|---|
| EPI_ISL_535960, EPI_ISL_535980, EPI_ISL_536159, EPI_ISL_536220, EPI_ISL_536241 | | |
| Centre Hospitalier Compiègne Laboratoire de Biologie | National Reference Center for Viruses of Respiratory Infections, Institut Pasteur, Paris | Mélanie Albert et al |
| EPI_ISL_429968, EPI_ISL_418221, EPI_ISL_418223, EPI_ISL_418225, EPI_ISL_418236, EPI_ISL_418238, EPI_ISL_418239 | | |
| Centre Hospitalier Compiègne Laboratoire de Biologie | National Reference Center for Viruses of Respiratory Infections, Institut Pasteur, Paris | Mélnie Albert et al |
| EPI_ISL_414629, EPI_ISL_414634, EPI_ISL_414637, EPI_ISL_415654, EPI_ISL_416495, EPI_ISL_416497 | | |
| Centre Hospitalier de Bourg en Bresse | CNR Virus des Infections Respiratoires - France SUD | Antonin Bal et al |
| EPI_ISL_508938, EPI_ISL_508944, EPI_ISL_525539, EPI_ISL_419185, EPI_ISL_419186, EPI_ISL_508968, EPI_ISL_417340 | | |
| Centre Hospitalier de Bourg en Bresse | CNR Virus des Infections Respiratoires - France SUD | Bal et al |
| EPI_ISL_416757 | | |
| Centre Hospitalier de Macon | CNR Virus des Infections Respiratoires - France SUD | Antonin Bal et al |
| EPI_ISL_508946, EPI_ISL_508876, EPI_ISL_419174, EPI_ISL_419175, EPI_ISL_509005, EPI_ISL_508949, EPI_ISL_419187, EPI_ISL_420614, EPI_ISL_508952 | | |
| Centre Hospitalier de Valence | CNR Virus des Infections Respiratoires - France SUD | Antonin Bal et al |
| EPI_ISL_508881, EPI_ISL_419168 | | |
| Centre Hospitalier de Valence | CNR Virus des Infections Respiratoires - France SUD | Bal et al |
| EPI_ISL_416749 | | |
| Centre Hospitalier de Villefranche | CNR Virus des Infections Respiratoires - France SUD | Antonin Bal et al |
| EPI_ISL_508978, EPI_ISL_508932 | | |
| Centre Hospitalier du Haut-Bugey | CNR Virus des Infections Respiratoires - France SUD | Antonin Bal et al |
| EPI_ISL_509016 | | |
| Centre Hospitalier Lucien Hussel | CNR Virus des Infections Respiratoires - France SUD | Antonin Bal et al |
| EPI_ISL_508947 | | |
| Centre hospitalier Métropole Savoie | CNR Virus des Infections Respiratoires - France SUD | Antonin Bal et al |

| | | |
|---|---|---|
| EPI_ISL_508934, EPI_ISL_508935, EPI_ISL_508936 | | |
| Centre Hospitalier Pierre Oudot | CNR Virus des Infections Respiratoires - France SUD | Antonin Bal et al |
| EPI_ISL_508998, EPI_ISL_508966, EPI_ISL_508975 | | |
| Centre Hospitalier Régional Universitaire de Nantes Laboratoire de Virologie | National Reference Center for Viruses of Respiratory Infections, Institut Pasteur, Paris | Mélnie Albert et al |
| EPI_ISL_414625 | | |
| Centre Hospitalier René Dubois Laboratoire de Microbiologie - Bât A | National Reference Center for Viruses of Respiratory Infections, Institut Pasteur, Paris | Mélnie Albert et al |
| EPI_ISL_414633 | | |
| Centre Hospitalier Saint Joseph Saint Luc | CNR Virus des Infections Respiratoires - France SUD | Antonin Bal et al |
| EPI_ISL_508879, EPI_ISL_418418, EPI_ISL_418419, EPI_ISL_508880, EPI_ISL_420617, EPI_ISL_508960 | | |
| Centro de Desenvolvimento Tecnologico em Saude, Fundacao Oswaldo Cruz | Centro de Desenvolvimento Tecnologico em Saude, Fundacao Oswaldo Cruz | Souza et al |
| EPI_ISL_509430, EPI_ISL_509432, EPI_ISL_509433, EPI_ISL_510541, EPI_ISL_510536, EPI_ISL_509435 | | |
| Centro de Investigación Biomédica de La Rioja - Hospital San Pedro Logroño | SeqCOVID-SPAIN consortium/IBV(CSIC) | María de Toro et al |
| EPI_ISL_500430, EPI_ISL_500433, EPI_ISL_500440, EPI_ISL_537382 | | |
| Centro de Investigaciones, Universidad de Especialidades Espíritu Santo | Institute of Microbiology, Universidad San Francisco de Quito | Derly Andrade et al |
| EPI_ISL_491934 | | |
| Centro Hospital do Porto, E.P.E. - H. Geral de Santo Antonio | Instituto Nacional de Saude (INSA) | Guiomar et al et al |
| EPI_ISL_417999 | | |
| CENTRO ONCOLOGICO DEL NORTE | Instituto de Salud Publica de Chile | Andrés E Castillo et al |
| EPI_ISL_445267 | | |
| CH Compiègne Laboratoire de Biologie | National Reference Center for Viruses of Respiratory Infections, Institut Pasteur, Paris | Mélanie Albert et al |
| EPI_ISL_421500, EPI_ISL_420041, EPI_ISL_421510, EPI_ISL_421511, EPI_ISL_428353, EPI_ISL_428359, EPI_ISL_428360 | | |

| | | |
|---|---|---|
| CH Jean de Navarre Laboratoire de Biologie | National Reference Center for Viruses of Respiratory Infections, Institut Pasteur, Paris | Mélanie Albert et al |
| EPI_ISL_420053, EPI_ISL_428350 | | |
| CH Jean de Navarre Laboratoire de Biologie | National Reference Center for Viruses of Respiratory Infections, Institut Pasteur, Paris | Mélnie Albert et al |
| EPI_ISL_416493 | | |
| Charité Universitätsmedizin Berlin, Institut für Virologie/Labor Berlin | Charité Universitätsmedizin Berlin, Institut für Virologie/Labor Berlin | Victor M Corman et al |
| EPI_ISL_516633, EPI_ISL_516634, EPI_ISL_516630, EPI_ISL_516635, EPI_ISL_516637, EPI_ISL_516636, EPI_ISL_516632, EPI_ISL_516642, EPI_ISL_516638, EPI_ISL_516639, EPI_ISL_516629, EPI_ISL_516640, EPI_ISL_516643, EPI_ISL_516641, EPI_ISL_516644 | | |
| Charité Universitätsmedizin Berlin, Institute of Virology; Institut für Mikrobiologie der Bundeswehr, Munich | Charité Universitätsmedizin Berlin, Institute of Virology | Victor M Corman et al |
| EPI_ISL_406862 | | |
| Château de la Source | National Reference Center for Viruses of Respiratory Infections, Institut Pasteur, Paris | Mélanie Albert et al |
| EPI_ISL_443315 | | |
| Child Health Research Foundation | Child Health Research Foundation | Senjuti Saha et al |
| EPI_ISL_492028 | | |
| CHRU Bretonneau - Serv. Bacterio-Virol. | National Reference Center for Viruses of Respiratory Infections, Institut Pasteur, Paris | Mélanie Albert et al |
| EPI_ISL_418222 | | |
| CHRU Pontchaillou - Laboratoire de Virologie | National Reference Center for Viruses of Respiratory Infections, Institut Pasteur, Paris | Mélanie Albert et al |
| EPI_ISL_443289, EPI_ISL_443290, EPI_ISL_443291, EPI_ISL_443292, EPI_ISL_443293, EPI_ISL_443294 | | |
| CHRU Pontchaillou - Laboratoire de Virologie | National Reference Center for Viruses of Respiratory Infections, Institut Pasteur, Paris | Mélnie Albert et al |
| EPI_ISL_416504, EPI_ISL_416506, EPI_ISL_416507, EPI_ISL_416511 | | |
| CHU - Hôpital Cavale Blanche - Labo. de Virologie | National Reference Center for Viruses of Respiratory Infections, Institut Pasteur, Paris | Mélanie Albert et al |

| | | |
|---|---|---|
| EPI_ISL_443268, EPI_ISL_443270, EPI_ISL_443271, EPI_ISL_443274, EPI_ISL_443275, EPI_ISL_443277, EPI_ISL_443278, EPI_ISL_443279, EPI_ISL_443280, EPI_ISL_443281 | | |
| CHU Coimbra - Pediátrico | Instituto Nacional de Saude (INSA) | Guiomar et al et al |
| EPI_ISL_418005 | | |
| CHU de Dijon - Laboratoire de Virologie | National Reference Center for Viruses of Respiratory Infections, Institut Pasteur, Paris | Mélanie Albert et al |
| EPI_ISL_443261, EPI_ISL_443262 | | |
| CHU Gabriel Montpied | CNR Virus des Infections Respiratoires - France SUD | Bal et al |
| EPI_ISL_416752, EPI_ISL_416751 | | |
| CHU Purpan - Laboratoire de Virologie - Institut Fédératif de Biologie | Laboratoire de virologie - École Nationale Vétérinaire de Toulouse | Guillaume Croville et al |
| EPI_ISL_434626, EPI_ISL_434627, EPI_ISL_434628, EPI_ISL_434629, EPI_ISL_434630, EPI_ISL_434631, EPI_ISL_434632, EPI_ISL_434633, EPI_ISL_434634, EPI_ISL_434635, EPI_ISL_482879, EPI_ISL_482880, EPI_ISL_482881, EPI_ISL_482882, EPI_ISL_482883, EPI_ISL_482884, EPI_ISL_482885, EPI_ISL_482886, EPI_ISL_482887, EPI_ISL_482888, EPI_ISL_482889 | | |
| CHULC - H Curry Cabral | Instituto Nacional de Saude (INSA) | Guiomar et al et al |
| EPI_ISL_418013 | | |
| Civil Hospital, Panchkula | CSIR-Institute of Microbial Technology | Kanika Bansal et al |
| EPI_ISL_547588 | | |
| Clínica Universidad de Navarra. Servicio de Enfermedades Infecciosas y Microbiología clínica | SeqCOVID-SPAIN consortium/IBV(CSIC) | Mirian Fernández-Alonso et al |
| EPI_ISL_538101 | | |
| Clinical Microbiology Laboratory, Sheba Medical Center | Stern Lab | Stern Lab et al |
| EPI_ISL_447441 | | |
| Clinical Virology Laboratory, Soroka Medical Center and the Faculty of Health Sciences, Ben-Gurion University of the Negev | Stern Lab | Stern Lab et al |
| EPI_ISL_447328 | | |

| | | |
|---|---|---|
| Clinical Virology Unit, Hadassah Hebrew University Medical Center | Stern Lab | Stern Lab et al |
| EPI_ISL_447359 | | |
| CMIP | National Reference Center for Viruses of Respiratory Infections, Institut Pasteur, Paris | Mélanie Albert et al |
| EPI_ISL_420043 | | |
| CNR Virus des Infections Respiratoires - France SUD | CNR Virus des Infections Respiratoires - France SUD | Antonin Bal et al |
| EPI_ISL_525536, EPI_ISL_508945, EPI_ISL_508948, EPI_ISL_525537, EPI_ISL_508999, EPI_ISL_525541, EPI_ISL_525543, EPI_ISL_509002, EPI_ISL_508951, EPI_ISL_508953, EPI_ISL_508956, EPI_ISL_508961, EPI_ISL_508964, EPI_ISL_508965, EPI_ISL_508967, EPI_ISL_508969, EPI_ISL_508970, EPI_ISL_508972, EPI_ISL_508974, EPI_ISL_508977, EPI_ISL_508979, EPI_ISL_508980, EPI_ISL_508981, EPI_ISL_508982, EPI_ISL_508985, EPI_ISL_508988, EPI_ISL_508912, EPI_ISL_508913, EPI_ISL_508914, EPI_ISL_508990, EPI_ISL_508915, EPI_ISL_508992, EPI_ISL_508916, EPI_ISL_508993, EPI_ISL_508989, EPI_ISL_508994, EPI_ISL_508995, EPI_ISL_508997, EPI_ISL_508917, EPI_ISL_508918, EPI_ISL_508919, EPI_ISL_508922, EPI_ISL_508923, EPI_ISL_508924, EPI_ISL_508925, EPI_ISL_508926, EPI_ISL_508927, EPI_ISL_508928, EPI_ISL_508929 | | |
| CNR Virus des Infections Respiratoires - France SUD | CNR Virus des Infections Respiratoires - France SUD | Bal et al |
| EPI_ISL_416745 | | |
| Contra Costa Public Health Lab | Chan-Zuckerberg Biohub | CZB Cliahub Consortium et al |
| EPI_ISL_548579, EPI_ISL_548557, EPI_ISL_548556, EPI_ISL_548577 | | |
| CoronaNet Lab- TaskForce Regione Campania, CEINGE Biotecnologie Avanzate, Via G. Salvatore | CoronaNet Lab- TaskForce Regione Campania, CEINGE Biotecnologie Avanzate, Via G. Salvatore | Zollo et al |
| EPI_ISL_514751 | | |
| County of San Luis Obispo Public Health Laboratory | Chan-Zuckerberg Biohub | CZB Cliahub Consortium et al |
| EPI_ISL_548391 | | |
| County of Santa Clara Public Health Department | Chan-Zuckerberg Biohub | CZB Cliahub Consortium et al |

| | | |
|---|---|---|
| EPI_ISL_548342, EPI_ISL_548392, EPI_ISL_548395, EPI_ISL_548562, EPI_ISL_548568, EPI_ISL_548604, EPI_ISL_548584, EPI_ISL_548588, EPI_ISL_548661, EPI_ISL_548610, EPI_ISL_548659, EPI_ISL_548607 | | |
| CSIR-Centre for Cellular and Molecular Biology | CSIR-Centre for Cellular and Molecular Biology | Lamuk Zaveri et al |
| EPI_ISL_495243 | | |
| CSIR-Centre for Cellular and Molecular Biology | CSIR-Centre for Cellular and Molecular Biology | Nikhil Hajirnis et al |
| EPI_ISL_539655 | | |
| CSIR-Centre for Cellular and Molecular Biology | CSIR-Centre for Cellular and Molecular Biology | Pratheus a Maccha et al |
| EPI_ISL_528856 | | |
| CSIR-Centre for Cellular and Molecular Biology | CSIR-Centre for Cellular and Molecular Biology | Sakshi Shambha vi et al |
| EPI_ISL_539756 | | |
| CSIR-Centre for Cellular and Molecular Biology | CSIR-Centre for Cellular and Molecular Biology | Shagufta Khan et al |
| EPI_ISL_447562, EPI_ISL_447849 | | |
| CSIR-Centre for Cellular and Molecular Biology | CSIR-Centre for Cellular and Molecular Biology | Sofia Banu et al |
| EPI_ISL_447564 | | |
| CSIR-Centre for Cellular and Molecular Biology | CSIR-Centre for Cellular and Molecular Biology | Tulasi Nagaban di et al |
| EPI_ISL_450331 | | |
| CSSS de Port-Cartier | Laboratoire de santé publique du Québec | Sandrine Moreira et al |
| EPI_ISL_536109 | | |
| DB Diagnósticos do Brasil | Instituto de Medicina Tropical da Univesidade de São Paulo | Samples: Nelson Gaburo Jr et al |
| EPI_ISL_476224 | | |

| | | |
|---|---|---|
| deCODE genetics | deCODE genetics | Daniel F Gudbjartsson et al |
| EPI_ISL_417663 | | |
| Defence Research & Development Establishment (DRDE) | Defence Research & Development Establishment (DRDE) | Shashi Sharma et al |
| EPI_ISL_476896 | | |
| Departamento de Microbiología, CDB, Hospital Clínic, Barcelona | SeqCOVID-SPAIN consortium/IBV(CSIC) | Andrea Vergara et al |
| EPI_ISL_508645 | | |
| Department for Virology, Molecular Biology and Genome Research, R. G. Lugar Center for Public Health Research, National Center for Disease Control and Public Health (NCDC) of Georgia. | Department for Virology, Molecular Biology and Genome Research, R. G. Lugar Center for Public Health Research, National Center for Disease Control and Public Health (NCDC) of Georgia. | Gvantsa Chanturia et al |
| EPI_ISL_420144 | | |
| Department of Clinical Microbiology | GIGA Medical Genomics | Durkin Keith et al |
| EPI_ISL_417009 | | |
| Department of Clinical Microbiology | GIGA Medical Genomics | Keith Durkin et al |
| EPI_ISL_421185, EPI_ISL_421199, EPI_ISL_498145, EPI_ISL_498149, EPI_ISL_498150, EPI_ISL_498628, EPI_ISL_515056, EPI_ISL_515057, EPI_ISL_515060, EPI_ISL_515061, EPI_ISL_515064, EPI_ISL_515065, EPI_ISL_515066, EPI_ISL_515069, EPI_ISL_515070, EPI_ISL_515073, EPI_ISL_515074, EPI_ISL_515076, EPI_ISL_515079, EPI_ISL_515081, EPI_ISL_540471, EPI_ISL_540476, EPI_ISL_540477, EPI_ISL_540479, EPI_ISL_540484, EPI_ISL_540487, EPI_ISL_540488, EPI_ISL_540490, EPI_ISL_540492, EPI_ISL_540494, EPI_ISL_540495, EPI_ISL_540498, EPI_ISL_540500, EPI_ISL_540501, EPI_ISL_540504, EPI_ISL_540507, EPI_ISL_540509, EPI_ISL_540510, EPI_ISL_540512, EPI_ISL_540514, EPI_ISL_540515, EPI_ISL_540516, EPI_ISL_540517, EPI_ISL_540520, EPI_ISL_540521, EPI_ISL_540522, EPI_ISL_540523, EPI_ISL_540524, EPI_ISL_540525, EPI_ISL_540526, EPI_ISL_540528, EPI_ISL_540529, EPI_ISL_540531, EPI_ISL_540532, EPI_ISL_540533, EPI_ISL_540534, EPI_ISL_540535, EPI_ISL_540537, EPI_ISL_540538, EPI_ISL_540539, EPI_ISL_540541, EPI_ISL_540543, EPI_ISL_540545, EPI_ISL_540547, EPI_ISL_540548, EPI_ISL_540551, EPI_ISL_540553, EPI_ISL_540554, EPI_ISL_540555, EPI_ISL_540556, EPI_ISL_540557, EPI_ISL_540558, EPI_ISL_540559, EPI_ISL_540560, EPI_ISL_540561, EPI_ISL_540563, EPI_ISL_540564, EPI_ISL_540565, EPI_ISL_540566, EPI_ISL_540567, EPI_ISL_540568, EPI_ISL_540570, EPI_ISL_540571, EPI_ISL_540572, EPI_ISL_540573, | | |

| | | |
|---|---|---|
| EPI_ISL_540575, EPI_ISL_540576, EPI_ISL_540577, EPI_ISL_540578, EPI_ISL_447120, EPI_ISL_447121 | | |
| Department of Clinical Microbiology, Copenhagen University Hospital, Hvidovre, Kettegaard Alle 30, 2650 Hvidovre. | Albertsen lab, Department of Chemistry and Bioscience, Aalborg University, Denmark | Rasmus Kirkegaard et al |
| EPI_ISL_451994, EPI_ISL_452007, EPI_ISL_452024, EPI_ISL_429266, EPI_ISL_429295 | | |
| Department of Food Safety, Nutrition and Veterinary public health, Istituto Superiore di Sanita' | Department of Biomedical, Surgical and Dental Sciences and Department of Biomedical Sciences for Health | Delbue et al |
| EPI_ISL_487276 | | |
| Department of Infection Prevention and Infectious Diseases, University Hospital Regensburg | Department of Infection Prevention and Infectious Diseases, University Hospital Regensburg | Fritsch et al |
| EPI_ISL_522407, EPI_ISL_522408 | | |
| Department of Infection Prevention and Infectious Diseases, University Hospital Regensburg | University Hospital Regensburg | Fritsch et al |
| EPI_ISL_513307, EPI_ISL_513298, EPI_ISL_513299, EPI_ISL_513300, EPI_ISL_513301, EPI_ISL_513302, EPI_ISL_513303, EPI_ISL_513305, EPI_ISL_513306 | | |
| Department of Infectious Diseases, Cantonal Hospital Baden | Institute of Medical Virology, University of Zurich | Stefan Schmutz et al |
| EPI_ISL_524474, EPI_ISL_524480, EPI_ISL_524481 | | |
| Department of Infectious Diseases, Istituto Superiore di Sanità, Roma , Italy | Army Medical and Veterinary Research Center | Paola Stefanelli et al |
| EPI_ISL_457732, EPI_ISL_457724, EPI_ISL_457736, EPI_ISL_457721 | | |
| Department of Infectious Diseases, Istituto Superiore di Sanità, Roma , Italy | Virology Laboratory, Scientific Department, Army Medical Center | Paola Stefanelli et al |
| EPI_ISL_412973 | | |
| Department of Internal Medicine, Triemli Hospital | Institute of Medical Virology, University of Zurich | Stefan Schmutz et al |
| EPI_ISL_413019, EPI_ISL_413020 | | |
| Department of Laboratory Medicine, Tan Tock Seng Hospital | Department of Laboratory Medicine, Tan Tock Seng Hospital | Chen YYC et al |

| | | |
|---|---|---|
| EPI_ISL_476816, EPI_ISL_538460 | | |
| Department of Medical Microbiology, University Malaya Medical Centre | Department of Medical Microbiology, Faculty of Medicine, University of Malaya | Yoong Min CHONG et al |
| EPI_ISL_501193, EPI_ISL_501224, EPI_ISL_501226 | | |
| Department of Medical Microbiology, Western Sussex Hospitals NHS Foundation Trust, St Richard's Hospital | Wellcome Sanger Institute for the COVID-19 Genomics UK Consortium | Manasa Mutingwende et al |
| EPI_ISL_492910, EPI_ISL_492915, EPI_ISL_492886 | | |
| Department of Microbiology, The University of Hong Kong | Department of Microbiology, The University of Hong Kong | Kelvin K.W. To et al |
| EPI_ISL_497796, EPI_ISL_497802 | | |
| Department of Pathology, University of Cambridge | COVID-19 Genomics UK (COG-UK) Consortium | Luke W Meredith et al |
| EPI_ISL_447998, EPI_ISL_514451, EPI_ISL_425236, EPI_ISL_425250, EPI_ISL_433667, EPI_ISL_433693, EPI_ISL_425287, EPI_ISL_425314, EPI_ISL_425315, EPI_ISL_425318, EPI_ISL_425324, EPI_ISL_425327, EPI_ISL_425423, EPI_ISL_425433, EPI_ISL_433729, EPI_ISL_433730, EPI_ISL_433848, EPI_ISL_433486 | | |
| Department of Pathology, University of Cambridge | Wellcome Sanger Institute for the COVID-19 Genomics UK Consortium | Luke W Meredith et al |
| EPI_ISL_443656, EPI_ISL_470351, EPI_ISL_470141, EPI_ISL_489520, EPI_ISL_489476, EPI_ISL_489544, EPI_ISL_459472, EPI_ISL_439656, EPI_ISL_439545, EPI_ISL_440109, EPI_ISL_439997, EPI_ISL_440405, EPI_ISL_440473, EPI_ISL_438505, EPI_ISL_438437, EPI_ISL_439484, EPI_ISL_441143, EPI_ISL_441224, EPI_ISL_441303, EPI_ISL_441333, EPI_ISL_441664, EPI_ISL_441710, EPI_ISL_441592, EPI_ISL_470485, EPI_ISL_442870, EPI_ISL_443029, EPI_ISL_440490, EPI_ISL_440553 | | |
| Department of Virology | Department of Virology | Boehmer et al |
| EPI_ISL_450209, EPI_ISL_450200, EPI_ISL_450198 | | |
| Department of Virology and Immunology, University of Helsinki and Helsinki University Hospital, Huslab Finland | Department of Virology, Faculty of Medicine, University of Helsinki, Helsinki, Finland | Teemu Smura et al |
| EPI_ISL_418403 | | |

| | | |
|---|---|---|
| Department of Virus and Microbiological Special Diagnostics, Statens Serum Institut, Copenhagen, Denmark, Artillerivej 5, 2300 Copenahgen S | Albertsen lab, Department of Chemistry and Bioscience, Aalborg University, Denmark | Rasmus Kirkegaard et al |
| EPI_ISL_444821, EPI_ISL_437642, EPI_ISL_437647, EPI_ISL_437657, EPI_ISL_437000, EPI_ISL_444893, EPI_ISL_429334, EPI_ISL_429354, EPI_ISL_429371, EPI_ISL_429410, EPI_ISL_429433, EPI_ISL_429437, EPI_ISL_429440, EPI_ISL_429453, EPI_ISL_429498, EPI_ISL_429516 | | |
| Department of Virus and Microbiological Special diagnostics, Statens Serum Institut, Copenhagen, Denmark. | Statens Serum Institute | Morten Rasmussen et al |
| EPI_ISL_415647 | | |
| Department of Virus and Microbiological Special diagnostics, Statens Serum Institut, Copenhagen, Denmark. | ViFU | Morten Rasmussen et al |
| EPI_ISL_415646 | | |
| Diagnostic- and Research Institute of Pathology, Medical University of Graz | Diagnostic- and Research Institute of Pathology, Medical University of Graz | Karl Kashofer et al |
| EPI_ISL_437197, EPI_ISL_437300, EPI_ISL_437198, EPI_ISL_437201 | | |
| Division of Infectious Diseases and Hospital Epidemiology, University Hospital Zürich | Institute of Medical Virology, University of Zurich | Maryam Zaheri et al |
| EPI_ISL_508865, EPI_ISL_508867, EPI_ISL_508869, EPI_ISL_509222 | | |
| Division of Infectious Diseases and Hospital Epidemiology, University Hospital Zürich | Institute of Medical Virology, University of Zurich | Verena Kufner et al |
| EPI_ISL_508864, EPI_ISL_508866, EPI_ISL_508868, EPI_ISL_508870 | | |
| Division of Infectious Diseases, University Hospital Zurich | Institute of Medical Virology, University of Zurich | Stefan Schmutz et al |
| EPI_ISL_413022, EPI_ISL_413023, EPI_ISL_413024 | | |
| Division of Infectious Diseases, University Hospital Zürich | Institute of Medical Virology, University of Zurich | Stefan Schmutz et al |
| EPI_ISL_524475, EPI_ISL_524477 | | |

| | | |
|---|---|---|
| Division of Infectious Diseases, University Hospital Zürich | Institute of Medical Virology, University of Zurich | Verena Kufner et al |
| EPI_ISL_524483, EPI_ISL_524485 | | |
| Division of Viral Diseases, Center for Laboratory Control of Infectious Diseases, Korea Centers for Diseases Control and Prevention | Division of Viral Diseases, Center for Laboratory Control of Infectious Diseases, Korea Centers for Diseases Control and Prevention | Jeong-Min Kim et al |
| EPI_ISL_426182, EPI_ISL_471444, EPI_ISL_497985, EPI_ISL_498032, EPI_ISL_498044 | | |
| Dr. Georges-L.-Dumont University Hospital Centre | National Microbiology Laboratory | Anna Majer et al |
| EPI_ISL_429806, EPI_ISL_429811 | | |
| Dutch COVID-19 response team | Erasmus Medical Center | Bas Oude Munnink et al |
| EPI_ISL_523121, EPI_ISL_523137, EPI_ISL_455115, EPI_ISL_523273, EPI_ISL_523275, EPI_ISL_523277, EPI_ISL_523280, EPI_ISL_460657, EPI_ISL_422641, EPI_ISL_461039, EPI_ISL_422684, EPI_ISL_422685, EPI_ISL_422693, EPI_ISL_422701, EPI_ISL_422720, EPI_ISL_422742, EPI_ISL_422762, EPI_ISL_422763, EPI_ISL_422785, EPI_ISL_455129, EPI_ISL_455133, EPI_ISL_455134, EPI_ISL_455167, EPI_ISL_455170, EPI_ISL_460759, EPI_ISL_461106, EPI_ISL_422843, EPI_ISL_422850, EPI_ISL_523343, EPI_ISL_523381, EPI_ISL_523416, EPI_ISL_461151, EPI_ISL_461152, EPI_ISL_461164, EPI_ISL_422868, EPI_ISL_422876, EPI_ISL_422889, EPI_ISL_523534, EPI_ISL_523535, EPI_ISL_523537, EPI_ISL_523540, EPI_ISL_523541, EPI_ISL_523542, EPI_ISL_523543, EPI_ISL_523546, EPI_ISL_422898, EPI_ISL_523599, EPI_ISL_523606, EPI_ISL_523611, EPI_ISL_523641, EPI_ISL_523643, EPI_ISL_523644, EPI_ISL_523646, EPI_ISL_523663, EPI_ISL_523687, EPI_ISL_523697, EPI_ISL_523698, EPI_ISL_523699, EPI_ISL_523701, EPI_ISL_523702, EPI_ISL_523706, EPI_ISL_523707, EPI_ISL_523709, EPI_ISL_523711, EPI_ISL_523712, EPI_ISL_523713, EPI_ISL_523716, EPI_ISL_523718, EPI_ISL_523721, EPI_ISL_523722, EPI_ISL_523723, EPI_ISL_523724, EPI_ISL_523725, EPI_ISL_523729, EPI_ISL_523730, EPI_ISL_523738, EPI_ISL_523740, EPI_ISL_523741, EPI_ISL_523743, EPI_ISL_523747, EPI_ISL_523748, EPI_ISL_523749, EPI_ISL_523751, EPI_ISL_523753, EPI_ISL_523754, EPI_ISL_523756, EPI_ISL_523757, EPI_ISL_523758, EPI_ISL_523760, EPI_ISL_523763, EPI_ISL_523764, EPI_ISL_523768, EPI_ISL_523769, EPI_ISL_523772, EPI_ISL_523773, EPI_ISL_523774, EPI_ISL_523775, EPI_ISL_523777, EPI_ISL_523779, EPI_ISL_523780, EPI_ISL_523785, EPI_ISL_523787, EPI_ISL_523790, EPI_ISL_523792, EPI_ISL_523793, EPI_ISL_523794, EPI_ISL_523795, EPI_ISL_523796, EPI_ISL_523804, EPI_ISL_523805, EPI_ISL_523806, EPI_ISL_461260, EPI_ISL_422901, EPI_ISL_461316, EPI_ISL_422918, EPI_ISL_422924, EPI_ISL_422941, EPI_ISL_523471, EPI_ISL_523512, EPI_ISL_523515 | | |

| | | |
|---|---|---|
| Dutch COVID-19 response team | Erasmus Medical Center | David Nieuwenhuijse et al |
| EPI_ISL_415468, EPI_ISL_415487, EPI_ISL_414536, EPI_ISL_415511, EPI_ISL_414442, EPI_ISL_414554, EPI_ISL_414551, EPI_ISL_414562, EPI_ISL_414564, EPI_ISL_415530, EPI_ISL_415535 | | |
| Dutch COVID-19 response team | National Institute for Public Health and the Environment (RIVM) | Adam Meijer et al |
| EPI_ISL_547461, EPI_ISL_454789, EPI_ISL_454794, EPI_ISL_454750, EPI_ISL_454755, EPI_ISL_547513, EPI_ISL_454767, EPI_ISL_454771, EPI_ISL_454785 | | |
| EHPAD - Résidences les Cèdres | National Reference Center for Viruses of Respiratory Infections, Institut Pasteur, Paris | Mélanie Albert et al |
| EPI_ISL_418226 | | |
| Emergency county Hospital Suceava | Stefan cel Mare University Metagenomics Lab | Lobiuc Andrei et al. et al |
| EPI_ISL_486855 | | |
| Federal Budget Institution of Science, State Research Center for Applied Microbiology & Biotechnology | Federal Budget Institution of Science, State Research Center for Applied Microbiology & Biotechnology | Dyatlov I et al |
| EPI_ISL_451970 | | |
| Félix Guyon Hospital | UMR PIMIT Université de La Réunion | David Wilkinson et al |
| EPI_ISL_522550 | | |
| FL Bureau of Public Health Laboratories-Tampa | Pathogen Discovery, Respiratory Viruses Branch, Division of Viral Diseases, Centers for Disease Control and Prevention | Anna Uehara et al |
| EPI_ISL_419559 | | |
| Florida Bureau of Public Health Laboratories | Florida Bureau of Public Health Laboratories | Sarah Schmedes et al |
| EPI_ISL_489764, EPI_ISL_509784, EPI_ISL_509785, EPI_ISL_509794, EPI_ISL_509796, EPI_ISL_512572, EPI_ISL_514202, EPI_ISL_517816, EPI_ISL_517850, EPI_ISL_517902, EPI_ISL_517914, EPI_ISL_517928, EPI_ISL_517930, EPI_ISL_517948, EPI_ISL_517950, EPI_ISL_526585, EPI_ISL_526597, EPI_ISL_526598, EPI_ISL_526610, EPI_ISL_526612, EPI_ISL_526619, EPI_ISL_526621, EPI_ISL_526636, EPI_ISL_526652, EPI_ISL_526653, EPI_ISL_526659, EPI_ISL_526665, EPI_ISL_526675, EPI_ISL_526685, EPI_ISL_568642 | | |

| | | |
|---|---|---|
| Florida Bureau of Public Health Laboratories, Florida Department of Health | Florida Bureau of Public Health Laboratories, Florida Department of Health | Schmedes et al |
| EPI_ISL_541197, EPI_ISL_541168, EPI_ISL_541263, EPI_ISL_541312 | | |
| FSBSI "Chumakov Federal Scientific Center for Research and Development of Immune-and-Biological Products of Russian Academy of Sciences" | FSBSI "Chumakov Federal Scientific Center for Research and Development of Immune-and-Biological Products of Russian Academy of Sciences" & NRC "Kurchatov institute" | Liubov Kozlovskaya et al |
| EPI_ISL_428851 | | |
| Fujian Center for Disease Control and Prevention | Fujian Center for Disease Control and Prevention | Lin Qi et al |
| EPI_ISL_431779 | | |
| Fukui Prefectural Institute of Public Health and Environmental Science | Pathogen Genomics Center, National Institute of Infectious Diseases | Tsuyoshi Sekizuka et al |
| EPI_ISL_479970, EPI_ISL_480128 | | |
| Furst Medical Laboratory | Norwegian Institute of Public Health, Department of Virology | Kathrine Stene-Johansen et al |
| EPI_ISL_549168 | | |
| General Hospital - Kumanovo | Research Center for Genetic Engineering and Biotechnology "Georgi D. Efremov" , Macedonian Academy of Sciences and Arts | RCGEB - MASA et al |
| EPI_ISL_516413 | | |
| General Hospital of Central Theater Command of People's Liberation Army of China | BGI & Institute of Microbiology, Chinese Academy of Sciences & Shandong First Medical University & Shandong Academy of Medical Sciences & General Hospital of Central Theater Command of People's Liberation Army of China | Weijun Chen et al |
| EPI_ISL_406798 | | |
| General Intensive Care Unit, Raymond Poincaré Hospital (AP-HP), Lab Inflammation & Infection, U1173 University Paris Saclay-UVSQ/INSERM, Garches, France. | Institut Pasteur, Laboratory for Urgent Response to biological Threats | Annane Djillali et al |
| EPI_ISL_430846 | | |

| | | |
|---|---|---|
| Genomic Laboratory (GLAB) (Conjoint lab of Health Directorate of Istanbul and Istanbul Technical University) | Genomic Laboratory (GLAB), Istanbul Technical University | Ilker Karacan et al |
| EPI_ISL_480244 | | |
| GH Nord Essonne Service de Biologie clinique | National Reference Center for Viruses of Respiratory Infections, Institut Pasteur, Paris | Mélanie Albert et al |
| EPI_ISL_428351, EPI_ISL_428363 | | |
| Gorgas Memorial Laboratory of Health Studies | Gorgas Memorial Laboratory of Health Studies | Danilo Franco et al |
| EPI_ISL_496614 | | |
| Government General Hospital, Jam Khambhaliya, Devbhoomi Dwarka | Gujarat Biotechnology Research Centre | Harish Matani et al |
| EPI_ISL_524735 | | |
| Government Medical College, Bhavnagar | Gujarat Biotechnology Research Centre | Dinesh Kumar et al |
| EPI_ISL_495025 | | |
| Grupo de Investigaciones Microbiológicas-UR (GIMUR), Departamento de Biología, Facultad de Ciencias Naturales, Universidad del Rosario, Bogotá, Colombia | Grupo de Investigaciones Microbiológicas-UR (GIMUR), Departamento de Biología, Facultad de Ciencias Naturales, Universidad del Rosario, Bogotá, Colombia Instituto Nacional de Salud, Bogotá, Colombia Icahn School of Medicine at Mount Sinai, New York, USA | Juan David Ramírez et al |
| EPI_ISL_447751 | | |
| Guangdong Provincial Center for Diseases Control and Prevention;Guangdong Provincial Institute of Public Health | School of Public Health, The University of Hong Kong | Bosheng Li et al |
| EPI_ISL_428449 | | |
| Gundersen Clinical Microbiology Laboratory | Kabara Cancer Research Institute | Craig S. Richmond et al |
| EPI_ISL_547672 | | |
| Gundersen Molecular Diagnostics Laboratory | Kabara Cancer Research Institute | Craig S. Richmond et al |

| | | |
|---|---|---|
| EPI_ISL_489953, EPI_ISL_547615, EPI_ISL_547630, EPI_ISL_547641, EPI_ISL_547687, EPI_ISL_547689, EPI_ISL_547695, EPI_ISL_547702, EPI_ISL_547730, EPI_ISL_547734, EPI_ISL_547746 | | |
| H Dr Nelio Mendonca - Funchal | Instituto Nacional de Saude (INSA) | Guiomar et al et al |
| EPI_ISL_421457 | | |
| H Dr. Nelio Mendonca - Funchal | Instituto Nacional de Saude (INSA) | Guiomar et al et al |
| EPI_ISL_421461 | | |
| Halmstad klinisk mikrobiologi | The Public Health Agency of Sweden | Anna-Malin Linde et al |
| EPI_ISL_454445 | | |
| Halmstad klinisk mikrobiologi | The Public Health Agency of Sweden | Oskar Karlsson Lindsjo et al |
| EPI_ISL_469073 | | |
| Hangzhou Center for Disease Control and Prevention | Insepction Center of Hangzhou Center for Disease Control and Prevention | Yu hua et al |
| EPI_ISL_418510 | | |
| Hangzhou Center for Diseases Control and Prevention | Hangzhou Center for Diseases Control and Prevention | Jun Li et al |
| EPI_ISL_482581, EPI_ISL_482585 | | |
| Hematology Laboratory, Section of Molecular Diagnostics, University Clinical Centre, Medical University of Gdansk | Department of Virology, Faculty of Medicine, University of Helsinki, Helsinki, Finland | Maciej Grzybek et al |
| EPI_ISL_512300, EPI_ISL_512301 | | |
| Hong Kong Department of Health | School of Public Health, The University of Hon g Kong | Dominic N.C. Tsang et al |
| EPI_ISL_412028 | | |
| hopital | National Reference Center for Viruses of Respiratory Infections, Institut Pasteur, Paris | Sylvie Behillil et al |
| EPI_ISL_560591, EPI_ISL_560595, EPI_ISL_560596, EPI_ISL_560597 | | |

| | | |
|---|---|---|
| Hopital | National Reference Center for Viruses of Respiratory Infections, Institut Pasteur, Paris | Sylvie Behillil et al |
| EPI_ISL_560581, EPI_ISL_560583, EPI_ISL_560584, EPI_ISL_560585, EPI_ISL_560586 | | |
| hôpital | National Reference Center for Viruses of Respiratory Infections, Institut Pasteur, Paris | Sylvie Behillil et al |
| EPI_ISL_560568, EPI_ISL_560569, EPI_ISL_560571, EPI_ISL_560572, EPI_ISL_560574, EPI_ISL_560575, EPI_ISL_560576, EPI_ISL_560578, EPI_ISL_560579, EPI_ISL_560580 | | |
| Hôpital Charles-LeMoyne | Laboratoire de santé publique du Québec | Sandrine Moreira et al |
| EPI_ISL_535874, EPI_ISL_535972, EPI_ISL_535973, EPI_ISL_536280, EPI_ISL_536281, EPI_ISL_536284, EPI_ISL_536350, EPI_ISL_450310 | | |
| Hôpital de Hull | Laboratoire de santé publique du Québec | Sandrine Moreira et al |
| EPI_ISL_536132, EPI_ISL_536367 | | |
| Hôpital de Saint-Eustache | Laboratoire de santé publique du Québec | Sandrine Moreira et al |
| EPI_ISL_536008 | | |
| Hôpital du Centre-de-la-Mauricie | Laboratoire de santé publique du Québec | Sandrine Moreira et al |
| EPI_ISL_536246, EPI_ISL_536251 | | |
| Hôpital du Suroît | Laboratoire de santé publique du Québec | Sandrine Moreira et al |
| EPI_ISL_536227 | | |
| Hôpital et Centre d'Hébergement de Sept-Iles | Laboratoire de santé publique du Québec | Sandrine Moreira et al |
| EPI_ISL_535991, EPI_ISL_536114 | | |
| Hopital franco britannique - Service des Urgences | National Reference Center for Viruses of Respiratory Infections, Institut Pasteur, Paris | Mélnie Albert et al |
| EPI_ISL_416501 | | |

| | | |
|---|---|---|
| Hôpital Necker - Enfants - Malades Laboratoire de Virologie | National Reference Center for Viruses of Respiratory Infections, Institut Pasteur, Paris | Mélanie Albert et al |
| EPI_ISL_443295, EPI_ISL_443296, EPI_ISL_443298 | | |
| Hôpital Pierre-Boucher | Laboratoire de santé publique du Québec | Sandrine Moreira et al |
| EPI_ISL_536039, EPI_ISL_536096, EPI_ISL_536305, EPI_ISL_536347 | | |
| Hopital Privé de l'Est Lyonnais | CNR Virus des Infections Respiratoires - France SUD | Antonin Bal et al |
| EPI_ISL_418427 | | |
| Hôpital régional de Saint-Jérôme | Laboratoire de santé publique du Québec | Sandrine Moreira et al |
| EPI_ISL_535791 | | |
| Hôpital Robert Debré Laboratoire de Virologie | National Reference Center for Viruses of Respiratory Infections, Institut Pasteur, Paris | Mélnie Albert et al |
| EPI_ISL_414631 | | |
| Hospital | National Reference Center for Viruses of Respiratory Infections, Institut Pasteur, Paris | Sylvie Behillil et al |
| EPI_ISL_560598, EPI_ISL_560599, EPI_ISL_560600, EPI_ISL_560601, EPI_ISL_560602, EPI_ISL_560603, EPI_ISL_560604, EPI_ISL_560605, EPI_ISL_560606, EPI_ISL_560607, EPI_ISL_560608, EPI_ISL_560611, EPI_ISL_560612, EPI_ISL_560613, EPI_ISL_560614, EPI_ISL_560615, EPI_ISL_560616, EPI_ISL_560618, EPI_ISL_560619, EPI_ISL_560620, EPI_ISL_560621, EPI_ISL_560622, EPI_ISL_560623, EPI_ISL_560624, EPI_ISL_560628, EPI_ISL_560629, EPI_ISL_560630, EPI_ISL_560631, EPI_ISL_560633, EPI_ISL_560634, EPI_ISL_560635, EPI_ISL_560636, EPI_ISL_560643, EPI_ISL_560645, EPI_ISL_560646 | | |
| Hospital AZ Rivierenland | Institute of Tropical Medicine | Philippe Selhorst et al |
| EPI_ISL_450726 | | |
| Hospital Clínica Bíblica | Charité Virology-University of Costa Rica | Andres Moreira-Soto et al |
| EPI_ISL_480316, EPI_ISL_480319 | | |
| Hospital Clínico Universitario de Santiago de Compostela | SeqCOVID-SPAIN consortium/IBV(CSIC) | José Javier Costa |

| | | |
|---|---|---|
| | | Alcalde et al |
| EPI_ISL_510283, EPI_ISL_510293, EPI_ISL_510301 | | |
| Hospital e Maternidade Nossa Senhora das Graças | Instituto Adolfo Lutz, Interdiciplinary Procedures Center, Strategic Laboratory | Claudio Tavares Sacchi et al |
| EPI_ISL_547577 | | |
| Hospital General Universitario Gregorio Marañón | SeqCOVID-SPAIN consortium/IBV(CSIC) | Laura Pérez-Lago et al |
| EPI_ISL_510204, EPI_ISL_510222, EPI_ISL_541888, EPI_ISL_541890, EPI_ISL_541891, EPI_ISL_541895, EPI_ISL_541901, EPI_ISL_541905, EPI_ISL_541906, EPI_ISL_541907, EPI_ISL_541908, EPI_ISL_541911, EPI_ISL_541913, EPI_ISL_541914, EPI_ISL_541916, EPI_ISL_541917, EPI_ISL_541919, EPI_ISL_541920, EPI_ISL_541922, EPI_ISL_541926, EPI_ISL_541927, EPI_ISL_541928, EPI_ISL_541929, EPI_ISL_541930, EPI_ISL_541932, EPI_ISL_541933, EPI_ISL_541934, EPI_ISL_541935, EPI_ISL_541936, EPI_ISL_541937, EPI_ISL_541940 | | |
| HOSPITAL HERMINDA MARTIN CHILLAN | Instituto de Salud Publica de Chile | Andrés E Castillo et al |
| EPI_ISL_445331 | | |
| Hospital Israelita Albert Einstein | Instituto Adolfo Lutz, Interdiciplinary Procedures Center, Strategic Laboratory | Claudio Tavares Sacchi et al |
| EPI_ISL_416034 | | |
| HOSPITAL REG.LAUTARO NAVARRO AVARIA | Instituto de Salud Publica de Chile | Andrés E Castillo et al |
| EPI_ISL_445268 | | |
| Hospital San Pedro de Alcántara (Cáceres) | SeqCOVID-SPAIN consortium/IBV(CSIC) | Cristina Muñoz Cuevas et al |
| EPI_ISL_510317 | | |
| Hospital Schwyz | Institute of Medical Virology, University of Zurich | Verena Kufner et al |
| EPI_ISL_524484 | | |

| | | |
|---|---|---|
| Hospital Universitario 12 de Octubre | Hospital Universitario 12 de Octubre | Esther Viedma et al |
| EPI_ISL_428709 | | |
| Hospital Universitario 12 de Octubre | Hospital Universitario 12 de Octubre | Raúl Recio et al |
| EPI_ISL_428696, EPI_ISL_534328 | | |
| Hospital Universitario 12 de Octubre | Hospital Universitario La Paz | Elias Dahdouh et al |
| EPI_ISL_417961 | | |
| Hospital Universitario Araba. Vitoria-Gasteiz | SeqCOVID-SPAIN consortium/IBV(CSIC) | Silvia Hernáez Crespo et al |
| EPI_ISL_500232 | | |
| Hospital Universitario Araba. Vitoria-Gasteiz, | SeqCOVID-SPAIN consortium/IBV(CSIC) | Silvia Hernáez Crespo et al |
| EPI_ISL_452706 | | |
| Hospital Universitario de Gran Canaria Dr. Negrín | SeqCOVID-SPAIN consortium/IBV(CSIC) | M. Carmen Pérez González et al |
| EPI_ISL_561370, EPI_ISL_537707, EPI_ISL_537795, EPI_ISL_537801 | | |
| Hospital Universitario de La Ribera (Alzira, València) | SeqCOVID-SPAIN consortium/IBV(CSIC) | Olalla Martínez Macias et al |
| EPI_ISL_539283 | | |
| Hospital Universitario La Paz | Hospital Universitario La Paz | María Rodrígue z et al |
| EPI_ISL_530034 | | |
| Hospital Universitario Virgen de las Nieves de Granada-SAS | SeqCOVID-SPAIN consortium/IBV(CSIC) | Mercede s Pérez Ruiz et al |

| | | |
|---|---|---|
| EPI_ISL_474929, EPI_ISL_510424, EPI_ISL_510440, EPI_ISL_510445, EPI_ISL_510452, EPI_ISL_510456, EPI_ISL_474920 | | |
| Houston Methodist Hospital | Houston Methodist Hospital | S. Wesley Long et al |
| EPI_ISL_542516, EPI_ISL_543882, EPI_ISL_543999, EPI_ISL_544116, EPI_ISL_544182, EPI_ISL_543638, EPI_ISL_544476, EPI_ISL_544531, EPI_ISL_547015, EPI_ISL_547213, EPI_ISL_547236, EPI_ISL_547237, EPI_ISL_547243, EPI_ISL_547252, EPI_ISL_547253, EPI_ISL_547306, EPI_ISL_547369, EPI_ISL_547397, EPI_ISL_547420, EPI_ISL_545962, EPI_ISL_545963, EPI_ISL_545986, EPI_ISL_546079, EPI_ISL_546654, EPI_ISL_545608, EPI_ISL_545741, EPI_ISL_545766, EPI_ISL_545771, EPI_ISL_545802, EPI_ISL_434845, EPI_ISL_434899, EPI_ISL_434801 | | |
| Humboldt County Public Health Laboratory | Chan-Zuckerberg Biohub | CZB Cliahub Consortium et al |
| EPI_ISL_454637 | | |
| ICMR-National Institute of Cholera and Enteric Diseases | National Institute of Biomedical Genomics | Arindam Maitra et al |
| EPI_ISL_455672 | | |
| Immunogenomics lab, Institute of Life Sciences, Bhubaneswar | Immunogenomics lab, Institute of Life Sciences, Bhubaneswar | Sunil Raghav et al |
| EPI_ISL_481118, EPI_ISL_481123 | | |
| INMI Lazzaro Spallanzani IRCCS | INMI Lazzaro Spallanzani IRCCS | Antonino Di Caro et al |
| EPI_ISL_419255 | | |
| INMI Lazzaro Spallanzani IRCCS | INMI Lazzaro Spallanzani IRCCS | Barbara Bartolini et al |
| EPI_ISL_493329 | | |
| INMI Lazzaro Spallanzani IRCCS | INMI Lazzaro Spallanzani IRCCS | Cesare E.M. Gruber et al |
| EPI_ISL_493330 | | |
| INMI Lazzaro Spallanzani IRCCS | INMI Lazzaro Spallanzani IRCCS | Martina Rueca et al |
| EPI_ISL_493328, EPI_ISL_493331 | | |

| | | |
|---|---|---|
| INMI Lazzaro Spallanzani IRCCS | Laboratory of Virology, INMI Lazzaro Spallanzani IRCCS | Barbara Bartolini et al |
| EPI_ISL_419254 | | |
| INMI Lazzaro Spallanzani IRCCS | Laboratory of Virology, INMI Lazzaro Spallanzani IRCCS | Concetta Castilletti et al |
| EPI_ISL_424342 | | |
| INMI Lazzaro Spallanzani IRCCS | Laboratory of Virology, INMI Lazzaro Spallanzani IRCCS | Eleonora Lalle et al |
| EPI_ISL_424344 | | |
| INMI Lazzaro Spallanzani IRCCS | Laboratory of Virology, INMI Lazzaro Spallanzani IRCCS | Fabrizio Carletti et al |
| EPI_ISL_424343 | | |
| Innovative Genomics Institute, UC Berkeley | Innovative Genomics Institute, UC Berkeley | Stacia Wyman et al |
| EPI_ISL_569889, EPI_ISL_569901, EPI_ISL_569907 | | |
| Institut des Agents Infectieux (IAI) Hospices Civils de Lyon | CNR Virus des Infections Respiratoires - France SUD | Bal et al |
| EPI_ISL_416750 | | |
| Institut des Agents Infectieux (IAI), Hospices Civils de Lyon | CNR Virus des Infections Respiratoires - France SUD | Antonin Bal et al |
| EPI_ISL_509010, EPI_ISL_508872, EPI_ISL_508874, EPI_ISL_508877, EPI_ISL_418420, EPI_ISL_418423, EPI_ISL_508882, EPI_ISL_508883, EPI_ISL_418430, EPI_ISL_418431, EPI_ISL_419169, EPI_ISL_419171, EPI_ISL_419172, EPI_ISL_419173, EPI_ISL_419180, EPI_ISL_420604, EPI_ISL_420606, EPI_ISL_420607, EPI_ISL_420608, EPI_ISL_420615, EPI_ISL_420616, EPI_ISL_420621, EPI_ISL_420622, EPI_ISL_420625, EPI_ISL_508884, EPI_ISL_508885, EPI_ISL_508886, EPI_ISL_508887, EPI_ISL_508888, EPI_ISL_508889, EPI_ISL_508890, EPI_ISL_508891, EPI_ISL_508893, EPI_ISL_508894, EPI_ISL_508895, EPI_ISL_508896, EPI_ISL_508900, EPI_ISL_508901, EPI_ISL_508902, EPI_ISL_508903, EPI_ISL_508907, EPI_ISL_508908, EPI_ISL_509009, EPI_ISL_509011, EPI_ISL_509013, EPI_ISL_509014, EPI_ISL_417333, EPI_ISL_417334, EPI_ISL_417336, EPI_ISL_417339, EPI_ISL_508909, EPI_ISL_508910, EPI_ISL_508911 | | |
| Institut für Virologie am Department für Hygiene, Mikrobiologie und Public Health | Bergthaler laboratory, CeMM Research Center for Molecular Medicine of the Austrian Academy of Sciences | Alexandra Popa et al |
| EPI_ISL_437915, EPI_ISL_437921, EPI_ISL_437922, EPI_ISL_437924, EPI_ISL_437974, EPI_ISL_437978, EPI_ISL_437981, EPI_ISL_437988, EPI_ISL_437990, EPI_ISL_475815, EPI_ISL_475817, EPI_ISL_475820 | | |

| | | |
|---|---|---|
| Institut für Virologie und Epidemiologie der Viruskrankheiten, Universitätsklinikum Tübingen | NGS Competence Center Tübingen, Institut für Medizinische Mikrobiologie und Hygiene, Universitätsklinikum Tübingen | Angel Angelov et al |
| EPI_ISL_508688, EPI_ISL_508689, EPI_ISL_508690, EPI_ISL_508692, EPI_ISL_508695, EPI_ISL_508696, EPI_ISL_508699 | | |
| Institut für Virologie und Epidemiologie der Viruskrankheiten, Universitätsklinikum Tübingen | NGS Competence Center Tübingen, Institut für Medizinische Mikrobiologie und Hygiene, Universitätsklinikum Tübingen | Angelov at al. et al |
| EPI_ISL_485809 | | |
| Institut für Virologie und Epidemiologie der Viruskrankheiten, Universitätsklinikum Tübingen | NGS Competence Center Tübingen, Institut für Medizinische Mikrobiologie und Hygiene, Universitätsklinikum Tübingen | Angelov et al. et al |
| EPI_ISL_485810, EPI_ISL_485811, EPI_ISL_485812, EPI_ISL_485813 | | |
| Institut Médico légal- Hop R. Poincaré | National Reference Center for Viruses of Respiratory Infections, Institut Pasteur, Paris | Mélanie Albert et al |
| EPI_ISL_428355, EPI_ISL_428357 | | |
| Institut Pasteur du Maroc | Institut Pasteur du Maroc | Marion Barbet et al |
| EPI_ISL_459977 | | |
| Institute for Medical Research, Infectious Disease Research Centre, National Institutes of Health, Ministry of Health Malaysia | Institute for Medical Research, Infectious Disease Research Centre, National Institutes of Health, Ministry of Health Malaysia | Suppiah.J et al |
| EPI_ISL_430440 | | |
| Institute of Clinical Microbiology and Hygiene, University Hospital Regensburg | Institute of Clinical Microbiology and Hygiene, University Hospital Regensburg | Hiergeist et al |
| EPI_ISL_525472, EPI_ISL_525473 | | |
| Institute of Disease Control and Prevention, People's Liberation Army | Institute of Disease Control and Prevention, People's Liberation Army | Qiu et al |
| EPI_ISL_539338 | | |
| Institute of Human Genetics, Polish Academy of Sciences | Institute of Human Genetics, Polish Academy of Sciences | Szymon Hryhorowicz et al |
| EPI_ISL_485399 | | |

| | | |
|---|---|---|
| Institute of Molecular Virology, University Münster | Institute of Molecular Virology, University Münster | Angeles Mecate Zambrano et al |
| EPI_ISL_463008 | | |
| Instituto Gulbenkian de Ciência | Instituto Gulbenkian de Ciência | Cathy Paulino et al |
| EPI_ISL_491224, EPI_ISL_491226 | | |
| Instituto Gulbenkian de Ciência | Instituto Gulbenkian de Ciência | João Costa et al |
| EPI_ISL_491189, EPI_ISL_491192, EPI_ISL_491197 | | |
| Instituto Gulbenkian de Ciência | Instituto Gulbenkian de Ciência | Joao Sobral et al |
| EPI_ISL_491231, EPI_ISL_491237, EPI_ISL_491239, EPI_ISL_491242, EPI_ISL_491250, EPI_ISL_491259 | | |
| Instituto Gulbenkian de Ciência | Instituto Gulbenkian de Ciência | Susana Ladeiro et al |
| EPI_ISL_491291, EPI_ISL_491292 | | |
| Instituto Nacional de Investigación en Salud Pública - INSPI | INSPI - Charité | Alfredo Bruno Caicedo et al |
| EPI_ISL_491944 | | |
| Instituto Nacional de Salud | Laboratorio de Infecciones Respiratorias Agudas | Eduardo Juscamayta Lopez et al |
| EPI_ISL_536536 | | |
| Instituto Nacional de Salud, Bogotá, Colombia | Grupo de Investigaciones Microbiológicas-UR (GIMUR), Departamento de Biología, Facultad de Ciencias Naturales, Universidad del Rosario, Bogotá, Colombia Instituto Nacional de Salud, Bogotá, Colombia Icahn School of Medicine at Mount Sinai, New York, USA | Juan David Ramírez et al |
| EPI_ISL_447784 | | |

| | | |
|---|---|---|
| Instituto Nacional de Salud, Bogotá, Colombia | Instituto Nacional de Salud, Bogotá, Colombia | Katherine Laiton-Donato et al |
| EPI_ISL_498158, EPI_ISL_498162 | | |
| Instituto Nacional de Saude (INSA) | Instituto Nacional de Saude (INSA) | Borges et al et al |
| EPI_ISL_511030, EPI_ISL_511085, EPI_ISL_511579, EPI_ISL_511396 | | |
| Instituto Nacional de Saude (INSA) | Instituto Nacional de Saude (INSA) and Instituto Gulbenkian de Ciencia (IGC) | Borges et al et al |
| EPI_ISL_511493, EPI_ISL_511494 | | |
| Instituto Nacional de Saude (INSA) and Instituto Gulbenkian de Ciencia (IGC) | Instituto Nacional de Saude (INSA) and Instituto Gulbenkian de Ciencia (IGC) | Borges et al et al |
| EPI_ISL_511200, EPI_ISL_511219 | | |
| Instituto Sabin | Laboratory of Virology | Fernando L Melo et al |
| EPI_ISL_426580 | | |
| IRCCS Sacro Cuore Don Calabria Hospital, Department of Infectious, Tropical Diseases & Microbiology | University of Verona, Department of Biotechnology | Antonio Mori et al |
| EPI_ISL_492981, EPI_ISL_492982, EPI_ISL_492983, EPI_ISL_492984, EPI_ISL_492986, EPI_ISL_492987 | | |
| Israel Central Virology laboratory | Israel Central Virology laboratory | Neta Zuckerman et al |
| EPI_ISL_474961, EPI_ISL_475011, EPI_ISL_514268, EPI_ISL_514276, EPI_ISL_514288, EPI_ISL_514289, EPI_ISL_514291, EPI_ISL_514295, EPI_ISL_516905 | | |
| Istituto Zooprofilattico Sperimentale del Mezzogiorno | INMI Lazzaro Spallanzani IRCCS | Barbara Bartolini et al |
| EPI_ISL_493333, EPI_ISL_560407 | | |
| Istituto Zooprofilattico Sperimentale del Mezzogiorno | INMI Lazzaro Spallanzani IRCCS | Cesare E.M. Gruber et al |
| EPI_ISL_493332 | | |
| Istituto Zooprofilattico Sperimentale Puglia e Basilicata; | Beaconlab (Bioinformatics, Evolution and Comparative Genomics lab), Dept of Biosciences, University on Mila | Parisi A. et al |

| | | |
|---|---|---|
| EPI_ISL_477194, EPI_ISL_477203, EPI_ISL_477202 | | |
| Istituto Zooprofilattico Sperimentale Puglia e Basilicata; | Beaconlab (Bioinformatics, Evolution and Comparative Genomics lab), Dept of Biosciences, University on Milan | Parisi A. et al |
| EPI_ISL_477195, EPI_ISL_477201, EPI_ISL_477200, EPI_ISL_477197, EPI_ISL_477196 | | |
| Istituto Zooprofilattico Sperimentale Puglia e Basilicata; Dipartimento di Bioscienze, Biotecnologie e Biofarmaceutica dell'Università degli Studi di Bari "A.Moro"; Istituto di Biomembrane. Bioenergetica e Biotecnologie Molecolari del Consiglio Nazionale delle Ricerche di Bari | Beaconlab (Bioinformatics Evolution and Comparative Genomics lab), Dept of Biosciences, University of Milan | Parisi A. et al |
| EPI_ISL_451961 | | |
| Istituto Zooprofilattico Sperimentale Puglia e Basilicata; Dipartimento di Bioscienze, Biotecnologie e Biofarmaceutica dell'Università degli Studi di Bari "A.Moro"; Istituto di Biomembrane. Bioenergetica e Biotecnologie Molecolari del Consiglio Nazionale delle Ricerche di Bari | Beaconlab (Bioinformatics, Evolution and Comparative Genomics lab), Dept of Biosciences, University on Milan | Parisi A. et al |
| EPI_ISL_525554, EPI_ISL_451962, EPI_ISL_525555, EPI_ISL_525557, EPI_ISL_525558, EPI_ISL_525559, EPI_ISL_527380, EPI_ISL_525560, EPI_ISL_525561, EPI_ISL_525562, EPI_ISL_525563, EPI_ISL_525564, EPI_ISL_525565, EPI_ISL_525566, EPI_ISL_525567, EPI_ISL_525568, EPI_ISL_525569, EPI_ISL_525570, EPI_ISL_525571, EPI_ISL_468914, EPI_ISL_469018, EPI_ISL_469020, EPI_ISL_469021, EPI_ISL_469022, EPI_ISL_469023, EPI_ISL_525574 | | |
| IZSM | IZSM | Maurizio Viscardi et al |
| EPI_ISL_572324 | | |
| Jaber Al Ahmad Al Sabah Hospital | Dasman diabetes Institute | Fahd Al-Mulla et al |
| EPI_ISL_422424 | | |
| Johns Hopkins Hospital Department of Pathology | Johns Hopkins Hospital Department of Pathology | Peter M. Thielen et al |
| EPI_ISL_457794 | | |

| | | |
|---|---|---|
| Karolinska Universitetslaboratoriet | The Public Health Agency of Sweden | Anna-Malin Linde et al |
| EPI_ISL_454459, EPI_ISL_454460, EPI_ISL_454872, EPI_ISL_454873, EPI_ISL_454875, EPI_ISL_454878, EPI_ISL_455855, EPI_ISL_455857, EPI_ISL_455858, EPI_ISL_454886, EPI_ISL_455872, EPI_ISL_455873, EPI_ISL_455879, EPI_ISL_455890, EPI_ISL_454896 | | |
| Karolinska Universitetslaboratoriet | The Public Health Agency of Sweden | Oskar Karlsson Lindsjo et al |
| EPI_ISL_475558, EPI_ISL_475136, EPI_ISL_475095, EPI_ISL_510838 | | |
| Karolinska universitetslaboratoriet SOLNA | The Public Health Agency of Sweden | Anna-Malin Linde et al |
| EPI_ISL_534236 | | |
| Kingston Health Sciences Center | Queen's Genomics Lab at Ongwanada (Q-GLO) | Sjaarda CP et al |
| EPI_ISL_459869, EPI_ISL_459889 | | |
| Kingston Health Sciences Centre / Queen's University | Ontario Institute for Cancer Research | Prameet M. Sheth et al |
| EPI_ISL_538365, EPI_ISL_538382, EPI_ISL_538384 | | |
| Klinik Hirslanden Zurich | Institute of Medical Virology, University of Zurich | Stefan Schmutz et al |
| EPI_ISL_413021 | | |
| Klinik Hirslanden Zürich | Institute of Medical Virology, University of Zurich | Stefan Schmutz et al |
| EPI_ISL_524479 | | |
| Klinisk mikrobiologi och vardhygien Halmstad | The Public Health Agency of Sweden | Arne Kotz et al |
| EPI_ISL_429120, EPI_ISL_429122, EPI_ISL_429125 | | |
| Klinisk mikrobiologi Vasternorrland | The Public Health Agency of Sweden | Oskar Karlsson Lindsjo et al |
| EPI_ISL_510856 | | |

| | | |
|---|---|---|
| Klinisk mikrobiologi, UAS | The Public Health Agency of Sweden | Anna-Malin Linde et al |
| EPI_ISL_455904, EPI_ISL_455905, EPI_ISL_455906 | | |
| Koshigaya City Public Health Center | Pathogen Genomics Center, National Institute of Infectious Diseases | Tsuyoshi Sekizuka et al |
| EPI_ISL_480108 | | |
| KU Leuven, Clinical and Epidemiological Virology | KU Leuven, Clinical and Epidemiological Virology | Bert Vanmechelen et al |
| EPI_ISL_416470, EPI_ISL_416471, EPI_ISL_418792, EPI_ISL_415158, EPI_ISL_418793, EPI_ISL_416472, EPI_ISL_418797 | | |
| KU Leuven, Clinical and Epidemiological Virology | KU Leuven, Clinical and Epidemiological Virology | Joan Marti-Carerras et al |
| EPI_ISL_417426, EPI_ISL_417425 | | |
| KU Leuven, Clinical and Epidemiological Virology | KU Leuven, Clinical and Epidemiological Virology | Joan Marti-Carreras et al |
| EPI_ISL_420406, EPI_ISL_420397, EPI_ISL_420447, EPI_ISL_420437, EPI_ISL_420445, EPI_ISL_420327, EPI_ISL_420364, EPI_ISL_420331, EPI_ISL_420330, EPI_ISL_420356, EPI_ISL_420314, EPI_ISL_420426, EPI_ISL_420384, EPI_ISL_420439, EPI_ISL_420443, EPI_ISL_420441, EPI_ISL_420379 | | |
| KU Leuven, Clinical and Epidemiological Virology | KU Leuven, Clinical and Epidemiological Virology | Tony Wawina et al |
| EPI_ISL_418270 | | |
| KU Leuven, Rega Institute, Clinical and Epidemiological Virology | KU Leuven, Rega Institute, Clinical and Epidemiological Virology | Tony Wawina-Bokalanga et al |
| EPI_ISL_462196, EPI_ISL_462210, EPI_ISL_462199, EPI_ISL_462223, EPI_ISL_462204, EPI_ISL_462203, EPI_ISL_462181, EPI_ISL_462193, EPI_ISL_462243, EPI_ISL_464067, EPI_ISL_458235, EPI_ISL_458158, EPI_ISL_458189, EPI_ISL_458194, EPI_ISL_458201, EPI_ISL_458206, EPI_ISL_458207, EPI_ISL_458208, EPI_ISL_458217, EPI_ISL_462232, EPI_ISL_522350 | | |

| | | |
|---|---|---|
| L'Air du Temps | National Reference Center for Viruses of Respiratory Infections, Institut Pasteur, Paris | Mélanie Albert et al |
| EPI_ISL_420039, EPI_ISL_420040 | | |
| L'Hôpital Nord-Ouest Tarare-Grandris | CNR Virus des Infections Respiratoires - France SUD | Antonin Bal et al |
| EPI_ISL_508931 | | |
| La Villa Papyri | National Reference Center for Viruses of Respiratory Infections, Institut Pasteur, Paris | Mélanie Albert et al |
| EPI_ISL_443307 | | |
| Lab Microbiology, Pathology Department, William Harvey Hospital | Wellcome Sanger Institute for the COVID-19 Genomics UK Consortium | Samuel Moses et al |
| EPI_ISL_501614, EPI_ISL_501626 | | |
| Lab voor klinische biologie | Onderzoeksgroep Virologie | Laurens Lambrechts et al |
| EPI_ISL_425056, EPI_ISL_434349, EPI_ISL_451165 | | |
| LABM GH nord Essonne | National Reference Center for Viruses of Respiratory Infections, Institut Pasteur, Paris | Mélanie Albert et al |
| EPI_ISL_418234, EPI_ISL_418240 | | |
| LABM GH nord Essonne | National Reference Center for Viruses of Respiratory Infections, Institut Pasteur, Paris | Mélnie Albert et al |
| EPI_ISL_416499 | | |
| LABM GH nord Essonne de Longjumeau - BP 125 | National Reference Center for Viruses of Respiratory Infections, Institut Pasteur, Paris | Mélanie Albert et al |
| EPI_ISL_428354, EPI_ISL_428361, EPI_ISL_428365, EPI_ISL_443305, EPI_ISL_443314 | | |
| Labo Analyses Med | National Reference Center for Viruses of Respiratory Infections, Institut Pasteur, Paris | Sylvie Behillil et al |
| EPI_ISL_560637, EPI_ISL_560638, EPI_ISL_560639, EPI_ISL_560640, EPI_ISL_560641, EPI_ISL_560642 | | |
| Labo BM - Site de Juvisy - Hopital Général | National Reference Center for Viruses of Respiratory Infections, Institut Pasteur, Paris | Mélanie Albert et al |

| | | |
|---|---|---|
| EPI_ISL_420063 | | |
| Labor Kneißler GmbH & Co. KG | Heinrich Pette Institute, Leibniz Institute for Experimental Virology | Thomas Günther et al |
| EPI_ISL_476705, EPI_ISL_487400, EPI_ISL_487403, EPI_ISL_487408, EPI_ISL_487409, EPI_ISL_487410, EPI_ISL_487411, EPI_ISL_487412, EPI_ISL_487413, EPI_ISL_487414, EPI_ISL_487415, EPI_ISL_487416, EPI_ISL_487417, EPI_ISL_487418, EPI_ISL_487419, EPI_ISL_487420, EPI_ISL_487421, EPI_ISL_487422, EPI_ISL_487423, EPI_ISL_487424, EPI_ISL_487425, EPI_ISL_487427, EPI_ISL_487428 | | |
| Laboratoire de Microbiologie - Bât A - CH René Dubois | National Reference Center for Viruses of Respiratory Infections, Institut Pasteur, Paris | Mélanie Albert et al |
| EPI_ISL_443285, EPI_ISL_443286, EPI_ISL_443287 | | |
| Laboratoire de microbiologie, Hopital de Verdun | Smith Laboratory, Centre de Recherche CHU Sainte-Justine | Martin Smith et al |
| EPI_ISL_450645, EPI_ISL_463904, EPI_ISL_515214, EPI_ISL_515229 | | |
| Laboratoire de Virologie, HUG | Swiss National Reference Centre for Influenza | LAUBSCHER F. et al |
| EPI_ISL_548147, EPI_ISL_548148, EPI_ISL_548149, EPI_ISL_548150, EPI_ISL_548151, EPI_ISL_548152, EPI_ISL_548153, EPI_ISL_548154, EPI_ISL_548155, EPI_ISL_548156, EPI_ISL_548157, EPI_ISL_548158, EPI_ISL_548159, EPI_ISL_548160, EPI_ISL_548161, EPI_ISL_548162, EPI_ISL_548163, EPI_ISL_548164, EPI_ISL_548165, EPI_ISL_548166, EPI_ISL_548167, EPI_ISL_548168, EPI_ISL_548169, EPI_ISL_548170, EPI_ISL_548171, EPI_ISL_548172, EPI_ISL_548173, EPI_ISL_548174, EPI_ISL_548175, EPI_ISL_548176, EPI_ISL_548177, EPI_ISL_548178, EPI_ISL_548179, EPI_ISL_548180, EPI_ISL_548181, EPI_ISL_548182, EPI_ISL_548183, EPI_ISL_548184, EPI_ISL_548185, EPI_ISL_548186, EPI_ISL_548187, EPI_ISL_548188, EPI_ISL_548189, EPI_ISL_548190, EPI_ISL_548191, EPI_ISL_548192, EPI_ISL_548193, EPI_ISL_548194, EPI_ISL_548195, EPI_ISL_548196, EPI_ISL_548197, EPI_ISL_548198, EPI_ISL_548199, EPI_ISL_548200, EPI_ISL_548201, EPI_ISL_548202, EPI_ISL_548203, EPI_ISL_548204, EPI_ISL_548205, EPI_ISL_548206, EPI_ISL_548207, EPI_ISL_548208, EPI_ISL_548209, EPI_ISL_548210, EPI_ISL_548211, EPI_ISL_548212, EPI_ISL_548213, EPI_ISL_548214, EPI_ISL_548215, EPI_ISL_548216, EPI_ISL_548217, EPI_ISL_548218, EPI_ISL_548219, EPI_ISL_548220, EPI_ISL_548221, EPI_ISL_548222, EPI_ISL_548223, EPI_ISL_548224, EPI_ISL_548225, EPI_ISL_548226, EPI_ISL_548227, EPI_ISL_548228, EPI_ISL_548229, EPI_ISL_548230, EPI_ISL_548231, EPI_ISL_548232, EPI_ISL_548233, EPI_ISL_548234, EPI_ISL_548235, EPI_ISL_548236, EPI_ISL_548237, EPI_ISL_548238, EPI_ISL_548239, EPI_ISL_548240, EPI_ISL_548241, EPI_ISL_548242 | | |
| Laboratoire de Virologie, HUG | Swiss National Reference Centre for Influenza | LAUBSCHER |

| | | |
|---|---|---|
| | | Florian et al. et al |
| EPI_ISL_413999, EPI_ISL_414021, EPI_ISL_413997, EPI_ISL_414019, EPI_ISL_414020, EPI_ISL_414022, EPI_ISL_413996, EPI_ISL_414023 | | |
| Laboratoire National de Sante, Microbiology, Virology | Laboratoire National de Sante, Microbiology, Epidemiology and Microbial Genomics | Anke Wienecke-Baldacchino et al |
| EPI_ISL_429712, EPI_ISL_421760, EPI_ISL_445069, EPI_ISL_421735, EPI_ISL_459896, EPI_ISL_429710, EPI_ISL_459899, EPI_ISL_459902, EPI_ISL_421755, EPI_ISL_429716, EPI_ISL_421744, EPI_ISL_429732, EPI_ISL_421734, EPI_ISL_421761, EPI_ISL_429755, EPI_ISL_429714, EPI_ISL_429759 | | |
| Laboratoire National de Santé, Microbiology, Virology | Laboratoire National de Santé, Microbiology, Epidemiology and Microbial Genomics | Anke Wienecke-Baldacchino et al |
| EPI_ISL_419572, EPI_ISL_419578, EPI_ISL_419581, EPI_ISL_419584, EPI_ISL_419586, EPI_ISL_419588, EPI_ISL_419589, EPI_ISL_419593, EPI_ISL_419594, EPI_ISL_419597, EPI_ISL_419601, EPI_ISL_419602, EPI_ISL_419607 | | |
| Laboratoire Nationale de Santé, Microbiology, Virology | Laboratoire Nationale de Santé, Microbiology, Epidemiology and Microbial Genomics | Anke Wienecke-Baldacchino et al |
| EPI_ISL_417531, EPI_ISL_417532 | | |
| Laboratorio Biologia Molecolare Sars Cov2 - UOC Laboratorio Analisi - Servizio Medicina di Laboratorio , Ospedale "San Francesco" - ATS- ASSL Nuoro | Laboratorio specialistico UOC Ematologia - Ospedale "San Francesco" - ATS-ASSL Nuoro | Piras Giovanna et al |
| EPI_ISL_458085 | | |
| Laboratorio Biologia Molecolare Sars Cov2 - UOC Laboratorio Analisi - Servizio Medicina di Laboratorio, Ospedale "San Francesco" - ATS-ASSL Nuoro | Laboratorio specialistico UOC Ematologia - Ospedale "San Francesco" - ATS-ASSL Nuoro | Piras Giovanna et al |
| EPI_ISL_458084 | | |
| Laboratorio de Biologia Molecular, Facultad de Medicina, Universidad de Atacama | Center for Mathematical Modeling and Center for Genome Regulation. Santiago, Chile | Gaete A et al |
| EPI_ISL_468759 | | |

| | | |
|---|---|---|
| Laboratorio de Infecciones Respiratorias Agudas. Centro Nacional de Salud Publica, Instituto Nacional de Salud | Laboratorio de Infecciones Respiratorias Agudas. Centro Nacional de Salud Publica, Instituto Nacional de Salud | Juscamayta et al |
| EPI_ISL_547890 | | |
| Laboratorio de Referencia Nacional de Virus Respiratorios, Instituto Nacional de Salud Peru | Laboratorio de Genómica Microbiana, Universidad Peruana Cayetano Heredia | Pablo Tsukayama et al |
| EPI_ISL_568516 | | |
| Laboratorio di Microbiologia e Virologia, Università Vita-Salute San Raffaele, Milano | Laboratorio di Microbiologia e Virologia, Università Vita-Salute San Raffaele, Milano | R.A Diotti et al |
| EPI_ISL_413489 | | |
| Laboratory for Respiratory Viruses, National Influenza Centre, Cantacuzino National Military-Medical Institute for Research and Development | Cantacuzino Institute | Luiza Ustea et al |
| EPI_ISL_471420, EPI_ISL_471416, EPI_ISL_471419, EPI_ISL_471424 | | |
| Laboratory for Urgent Response to Biological Threats | Institut Pasteur CIBU /ERI | V. Caro et al |
| EPI_ISL_437690 | | |
| Laboratory of Applied Genetics | RSE "National Center for Biotechnology" | Shevtsov et al |
| EPI_ISL_435047 | | |
| Laboratory of Infectious Diseases Center of Beijing Ditan Hospital | Laboratory of Infectious Diseases Center of Beijing Ditan Hospital | Siyuan Yang et al |
| EPI_ISL_452343, EPI_ISL_452347, EPI_ISL_452341, EPI_ISL_452339 | | |
| Laboratory of Microbiology, Medical School, National and Kapodistrian University of Athens | Laboratory of Biology, Department of Medicine, Democritus University of Thrace | Kassela K. et al |
| EPI_ISL_434479, EPI_ISL_434471 | | |
| Laboratory of Molecular Virology International Center for Genetic Engineering and Biotechnology (ICGEB) | ARGO Open Lab Platform for Genome sequencing | Licastro D et al |
| EPI_ISL_417423 | | |
| Laboratory of Molecular Virology International Center for Genetic Engineering and Biotechnology (ICGEB) | ARGO Open Lab Platform for Genome Sequencing | Licastro D et al |
| EPI_ISL_428853, EPI_ISL_498559, EPI_ISL_498560, EPI_ISL_498561, EPI_ISL_498562, EPI_ISL_498563 | | |

| | | |
|---|---|---|
| Laboratory of Molecular Virology International Center fro Genetic Engineering and Biotechnology (ICGEB) | ARGO Open Lab Platform for Genome sequencing | Licastro D et al |
| EPI_ISL_417418 | | |
| Laboratory of Molecular Virology of the International Centre for Genetic Engineering and Biotechnology (ICGEB) | ARGO Open Lab Platform for Genome Sequencing | Licastro et al |
| EPI_ISL_479791, EPI_ISL_479619, EPI_ISL_479790, EPI_ISL_479618 | | |
| Laboratory of Virology, INMI Lazzaro Spallanzani IRCCS | Laboratory of Virology, INMI Lazzaro Spallanzani IRCCS | Cesare E.M. Gruber et al |
| EPI_ISL_451304 | | |
| Laboratory of Virology, INMI Lazzaro Spallanzani IRCCS | Laboratory of Virology, INMI Lazzaro Spallanzani IRCCS | Martina Rueca et al |
| EPI_ISL_451305, EPI_ISL_451303 | | |
| Lanssjukhuset Kalmar | The Public Health Agency of Sweden | Anna-Malin Linde et al |
| EPI_ISL_534237 | | |
| Le Château de Seine-Port | National Reference Center for Viruses of Respiratory Infections, Institut Pasteur, Paris | Mélanie Albert et al |
| EPI_ISL_421507 | | |
| Leeds Teaching Hospitals NHS Trust and Public Health England, National Infection Service (Leeds laboratory) | Wellcome Sanger Institute for the COVID-19 Genomics UK Consortium | Louissa Macfarlane-Smith et al |
| EPI_ISL_538988, EPI_ISL_538995, EPI_ISL_539015, EPI_ISL_539144, EPI_ISL_539124, EPI_ISL_538831, EPI_ISL_538947, EPI_ISL_538974 | | |
| Lighthouse Lab in Alderley Park | Wellcome Sanger Institute for the COVID-19 Genomics UK Consortium | The Lighthouse Lab in Alderley Park et al |
| EPI_ISL_559311, EPI_ISL_559408, EPI_ISL_559380, EPI_ISL_559327, EPI_ISL_559556, EPI_ISL_559577, EPI_ISL_559343, EPI_ISL_558902, EPI_ISL_558660, EPI_ISL_558765, EPI_ISL_555015, EPI_ISL_552731, EPI_ISL_552773, EPI_ISL_552539 | | |

| | | |
|---|---|---|
| Lighthouse Lab in Cambridge | Wellcome Sanger Institute for the COVID-19 Genomics UK Consortium | Rob Howes et al |
| EPI_ISL_556861, EPI_ISL_556858, EPI_ISL_556848, EPI_ISL_551910, EPI_ISL_551849, EPI_ISL_552173 | | |
| Lighthouse Lab in Glasgow | Wellcome Sanger Institute for the COVID-19 Genomics UK Consortium | Harper VanSteenhouse et al |
| EPI_ISL_532268, EPI_ISL_533431, EPI_ISL_532889, EPI_ISL_532743, EPI_ISL_532544, EPI_ISL_532548, EPI_ISL_532618, EPI_ISL_536971, EPI_ISL_537087, EPI_ISL_537141, EPI_ISL_532291, EPI_ISL_532504, EPI_ISL_532092, EPI_ISL_532001, EPI_ISL_537063, EPI_ISL_537081, EPI_ISL_531600, EPI_ISL_531621, EPI_ISL_531710, EPI_ISL_531089, EPI_ISL_530839, EPI_ISL_531323, EPI_ISL_531290, EPI_ISL_530814, EPI_ISL_530694, EPI_ISL_530788, EPI_ISL_530641, EPI_ISL_530724, EPI_ISL_530467, EPI_ISL_530486, EPI_ISL_540245, EPI_ISL_540306, EPI_ISL_540259, EPI_ISL_540263, EPI_ISL_540395, EPI_ISL_540095, EPI_ISL_540029, EPI_ISL_540070, EPI_ISL_530380, EPI_ISL_530358, EPI_ISL_530352, EPI_ISL_537175, EPI_ISL_537180, EPI_ISL_540042, EPI_ISL_540014, EPI_ISL_540056, EPI_ISL_539986, EPI_ISL_540086, EPI_ISL_541818, EPI_ISL_541835, EPI_ISL_532897, EPI_ISL_532676, EPI_ISL_532643, EPI_ISL_532520, EPI_ISL_531985, EPI_ISL_531751, EPI_ISL_530576, EPI_ISL_540243, EPI_ISL_539919 | | |
| Lighthouse Lab in Milton Keynes | Wellcome Sanger Institute for the COVID-19 Genomics UK Consortium | The Lighthouse Lab in Alderley Park et al |
| EPI_ISL_553008 | | |
| Lighthouse Lab in Milton Keynes | Wellcome Sanger Institute for the COVID-19 Genomics UK Consortium | The Lighthouse Lab in Milton Keynes et al |
| EPI_ISL_558284, EPI_ISL_557941, EPI_ISL_557963, EPI_ISL_556109, EPI_ISL_557191, EPI_ISL_557413, EPI_ISL_557168, EPI_ISL_557225, EPI_ISL_557277, EPI_ISL_553265, EPI_ISL_553192, EPI_ISL_553147, EPI_ISL_553088, EPI_ISL_553081, EPI_ISL_553205, EPI_ISL_553074, EPI_ISL_553214, EPI_ISL_552609, EPI_ISL_552454, EPI_ISL_552710, EPI_ISL_552450, EPI_ISL_552536, EPI_ISL_552661, EPI_ISL_552610, EPI_ISL_552656, EPI_ISL_552687, EPI_ISL_552472, EPI_ISL_552743, EPI_ISL_550435, EPI_ISL_552400, EPI_ISL_552067, EPI_ISL_552152, EPI_ISL_552078, EPI_ISL_552025, EPI_ISL_552336, EPI_ISL_552226, EPI_ISL_552318, EPI_ISL_551653, EPI_ISL_552297, EPI_ISL_552320, EPI_ISL_552331, EPI_ISL_552235, EPI_ISL_551695, EPI_ISL_551732, EPI_ISL_551595, EPI_ISL_551690, EPI_ISL_550303, EPI_ISL_550267, EPI_ISL_550269, EPI_ISL_550415, | | |

| | | |
|---|---|---|
| EPI_ISL_550288, EPI_ISL_550307, EPI_ISL_550644, EPI_ISL_550577, EPI_ISL_549891, EPI_ISL_550791, EPI_ISL_549857, EPI_ISL_550589, EPI_ISL_550704, EPI_ISL_550617, EPI_ISL_550944, EPI_ISL_551221 | | |
| Lithuanian University of Health Sciences Hospital, Department of Laboratory Medicine | Lithuanian University of Health Sciences, Laboratory of Molecular Cardiology | Lukas Zemaitis et al |
| EPI_ISL_541848, EPI_ISL_541853 | | |
| Liverpool Clinical Laboratories | COVID-19 Genomics UK (COG-UK) Consortium | Sam Haldenby et al |
| EPI_ISL_439701, EPI_ISL_440856, EPI_ISL_449469, EPI_ISL_478378, EPI_ISL_500022, EPI_ISL_500025, EPI_ISL_500033, EPI_ISL_500046, EPI_ISL_499876, EPI_ISL_500084, EPI_ISL_500086, EPI_ISL_500102, EPI_ISL_500151, EPI_ISL_499642, EPI_ISL_499632, EPI_ISL_499753, EPI_ISL_499754, EPI_ISL_499767, EPI_ISL_517010, EPI_ISL_517011, EPI_ISL_517089, EPI_ISL_517250 | | |
| M Health Fairview | Minnesota Department of Health, Public Health Laboratory | Matt Plumb et al |
| EPI_ISL_560800 | | |
| MA State Public Health Laboratory | Pathogen Discovery, Respiratory Viruses Branch, Division of Viral Diseases, Centers for Disease Control and Prevention | Ying Tao et al |
| EPI_ISL_424918 | | |
| Maine HETL | Tewhey Lab, The Jackson Laboratory | Matluk et al |
| EPI_ISL_513492, EPI_ISL_513496 | | |
| Maison de Santé du Val d'Ormois | National Reference Center for Viruses of Respiratory Infections, Institut Pasteur, Paris | Mélanie Albert et al |
| EPI_ISL_428348 | | |
| Maryland Department of Health | Maryland Department of Health | Maryland Department of Health Laboratories Administration et al |
| EPI_ISL_522851 | | |
| Massachusetts General Hospital | Infectious Disease Program, Broad Institute of Harvard and MIT | Lemieux et al |

| | | |
|---|---|---|
| EPI_ISL_460472, EPI_ISL_460333, EPI_ISL_460231 | | |
| Max von Pettenkofer Institute, Virology, National Reference Center for Retroviruses, LMU München | Laboratory for Functional Genome Analysis, Dept. Genomics, Gene Center of the LMU Munich | Max Muenchhoff et al |
| EPI_ISL_548949, EPI_ISL_548950, EPI_ISL_549025, EPI_ISL_548952, EPI_ISL_548953, EPI_ISL_548958, EPI_ISL_548960, EPI_ISL_437359, EPI_ISL_437204, EPI_ISL_437205, EPI_ISL_437206, EPI_ISL_437208, EPI_ISL_437209, EPI_ISL_437210, EPI_ISL_437211, EPI_ISL_437212, EPI_ISL_437214, EPI_ISL_437215, EPI_ISL_437216, EPI_ISL_437217, EPI_ISL_437218, EPI_ISL_437221, EPI_ISL_437223, EPI_ISL_437224, EPI_ISL_437225, EPI_ISL_437227, EPI_ISL_437228, EPI_ISL_437232, EPI_ISL_437234, EPI_ISL_437235, EPI_ISL_437237, EPI_ISL_437238, EPI_ISL_437239, EPI_ISL_437240, EPI_ISL_437241, EPI_ISL_437243, EPI_ISL_437245, EPI_ISL_437246, EPI_ISL_437247, EPI_ISL_437249, EPI_ISL_437250, EPI_ISL_437251, EPI_ISL_437252, EPI_ISL_437253, EPI_ISL_437254, EPI_ISL_437255, EPI_ISL_437256, EPI_ISL_437257, EPI_ISL_437258, EPI_ISL_437259, EPI_ISL_437260, EPI_ISL_437262, EPI_ISL_437263, EPI_ISL_437264, EPI_ISL_437265, EPI_ISL_437266, EPI_ISL_437267, EPI_ISL_437268, EPI_ISL_437269, EPI_ISL_437270, EPI_ISL_437271, EPI_ISL_437272, EPI_ISL_437273, EPI_ISL_437275, EPI_ISL_437276, EPI_ISL_437277, EPI_ISL_437278, EPI_ISL_437279, EPI_ISL_437280, EPI_ISL_437283, EPI_ISL_437284, EPI_ISL_437287, EPI_ISL_437288, EPI_ISL_437289, EPI_ISL_437290, EPI_ISL_437291, EPI_ISL_437292, EPI_ISL_437293, EPI_ISL_437295, EPI_ISL_437296, EPI_ISL_437297, EPI_ISL_451936, EPI_ISL_451938, EPI_ISL_451939, EPI_ISL_451940, EPI_ISL_451941, EPI_ISL_451942, EPI_ISL_451943, EPI_ISL_451944, EPI_ISL_451945, EPI_ISL_451947, EPI_ISL_466876, EPI_ISL_466877, EPI_ISL_466878, EPI_ISL_466879, EPI_ISL_466880, EPI_ISL_466881, EPI_ISL_466883, EPI_ISL_466886, EPI_ISL_466888, EPI_ISL_466889, EPI_ISL_466890, EPI_ISL_466891, EPI_ISL_466892, EPI_ISL_466893, EPI_ISL_466895, EPI_ISL_466896, EPI_ISL_466897, EPI_ISL_466901, EPI_ISL_466902, EPI_ISL_466903, EPI_ISL_466904, EPI_ISL_466907, EPI_ISL_466874, EPI_ISL_466918, EPI_ISL_466909, EPI_ISL_466917, EPI_ISL_466916, EPI_ISL_466922 | | |
| Max von Pettenkofer Institute, Virology, National Reference Center for Retroviruses, LMU Munich | Laboratory for Functional Genome Analysis, Dept. Genomics, Gene Center of the LMU Munich | Max Muenchhoff et al |
| EPI_ISL_420899, EPI_ISL_420900, EPI_ISL_420901, EPI_ISL_420902, EPI_ISL_420904, EPI_ISL_420905, EPI_ISL_420906, EPI_ISL_420907, EPI_ISL_420908, EPI_ISL_420909, EPI_ISL_420911, EPI_ISL_420898 | | |
| Mayo Clinic & Mayo Clinic Laboratories | Minnesota Department of Health, Public Health Laboratory | Matt Plumb et al |
| EPI_ISL_518881, EPI_ISL_539847 | | |
| Mayo Clinic Laboratories | University of Washington Virology Lab | Pavitra Roychoudhury et al |

| | | |
|---|---|---|
| EPI_ISL_500510 | | |
| MD PHL | MD PHL | Maryland Department of Health Laboratories Administration et al |
| EPI_ISL_534702, EPI_ISL_524882, EPI_ISL_534704 | | |
| Medical Disagnostics Services (MDS) | KRISP, KZN Research Innovation and Sequencing Platform | Giandhari J et al |
| EPI_ISL_495537, EPI_ISL_515839 | | |
| Medical Microbiology Unit, Department for Laboratory Medicine, Drammen Hospital, Vestre Viken Health Trust, | Norwegian Institute of Public Health, Department of Virology | Kathrine Stene-Johansen et al |
| EPI_ISL_549166 | | |
| MEPHI, Aix Marseille University | MEPHI, Aix Marseille University | Anthony LEVASSEUR et al |
| EPI_ISL_569243, EPI_ISL_569244, EPI_ISL_569246, EPI_ISL_569247, EPI_ISL_569249, EPI_ISL_569250, EPI_ISL_569252, EPI_ISL_569258, EPI_ISL_569259, EPI_ISL_569260, EPI_ISL_569261, EPI_ISL_569262, EPI_ISL_569264, EPI_ISL_569266, EPI_ISL_569271, EPI_ISL_569273, EPI_ISL_569274, EPI_ISL_569275, EPI_ISL_569278, EPI_ISL_569280, EPI_ISL_569282, EPI_ISL_569287, EPI_ISL_569290, EPI_ISL_569294, EPI_ISL_569295, EPI_ISL_569296, EPI_ISL_569298, EPI_ISL_569301, EPI_ISL_569304, EPI_ISL_569305, EPI_ISL_569309, EPI_ISL_569311, EPI_ISL_569314, EPI_ISL_569316, EPI_ISL_569324, EPI_ISL_569325, EPI_ISL_569326, EPI_ISL_569329, EPI_ISL_569331, EPI_ISL_569333, EPI_ISL_569334, EPI_ISL_569335, EPI_ISL_569336, EPI_ISL_569338, EPI_ISL_569341, EPI_ISL_569343, EPI_ISL_569346, EPI_ISL_569348, EPI_ISL_569349, EPI_ISL_569350, EPI_ISL_569351, EPI_ISL_569352, EPI_ISL_569354, EPI_ISL_569355, EPI_ISL_569357, EPI_ISL_569359, EPI_ISL_569360, EPI_ISL_569362, EPI_ISL_569363, EPI_ISL_569365, EPI_ISL_569366, EPI_ISL_569371, EPI_ISL_569374, EPI_ISL_569378, EPI_ISL_569379, EPI_ISL_569380, EPI_ISL_569381, EPI_ISL_569383, EPI_ISL_569384, EPI_ISL_569387, EPI_ISL_569388, EPI_ISL_569390, EPI_ISL_569392, EPI_ISL_569393, EPI_ISL_569394, EPI_ISL_569396, EPI_ISL_569397, EPI_ISL_569398, EPI_ISL_569399, EPI_ISL_569400, EPI_ISL_569401, EPI_ISL_569402, EPI_ISL_569404, EPI_ISL_569405, EPI_ISL_569406, EPI_ISL_569407, EPI_ISL_569408, EPI_ISL_569409, EPI_ISL_569411, EPI_ISL_569412, EPI_ISL_569413, EPI_ISL_569414, EPI_ISL_569415, EPI_ISL_569416, EPI_ISL_569417, EPI_ISL_569419, EPI_ISL_569420, EPI_ISL_569421, EPI_ISL_569422, EPI_ISL_569423, | | |

EPI_ISL_569426, EPI_ISL_569427, EPI_ISL_569429, EPI_ISL_569430, EPI_ISL_569432, EPI_ISL_569434, EPI_ISL_569435, EPI_ISL_569436, EPI_ISL_569437, EPI_ISL_569442, EPI_ISL_569447, EPI_ISL_569448, EPI_ISL_569449, EPI_ISL_569450, EPI_ISL_569451, EPI_ISL_569452, EPI_ISL_569453, EPI_ISL_569454, EPI_ISL_569455, EPI_ISL_569456, EPI_ISL_569457, EPI_ISL_569458, EPI_ISL_569459, EPI_ISL_569460, EPI_ISL_569461, EPI_ISL_569462, EPI_ISL_569465, EPI_ISL_569466, EPI_ISL_569467, EPI_ISL_569468, EPI_ISL_569469, EPI_ISL_569470, EPI_ISL_569471, EPI_ISL_569472, EPI_ISL_569474, EPI_ISL_569475, EPI_ISL_569476, EPI_ISL_569479, EPI_ISL_569480, EPI_ISL_569482, EPI_ISL_569484, EPI_ISL_569489, EPI_ISL_569490, EPI_ISL_569492, EPI_ISL_569493, EPI_ISL_569494, EPI_ISL_569495, EPI_ISL_569497, EPI_ISL_569498, EPI_ISL_569499, EPI_ISL_569500, EPI_ISL_569503, EPI_ISL_569504, EPI_ISL_569505, EPI_ISL_569506, EPI_ISL_569507, EPI_ISL_569508, EPI_ISL_569509, EPI_ISL_569510, EPI_ISL_569511, EPI_ISL_569512, EPI_ISL_569513, EPI_ISL_569514, EPI_ISL_569516, EPI_ISL_569518, EPI_ISL_569520, EPI_ISL_569521, EPI_ISL_569522, EPI_ISL_569524, EPI_ISL_569526, EPI_ISL_569527, EPI_ISL_569528, EPI_ISL_569529, EPI_ISL_569530, EPI_ISL_569531, EPI_ISL_569532, EPI_ISL_569535, EPI_ISL_569537, EPI_ISL_569538, EPI_ISL_569539, EPI_ISL_569540, EPI_ISL_569541, EPI_ISL_569543, EPI_ISL_569544, EPI_ISL_569545, EPI_ISL_569547, EPI_ISL_569548, EPI_ISL_569549, EPI_ISL_569550, EPI_ISL_569551, EPI_ISL_569552, EPI_ISL_569554, EPI_ISL_569555, EPI_ISL_569556, EPI_ISL_569557, EPI_ISL_569558, EPI_ISL_569559, EPI_ISL_569560, EPI_ISL_569561, EPI_ISL_569562, EPI_ISL_569563, EPI_ISL_569564, EPI_ISL_569565, EPI_ISL_569566, EPI_ISL_569567, EPI_ISL_569568, EPI_ISL_569569, EPI_ISL_569570, EPI_ISL_569571, EPI_ISL_569572, EPI_ISL_569574, EPI_ISL_569575, EPI_ISL_569576, EPI_ISL_569579, EPI_ISL_569580, EPI_ISL_569581, EPI_ISL_569582, EPI_ISL_569583, EPI_ISL_569585, EPI_ISL_569586, EPI_ISL_569587, EPI_ISL_569588, EPI_ISL_569589, EPI_ISL_569590, EPI_ISL_569591, EPI_ISL_569592, EPI_ISL_569593, EPI_ISL_569594, EPI_ISL_569595, EPI_ISL_569596, EPI_ISL_569597, EPI_ISL_569598, EPI_ISL_569599, EPI_ISL_569600, EPI_ISL_569601, EPI_ISL_569603, EPI_ISL_569604, EPI_ISL_569605, EPI_ISL_569606, EPI_ISL_568877, EPI_ISL_568878, EPI_ISL_568880, EPI_ISL_568881, EPI_ISL_568882, EPI_ISL_568883, EPI_ISL_568885, EPI_ISL_568888, EPI_ISL_568891, EPI_ISL_568892, EPI_ISL_568893, EPI_ISL_568894, EPI_ISL_568895, EPI_ISL_568896, EPI_ISL_568897, EPI_ISL_568898, EPI_ISL_568899, EPI_ISL_568900, EPI_ISL_568902, EPI_ISL_568903, EPI_ISL_568904, EPI_ISL_568905, EPI_ISL_568906, EPI_ISL_568907, EPI_ISL_568908, EPI_ISL_568909, EPI_ISL_568910, EPI_ISL_568915, EPI_ISL_568917, EPI_ISL_568918, EPI_ISL_568920, EPI_ISL_568921, EPI_ISL_568923, EPI_ISL_568924, EPI_ISL_568925, EPI_ISL_568926, EPI_ISL_568927, EPI_ISL_568928, EPI_ISL_568929, EPI_ISL_568930, EPI_ISL_568931, EPI_ISL_568932, EPI_ISL_568933, EPI_ISL_568934, EPI_ISL_568936, EPI_ISL_568937, EPI_ISL_568938, EPI_ISL_568939, EPI_ISL_568940, EPI_ISL_568941, EPI_ISL_568943, EPI_ISL_568944, EPI_ISL_568945, EPI_ISL_568946, EPI_ISL_568947, EPI_ISL_568948, EPI_ISL_568951, EPI_ISL_568952, EPI_ISL_568953, EPI_ISL_568954, EPI_ISL_568957, EPI_ISL_568958, EPI_ISL_568959, EPI_ISL_568961, EPI_ISL_568962, EPI_ISL_568970, EPI_ISL_568985, EPI_ISL_568986, EPI_ISL_568987, EPI_ISL_568988, EPI_ISL_568990, EPI_ISL_568991, EPI_ISL_568992, EPI_ISL_568994, EPI_ISL_568995, EPI_ISL_568996, EPI_ISL_568997, EPI_ISL_568998, EPI_ISL_568999, EPI_ISL_569000, EPI_ISL_569001,

| | | |
|---|---|---|
| EPI_ISL_569002, EPI_ISL_569003, EPI_ISL_569004, EPI_ISL_569005, EPI_ISL_569006, EPI_ISL_569007, EPI_ISL_569008, EPI_ISL_569009, EPI_ISL_569010, EPI_ISL_569011, EPI_ISL_569012, EPI_ISL_569013, EPI_ISL_569014, EPI_ISL_569015, EPI_ISL_569016, EPI_ISL_569017, EPI_ISL_569018, EPI_ISL_569019, EPI_ISL_569020, EPI_ISL_569021, EPI_ISL_569022, EPI_ISL_569023, EPI_ISL_569024, EPI_ISL_569025, EPI_ISL_569026, EPI_ISL_569027, EPI_ISL_569028, EPI_ISL_569029, EPI_ISL_569030, EPI_ISL_569031, EPI_ISL_569033, EPI_ISL_569034, EPI_ISL_569035, EPI_ISL_569036, EPI_ISL_569037, EPI_ISL_569038, EPI_ISL_569039, EPI_ISL_569040, EPI_ISL_569042, EPI_ISL_569043, EPI_ISL_569044, EPI_ISL_569045, EPI_ISL_569047, EPI_ISL_569048, EPI_ISL_569049, EPI_ISL_569051, EPI_ISL_569052, EPI_ISL_569053, EPI_ISL_569054, EPI_ISL_569055, EPI_ISL_569057, EPI_ISL_569058, EPI_ISL_569059, EPI_ISL_569060, EPI_ISL_569061, EPI_ISL_569062, EPI_ISL_569063, EPI_ISL_569064, EPI_ISL_569065, EPI_ISL_569068, EPI_ISL_569069, EPI_ISL_569070, EPI_ISL_569072, EPI_ISL_569074, EPI_ISL_569075, EPI_ISL_569076, EPI_ISL_569077, EPI_ISL_569078, EPI_ISL_569079, EPI_ISL_569080, EPI_ISL_569081, EPI_ISL_569082, EPI_ISL_569083, EPI_ISL_569086, EPI_ISL_569087, EPI_ISL_569089, EPI_ISL_569091, EPI_ISL_569092, EPI_ISL_569095, EPI_ISL_569098, EPI_ISL_569099, EPI_ISL_569100, EPI_ISL_569101, EPI_ISL_569102, EPI_ISL_569105, EPI_ISL_569106, EPI_ISL_569107, EPI_ISL_569108, EPI_ISL_569110, EPI_ISL_569111, EPI_ISL_569113, EPI_ISL_569114, EPI_ISL_569115, EPI_ISL_569116, EPI_ISL_569117, EPI_ISL_569118, EPI_ISL_569122, EPI_ISL_569123, EPI_ISL_569124, EPI_ISL_569125, EPI_ISL_569126, EPI_ISL_569127, EPI_ISL_569133, EPI_ISL_569134, EPI_ISL_569135, EPI_ISL_569137, EPI_ISL_569138, EPI_ISL_569139, EPI_ISL_569140, EPI_ISL_569141, EPI_ISL_569142, EPI_ISL_569143, EPI_ISL_569144, EPI_ISL_569145, EPI_ISL_569146, EPI_ISL_569147, EPI_ISL_569148, EPI_ISL_569150, EPI_ISL_569151, EPI_ISL_569152, EPI_ISL_569153, EPI_ISL_569154, EPI_ISL_569156, EPI_ISL_569158, EPI_ISL_569159, EPI_ISL_569162, EPI_ISL_569163, EPI_ISL_569164, EPI_ISL_569165, EPI_ISL_569166, EPI_ISL_569167, EPI_ISL_569168, EPI_ISL_569169, EPI_ISL_569170, EPI_ISL_569171, EPI_ISL_569172, EPI_ISL_569173, EPI_ISL_569174, EPI_ISL_569176, EPI_ISL_569177, EPI_ISL_569178, EPI_ISL_569179, EPI_ISL_569180, EPI_ISL_569181, EPI_ISL_569182, EPI_ISL_569183, EPI_ISL_569184, EPI_ISL_569185, EPI_ISL_569186, EPI_ISL_569188, EPI_ISL_569189, EPI_ISL_569190, EPI_ISL_569191, EPI_ISL_569192, EPI_ISL_569193, EPI_ISL_569195, EPI_ISL_569196 | | |
| MHC West-Brabant | Erasmus Medical Center | David Nieuwenhuijse et al |
| EPI_ISL_413574 | | |
| Michigan Department of Health and Human Services, Bureau of Laboratories | Michigan Department of Health and Human Services, Bureau of Laboratories | Blankenship HM et al |
| EPI_ISL_436819, EPI_ISL_447178, EPI_ISL_450593, EPI_ISL_460009, EPI_ISL_507428, EPI_ISL_507557, EPI_ISL_507651, EPI_ISL_516197, EPI_ISL_516204, EPI_ISL_516382, | | |

| | | |
|---|---|---|
| EPI_ISL_516395, EPI_ISL_529890, EPI_ISL_529891, EPI_ISL_529901, EPI_ISL_565844, EPI_ISL_565850, EPI_ISL_565987, EPI_ISL_566005 | | |
| Microbiological Diagnostic Unit Public Health Laboratory | Microbiological Diagnostic Unit Public Health Laboratory | Seemann T. et al |
| EPI_ISL_430644 | | |
| Microbiology Division, Barzilai University Medical Center | Stern Lab | Stern Lab et al |
| EPI_ISL_447294 | | |
| Microbiology Division, South Carolina Department of Health and Environmental Control | Microbiology Division, South Carolina Department of Health and Environmental Control | Flores et al |
| EPI_ISL_541665 | | |
| Microbiology, Virology and Biemergency Laboratory-ASST FBF Sacco | Microbiology, Virology and Biemergency Laboratory-ASST FBF Sacco | Mancon A et al |
| EPI_ISL_486647, EPI_ISL_486651, EPI_ISL_486655, EPI_ISL_486646, EPI_ISL_486662 | | |
| Microbiology, Virology and Biemergency Laboratory-ASST FBF Sacco | Microbiology, Virology and Biemergency Laboratory-ASST FBF Sacco | Micheli V et al |
| EPI_ISL_486652 | | |
| Microbiology, Virology and Biemergency Laboratory-ASST FBF Sacco | Microbiology, Virology and Biemergency Laboratory-ASST FBF Sacco | Rimoldi SG et al |
| EPI_ISL_486653, EPI_ISL_486664 | | |
| Microbiology, Virology and Biemergency Laboratory-ASST FBF Sacco | Microbiology, Virology and Biemergency Laboratory-ASST FBF Sacco | Romeri F et al |
| EPI_ISL_486650, EPI_ISL_486658 | | |
| Ministry of Health Turkey | Ministry of Health Turkey | Fatma Bayrakdar et al |
| EPI_ISL_437315 | | |
| Minnesota Department of Health, Public Health Laboratory | Minnesota Department of Health, Public Health Laboratory | Matt Plumb et al |
| EPI_ISL_482998, EPI_ISL_507976, EPI_ISL_527587, EPI_ISL_527615, EPI_ISL_527620, EPI_ISL_450768, EPI_ISL_414589, EPI_ISL_417479, EPI_ISL_417502 | | |
| Mohammed Bin Rashid University of Medicine and Health Sciences | Al Jalila Genomics Center | Ahmad Abou |

| | | |
|---|---|---|
| | | Tayoun et al |
| EPI_ISL_520678 | | |
| Molecular diagnostic laboratory of Federal Budget Institution of Science "Central Research Institute of Epidemiology" of The Federal Service on Customers' Rights Protection and Human Well-being Surveillance | Group of Genomics and Postgenomic Technologies of Central Research Institute of Epidemilology | Speranskaya AS et al |
| EPI_ISL_534343 | | |
| Molecular Virology Unit, Fondazione IRCCS Policlinico San Matteo , Pavia | Laboratory of Virology, INMI Lazzaro Spallanzani IRCCS | Antonio Piralla et al |
| EPI_ISL_451308, EPI_ISL_451306 | | |
| Molecular Virology Unit, Fondazione IRCCS Policlinico San Matteo , Pavia | Laboratory of Virology, INMI Lazzaro Spallanzani IRCCS | Fausto Baldanti et al |
| EPI_ISL_460081 | | |
| Molecular Virology Unit, Fondazione IRCCS Policlinico San Matteo , Pavia | Laboratory of Virology, INMI Lazzaro Spallanzani IRCCS | Maria R. Capobianchi et al |
| EPI_ISL_460086 | | |
| Molecular Virology Unit, Fondazione IRCCS Policlinico San Matteo , Pavia | Laboratory of Virology, INMI Lazzaro Spallanzani IRCCS | Martina Rueca et al |
| EPI_ISL_460082 | | |
| Motol University Hospital | Institute of Applied Biotechnologies a.s. | Petr Brož et al |
| EPI_ISL_426893 | | |
| MSHS Clinical Microbiology Laboratories | MSHS Pathogen Surveillance Program | Ana S. Gonzalez-Reiche et al |
| EPI_ISL_421352, EPI_ISL_450025, EPI_ISL_450055 | | |
| München Klinik Schwabing | MGZ Medical Genetics Center | Dieter A. Wolf et al |
| EPI_ISL_490205, EPI_ISL_490206, EPI_ISL_490207 | | |
| National Centre For Cell Science | National Centre For Cell Science | Dhiraj Paul et al |
| EPI_ISL_496562, EPI_ISL_496566 | | |

| | | |
|---|---|---|
| National Centre for Disease control (NCDC) | NCDC/CSIR-IGIB | Pramod Kumar# et al |
| EPI_ISL_436453, EPI_ISL_436463 | | |
| National Institute for Communicable Disease Control and Prevention (ICDC) Chinese Center for Disease Control and Prevention (China CDC) | National Institute for Communicable Disease Control and Prevention (ICDC) Chinese Center for Disease Control and Prevention (China CDC) | Zhang et al |
| EPI_ISL_402125 | | |
| National Institute for Communicable Diseases of the National Health Laboratory Service | National Institute for Communicable Diseases of the National Health Laboratory Service | Allam M et al |
| EPI_ISL_515156, EPI_ISL_490275 | | |
| National Institute of Virology, NIV Influenza | National Institute of Virology, NIV Influenza | Potdar V et al |
| EPI_ISL_541699 | | |
| National Public Health Laboratory, National Centre for Infectious Diseases | National Public Health Laboratory, National Centre for Infectious Diseases | Mak TM et al |
| EPI_ISL_462369, EPI_ISL_469147, EPI_ISL_483607, EPI_ISL_498579 | | |
| National Public Health Laboratory, National Centre for Infectious Diseases | National Public Health Laboratory, National Centre for Infectious Diseases | Mak Tze Minn et al |
| EPI_ISL_443208 | | |
| National Virus Reference Laboratory | National Virus Reference Laboratory | Michael Carr et al |
| EPI_ISL_528462 | | |
| National Virus Resource Center, Chinese Academy of Sciences, Wuhan 430071, China | Computational Virology Group, Center for Bacteria and Viruses Resources and Bioinformation, Wuhan Institute of Virology, Chinese Academy of Sciences，Wuhan 430071, China | Jianjun Chen et al |
| EPI_ISL_493177 | | |
| Naval Health Research Center | Naval Medical Research Center Biological Defense Research Directorate | Logan Voegtly et al |
| EPI_ISL_444999 | | |
| Nevada State Public Health Laboratory | Nevada State Public Health Laboratory | Richard Tillett et al |
| EPI_ISL_515297, EPI_ISL_515320, EPI_ISL_515384 | | |

| | | |
|---|---|---|
| New Mexico Department of Health Scientific Laboratory | New Mexico Department of Health Scientific Laboratory | Ellie Johnson et al |
| EPI_ISL_535300, EPI_ISL_535303, EPI_ISL_542025, EPI_ISL_542032, EPI_ISL_542059 | | |
| New Mexico Department of Health Scientific Laboratory Division | Center for Global Health, University of New Mexico Health Sciences Center | Daryl Domman et al |
| EPI_ISL_508108 | | |
| NewYork-Presbyterian & Mason Lab | Mason Lab | Daniel J. Butler et al |
| EPI_ISL_427576, EPI_ISL_427581, EPI_ISL_427584, EPI_ISL_427600 | | |
| NHSGGC West of Scotland Specialist Virology Centre / MRC-University of Glasgow Centre for Virus Research | Wellcome Sanger Institute for the COVID-19 Genomics UK Consortium | Ana da Silva Filipe et al |
| EPI_ISL_459548, EPI_ISL_532932, EPI_ISL_532563, EPI_ISL_531982, EPI_ISL_530959, EPI_ISL_534660, EPI_ISL_534549, EPI_ISL_534583, EPI_ISL_530488, EPI_ISL_534493 | | |
| NIV Influenza | NIV Influenza | Potdar V et al |
| EPI_ISL_452208 | | |
| Norra Alvsborgs lanssjukhus | The Public Health Agency of Sweden | Anna-Malin Linde et al |
| EPI_ISL_534240 | | |
| North West London Pathology, Imperial College Healthcare NHS Trust | Wellcome Sanger Institute for the COVID-19 Genomics UK Consortium | Ling Li et al |
| EPI_ISL_516867, EPI_ISL_524705, EPI_ISL_524630, EPI_ISL_524711 | | |
| Northumbria University / South Tees Hospitals NHS Foundation Trust / North Cumbria Integrated Care NHS Foundation Trust / North Tees and Hartlepool NHS Foundation Trust / Newcastle Hospitals NHS Foundation Trust | COVID-19 Genomics UK (COG-UK) Consortium | Darren L Smith et al |

| | | |
|---|---|---|
| EPI_ISL_472172, EPI_ISL_472190, EPI_ISL_472251, EPI_ISL_478587, EPI_ISL_484327 | | |
| NRL for Influenza, Centrum Epidemiology and Microbiology of National Institute of Public Health, Czech Republic | Charite Universitaetsmedizin Berlin, Institute of Virology | Victor M Corman et al |
| EPI_ISL_416743 | | |
| NU-OMICS DNA Sequencing research facility, Northumbria University | Wellcome Sanger Institute for the COVID-19 Genomics UK Consortium | Chris Duncan et al |
| EPI_ISL_488817, EPI_ISL_488807, EPI_ISL_488748, EPI_ISL_488806, EPI_ISL_488747, EPI_ISL_488759, EPI_ISL_488687, EPI_ISL_488601, EPI_ISL_488627, EPI_ISL_488587, EPI_ISL_488820, EPI_ISL_458678, EPI_ISL_488727, EPI_ISL_488721, EPI_ISL_488141, EPI_ISL_488172 | | |
| NYU Langone Health | Department of Pathology and Medicine, New York University School of Medicine | Margaret Black et al |
| EPI_ISL_418200 | | |
| NYU Langone Health | Department of Pathology and Medicine, New York University School of Medicine | Maria Aguero-Rosenfeld et al |
| EPI_ISL_418969 | | |
| NYU Langone Health | Departments of Pathology and Medicine, New York University School of Medicine | Maria Aguero-Rosenfeld et al |
| EPI_ISL_428786, EPI_ISL_430377, EPI_ISL_419697, EPI_ISL_444784, EPI_ISL_420584, EPI_ISL_450398, EPI_ISL_451404, EPI_ISL_451469, EPI_ISL_456056, EPI_ISL_421591, EPI_ISL_467420 | | |
| OHSU Lab Services Molecular Microbiology Lab | Oregon SARS-CoV-2 Genome Sequencing Center | Brendan L. O'Connell et al |
| EPI_ISL_525957, EPI_ISL_525963, EPI_ISL_509147, EPI_ISL_509159, EPI_ISL_509177, EPI_ISL_509192, EPI_ISL_509211, EPI_ISL_526038, EPI_ISL_526045, EPI_ISL_526079 | | |
| Oman-NIC | Department of Microbiology and Immunology-SQUH | Fahad Zadjali et al |
| EPI_ISL_491986 | | |

| | | |
|---|---|---|
| Oman-NIC | Oman-NIC | Samira Al-Maruqi et al |
| EPI_ISL_457977 | | |
| Omsk Research Institute of Natural Focal Infections | WHO National Influenza Centre Russian Federation | Artem Fadeev et al |
| EPI_ISL_569836, EPI_ISL_569804, EPI_ISL_569809, EPI_ISL_569851, EPI_ISL_569801, EPI_ISL_569794, EPI_ISL_569843 | | |
| Omtanken Grimmered | The Public Health Agency of Sweden | Oskar Karlsson Lindsjo et al |
| EPI_ISL_475535 | | |
| Orange County Public Health Laboratory | Chan-Zuckerberg Biohub | CZB Cliahub Consortium et al |
| EPI_ISL_548291, EPI_ISL_548416, EPI_ISL_548465, EPI_ISL_548592, EPI_ISL_548677, EPI_ISL_548601 | | |
| Orebro klinisk mikrobiologi | The Public Health Agency of Sweden | Oskar Karlsson Lindsjo et al |
| EPI_ISL_475135 | | |
| Originating lab: Wales Specialist Virology Centre Sequencing lab: Pathogen Genomics Unit | COVID-19 Genomics UK (COG-UK) Consortium | Catherine Moore et al |
| EPI_ISL_494277, EPI_ISL_474230, EPI_ISL_474291 | | |
| Osmania Medical College | CSIR-Centre for Cellular and Molecular Biology | Shashikala Reddy et al |
| EPI_ISL_458066 | | |
| Ospedale "Giuseppe Mazzini"-Teramo | Istitituto Zooprofilattico Sperimentale dell'Abruzzo e Molise "G.Caporale" | Lorusso A et al |
| EPI_ISL_529018, EPI_ISL_528922, EPI_ISL_528924, EPI_ISL_528926, EPI_ISL_528927, EPI_ISL_528928 | | |
| Ospedale "Ss. Annunziata" | Istitituto Zooprofilattico Sperimentale dell'Abruzzo e Molise "G.Caporale" | Lorusso A et al |
| EPI_ISL_529022, EPI_ISL_529014, EPI_ISL_529015 | | |

| | | |
|---|---|---|
| Ospedale “San Liberatore” di Atri | Istituto Zooprofilattico Sperimentale dell'Abruzzo e Molise "G. Caporale" | Lorusso A et al |
| EPI_ISL_418256 | | |
| Ospedale Civile Castel Di Sangro | Istituto Zooprofilattico Sperimentale dell'Abruzzo e Molise "G.Caporale" | Lorusso A et al |
| EPI_ISL_420564 | | |
| Ospedale Civile Giuseppe Mazzini | Istituto Zooprofilattico Sperimentale dell'Abruzzo e Molise "G. Caporale" | Lorusso A et al |
| EPI_ISL_418260, EPI_ISL_418261, EPI_ISL_429228, EPI_ISL_429231, EPI_ISL_429233, EPI_ISL_429234, EPI_ISL_429235 | | |
| Ospedale Civile Giuseppe Mazzini | Istituto Zooprofilattico Sperimentale dell'Abruzzo e Molise "G.Caporale" | Lorusso A et al |
| EPI_ISL_420568, EPI_ISL_420565 | | |
| Ospedale Civile Giuseppe Mazzini, Teramo | Istituto Zooprofilattico Sperimentale dell'Abruzzo e Molise "G.Caporale" | Lorusso A et al |
| EPI_ISL_418257 | | |
| Ospedale Civile Maria SS. dello Splendore | Istitituto Zooprofilattico Sperimentale dell'Abruzzo e Molise "G.Caporale" | Lorusso A et al |
| EPI_ISL_528990 | | |
| Ospedale Civile S. Liberatore di Atri | Istituto Zooprofilattico Sperimentale dell'Abruzzo e Molise "G.Caporale" | Lorusso A et al |
| EPI_ISL_436731, EPI_ISL_436732, EPI_ISL_436719, EPI_ISL_436720, EPI_ISL_436722, EPI_ISL_436724 | | |
| Ospedale Civile S. Liberatore-Atri | Istitituto Zooprofilattico Sperimentale dell'Abruzzo e Molise "G.Caporale" | Lorusso A et al |
| EPI_ISL_529020, EPI_ISL_529021, EPI_ISL_528929, EPI_ISL_529009 | | |
| Ospedale Regionale San Salvatore | Istituto Zooprofilattico Sperimentale dell'Abruzzo e Molise "G. Caporale" | Lorusso A et al |
| EPI_ISL_429229 | | |
| Ospedale Regionale San Salvatore | Istituto Zooprofilattico Sperimentale dell'Abruzzo e Molise "G.Caporale" | Lorusso A et al |
| EPI_ISL_420567 | | |
| Ospedale SS Annunziata | Istituto Zooprofilattico Sperimentale dell'Abruzzo e Molise "G.Caporale" | Lorusso A et al |
| EPI_ISL_435150, EPI_ISL_435151, EPI_ISL_435148 | | |
| Ospedale SS Annunziata-Sulmona | Istitituto Zooprofilattico Sperimentale dell'Abruzzo e Molise "G.Caporale" | Lorusso A et al |
| EPI_ISL_528991, EPI_ISL_529016, EPI_ISL_528992 | | |

| | | |
|---|---|---|
| Ostfold Hospital Trust - Kalnes, Centre for Laboratory Medicine, Section for gene technology and infection serology | Norwegian Institute of Public Health, Department of Virology | Kathrine Stene-Johansen et al |
| EPI_ISL_549117, EPI_ISL_549118 | | |
| Oxford Viromics, NDM, University of Oxford; Oxford University Hospitals; Basingstoke and North Hampshire Hospital | COVID-19 Genomics UK (COG-UK) Consortium | Tanya Golubchik et al |
| EPI_ISL_478823, EPI_ISL_478850, EPI_ISL_478865, EPI_ISL_479038, EPI_ISL_479119, EPI_ISL_479144, EPI_ISL_534773, EPI_ISL_534795, EPI_ISL_534808, EPI_ISL_534830, EPI_ISL_534938, EPI_ISL_549345, EPI_ISL_559754, EPI_ISL_559757, EPI_ISL_559782, EPI_ISL_559990 | | |
| Parc des Dames | National Reference Center for Viruses of Respiratory Infections, Institut Pasteur, Paris | Mélanie Albert et al |
| EPI_ISL_421502 | | |
| Pathogen Genomics Lab King Abdullah University of Science and Technology(KAUST) | Pathogen Genomics Lab King Abdullah University of Science and Technology(KAUST) | Afrah Alsomali et al |
| EPI_ISL_513008 | | |
| Pathogen Genomics Lab King Abdullah University of Science and Technology(KAUST) | Pathogen Genomics Lab King Abdullah University of Science and Technology(KAUST) | Fadwa Alofi et al |
| EPI_ISL_512922, EPI_ISL_512923, EPI_ISL_512926 | | |
| Pathogen Genomics Lab King Abdullah University of Science and Technology(KAUST) | Pathogen Genomics Lab King Abdullah University of Science and Technology(KAUST) | Fathia Ben Rached et al |
| EPI_ISL_513081, EPI_ISL_513102 | | |
| Pathogen Genomics Lab King Abdullah University of Science and Technology(KAUST) | Pathogen Genomics Lab King Abdullah University of Science and Technology(KAUST) | Raeece Naeem et al |
| EPI_ISL_512880, EPI_ISL_512889, EPI_ISL_512891, EPI_ISL_512893, EPI_ISL_512894, EPI_ISL_512897, EPI_ISL_513065, EPI_ISL_513211 | | |
| Pathogen Genomics Lab King Abdullah University of Science and Technology(KAUST) | Pathogen Genomics Lab King Abdullah University of Science and Technology(KAUST) | Rahul P Salunke et al |
| EPI_ISL_513036, EPI_ISL_513049, EPI_ISL_513054, EPI_ISL_513198 | | |

| | | |
|---|---|---|
| Pathogen Genomics Lab King Abdullah University of Science and Technology(KAUST) | Pathogen Genomics Lab King Abdullah University of Science and Technology(KAUST) | Sara Mfarrej et al |
| EPI_ISL_512977 | | |
| Pathogen Genomics Lab King Abdullah University of Science and Technology(KAUST) | Pathogen Genomics Lab King Abdullah University of Science and Technology(KAUST) | Sharif Hala et al |
| EPI_ISL_437754 | | |
| Pathology West - NSW Health Pathology | NSW Health Pathology - Institute of Clinical Pathology and Medical Research; Westmead Hospital; University of Sydney | CIDM-PH et al. et al |
| EPI_ISL_451577 | | |
| PHE South West Regional Laboratory, National Infection Service | Wellcome Sanger Institute for the COVID-19 Genomics UK Consortium | Stephanie Hutchings et al |
| EPI_ISL_440154, EPI_ISL_440269, EPI_ISL_440718, EPI_ISL_442642, EPI_ISL_492793, EPI_ISL_443884, EPI_ISL_443973, EPI_ISL_443697, EPI_ISL_443818, EPI_ISL_443913, EPI_ISL_443791, EPI_ISL_469568, EPI_ISL_458862, EPI_ISL_488435, EPI_ISL_488237, EPI_ISL_488444, EPI_ISL_488223, EPI_ISL_488349, EPI_ISL_492689, EPI_ISL_492694, EPI_ISL_492575, EPI_ISL_501588, EPI_ISL_501567, EPI_ISL_492311, EPI_ISL_495101 | | |
| Plaisance | National Reference Center for Viruses of Respiratory Infections, Institut Pasteur, Paris | Mélanie Albert et al |
| EPI_ISL_443308 | | |
| Presidio Ospedaliero "S. Spirito" - PESCARA | Istituto Zooprofilattico Sperimentale dell'Abruzzo e Molise "G. Caporale" | Lorusso A et al |
| EPI_ISL_418255 | | |
| Presidio Ospedaliero "S.Filippo e Nicola"-Avezzano | Istitituto Zooprofilattico Sperimentale dell'Abruzzo e Molise "G.Caporale" | Lorusso A et al |
| EPI_ISL_529013 | | |
| Presidio Ospedaliero "Santo Spirito"-Pescara | Istitituto Zooprofilattico Sperimentale dell'Abruzzo e Molise "G.Caporale" | Lorusso A et al |
| EPI_ISL_528920, EPI_ISL_528921 | | |
| Presidio Ospedaliero Santo Spirito | Istituto Zooprofilattico Sperimentale dell'Abruzzo e Molise "G. Caporale" | Lorusso A et al |
| EPI_ISL_429226 | | |
| Prof. Massimo Zollo CEINGE TASK-FORCE COVID19 - Regione Campania | Prof. Massimo Zollo CEINGE TASK-FORCE COVID19 - Regione Campania | Veronica Ferrucci et al |
| EPI_ISL_514432 | | |

| | | |
|---|---|---|
| Prof. Massimo Zollo CEINGE TASK-FORCE COVID19 - Regione Campania | Prof. Massimo Zollo CEINGE TASK-FORCE COVID19 - Regione Campania | Veronica Ferrucci1 et al |
| EPI_ISL_477204 | | |
| Providence St. Joseph Health Molecular Genomics Laboratory | Providence St. Joseph Health Molecular Genomics Laboratory | Alexa K Dowdell et al |
| EPI_ISL_482403, EPI_ISL_482447 | | |
| Public Health Laboratory | National Microbiology Laboratory | Anna Majer et al |
| EPI_ISL_469239, EPI_ISL_469226, EPI_ISL_469227, EPI_ISL_469228, EPI_ISL_469229, EPI_ISL_469230, EPI_ISL_469231, EPI_ISL_469232, EPI_ISL_469233, EPI_ISL_469235, EPI_ISL_469238 | | |
| Quadram Institute Bioscience | COVID-19 Genomics UK (COG-UK) Consortium | Dave J. Baker et al |
| EPI_ISL_457333 | | |
| Queens Medical Centre, Clinical Microbiology Department / DeepSeq Nottingham | COVID-19 Genomics UK (COG-UK) Consortium | Gemma Clark et al |
| EPI_ISL_425512, EPI_ISL_425584, EPI_ISL_432928 | | |
| Quest Diagnostics | Quest Diagnostics | Rosenthal et al |
| EPI_ISL_571267, EPI_ISL_571181, EPI_ISL_571593, EPI_ISL_571030, EPI_ISL_571062, EPI_ISL_571118, EPI_ISL_571131, EPI_ISL_571362, EPI_ISL_571638, EPI_ISL_571760, EPI_ISL_571762, EPI_ISL_571381, EPI_ISL_571387, EPI_ISL_571395, EPI_ISL_572011, EPI_ISL_571399 | | |
| Quick Care Huron | South Dakota Public Health Laboratory | Matt Plumb et al |
| EPI_ISL_569615 | | |
| R. G. Lugar Center for Public Health Research, National Center for Disease Control and Public Health (NCDC) of Georgia. | R. G. Lugar Center for Public Health Research, National Center for Disease Control and Public Health (NCDC) of Georgia. | Adam Kotorashvili et al |
| EPI_ISL_416482 | | |
| Regional Virus Laboratory, Belfast Health and Social Care Trust | COVID-19 Genomics UK (COG-UK) Consortium | Conall McCaughey et al |
| EPI_ISL_453483, EPI_ISL_441381, EPI_ISL_441382, EPI_ISL_461786, EPI_ISL_441417, EPI_ISL_441425, EPI_ISL_441433 | | |

| | | |
|---|---|---|
| Regional Virus Laboratory, Belfast Health and Social Care Trust | Wellcome Sanger Institute for the COVID-19 Genomics UK Consortium | Conall McCaughey et al |
| EPI_ISL_489196, EPI_ISL_482091, EPI_ISL_441781 | | |
| Résidence de maintenon | National Reference Center for Viruses of Respiratory Infections, Institut Pasteur, Paris | Mélanie Albert et al |
| EPI_ISL_420054 | | |
| Résidence Eleusis | National Reference Center for Viruses of Respiratory Infections, Institut Pasteur, Paris | Mélanie Albert et al |
| EPI_ISL_420051 | | |
| Résidence Esterel | National Reference Center for Viruses of Respiratory Infections, Institut Pasteur, Paris | Mélanie Albert et al |
| EPI_ISL_443304 | | |
| Résidence Les Marines | National Reference Center for Viruses of Respiratory Infections, Institut Pasteur, Paris | Mélanie Albert et al |
| EPI_ISL_443303 | | |
| Résidence Ornano | National Reference Center for Viruses of Respiratory Infections, Institut Pasteur, Paris | Mélanie Albert et al |
| EPI_ISL_443258, EPI_ISL_443259 | | |
| Résidence Villa Caroline | National Reference Center for Viruses of Respiratory Infections, Institut Pasteur, Paris | Mélanie Albert et al |
| EPI_ISL_420047 | | |
| Respiratory Virus Unit, Microbiology Services Colindale, Public Health England | Respiratory Virus Unit, Microbiology Services Colindale, Public Health England | Monica Galiano et al |
| EPI_ISL_414013, EPI_ISL_417233, EPI_ISL_417236, EPI_ISL_417246, EPI_ISL_417252, EPI_ISL_417255, EPI_ISL_417270, EPI_ISL_417301, EPI_ISL_417302, EPI_ISL_423900, EPI_ISL_424046, EPI_ISL_424082, EPI_ISL_424084, EPI_ISL_423784, EPI_ISL_423818, EPI_ISL_423236, EPI_ISL_420636, EPI_ISL_423298, EPI_ISL_423324, EPI_ISL_421892, EPI_ISL_420693, EPI_ISL_423358, EPI_ISL_420765, EPI_ISL_420771 | | |
| Respiratory Virus Unit, Microbiology Services Colindale, Public Health England | Respiratory Virus Unit, Microbiology Services Colindale, Public Health England | PHE Covid Sequencing Team et al |

| | | |
|---|---|---|
| EPI_ISL_464194, EPI_ISL_464215, EPI_ISL_464362, EPI_ISL_464404, EPI_ISL_464541, EPI_ISL_464582, EPI_ISL_464655, EPI_ISL_464719, EPI_ISL_464745, EPI_ISL_464772, EPI_ISL_464802, EPI_ISL_464901, EPI_ISL_465017, EPI_ISL_465023, EPI_ISL_465058, EPI_ISL_465086, EPI_ISL_465091, EPI_ISL_465858, EPI_ISL_465937, EPI_ISL_465704, EPI_ISL_465612, EPI_ISL_465618, EPI_ISL_490224 | | |
| Robert Garry lab | Andersen lab at Scripps Research | Allison Smither et al |
| EPI_ISL_434520 | | |
| Robert Koch Institute, National Reference center for Influenza, Berlin, Germany | Robert Koch Institute, Bioinformatics MF1, Berlin, Germany | Marianne Wedde et al |
| EPI_ISL_481264, EPI_ISL_481253, EPI_ISL_481262 | | |
| Robert Koch Institute, ZBS1 Highly Pathogenic Viruses, Berlin, Germany | Robert Koch Institute, Bioinformatics MF1, Berlin, Germany | Janine Michel et al |
| EPI_ISL_483139, EPI_ISL_483140, EPI_ISL_483141, EPI_ISL_483143, EPI_ISL_483144, EPI_ISL_483145, EPI_ISL_483146, EPI_ISL_483147, EPI_ISL_483148, EPI_ISL_483149, EPI_ISL_483150, EPI_ISL_483151, EPI_ISL_483152, EPI_ISL_483153, EPI_ISL_483154, EPI_ISL_483155, EPI_ISL_483156, EPI_ISL_483157 | | |
| RSA San Raffaele Sulmona | Istitituto Zooprofilattico Sperimentale dell'Abruzzo e Molise "G.Caporale" | Lorusso A et al |
| EPI_ISL_529017 | | |
| RSA/RP Villa San Giovanni - Gruppo Edos | Istitituto Zooprofilattico Sperimentale dell'Abruzzo e Molise "G.Caporale" | Lorusso A et al |
| EPI_ISL_529019 | | |
| RSA/RP Villa San Giovanni - Gruppo Edos | Istituto Zooprofilattico Sperimentale dell'Abruzzo e Molise "G.Caporale" | Lorusso A et al |
| EPI_ISL_436725 | | |
| SA Pathology | SA Pathology | Lex Leong et al |
| EPI_ISL_468009, EPI_ISL_451092, EPI_ISL_508124 | | |
| San Diego County Public Health Laboratory | Andersen lab at Scripps Research | SEARCH Alliance San Diego with Tracy Basler et al |

| | | |
|---|---|---|
| EPI_ISL_494456, EPI_ISL_494590 | | |
| San Joaquin County Public Health Lab | Chan-Zuckerberg Biohub | CZB Cliahub Consortium et al |
| EPI_ISL_486335 | | |
| San Matteo Hospital Pavia | Dep. Of Oncology and Hemato-Oncology University of Milan | Claudia Alteri et al |
| EPI_ISL_542278, EPI_ISL_542281, EPI_ISL_542282, EPI_ISL_542283, EPI_ISL_542284, EPI_ISL_542285, EPI_ISL_542288, EPI_ISL_542289, EPI_ISL_542292, EPI_ISL_542294, EPI_ISL_542295, EPI_ISL_542296, EPI_ISL_542297, EPI_ISL_542298, EPI_ISL_542299, EPI_ISL_542300, EPI_ISL_542301, EPI_ISL_542303, EPI_ISL_542304, EPI_ISL_542305, EPI_ISL_542306, EPI_ISL_542307, EPI_ISL_542308, EPI_ISL_542311, EPI_ISL_542312, EPI_ISL_542313, EPI_ISL_542314, EPI_ISL_542315, EPI_ISL_542316, EPI_ISL_542317, EPI_ISL_542318, EPI_ISL_542319, EPI_ISL_542321, EPI_ISL_542322, EPI_ISL_542323, EPI_ISL_542324, EPI_ISL_542325, EPI_ISL_542327, EPI_ISL_542328, EPI_ISL_542329, EPI_ISL_542330, EPI_ISL_542331, EPI_ISL_542333, EPI_ISL_542335, EPI_ISL_542336, EPI_ISL_542337, EPI_ISL_542338, EPI_ISL_542340, EPI_ISL_542341, EPI_ISL_542342, EPI_ISL_542343, EPI_ISL_542344, EPI_ISL_542345, EPI_ISL_542346, EPI_ISL_542347, EPI_ISL_542348, EPI_ISL_542352, EPI_ISL_542353, EPI_ISL_542354, EPI_ISL_542356, EPI_ISL_542357, EPI_ISL_542359, EPI_ISL_542362, EPI_ISL_542363, EPI_ISL_542364, EPI_ISL_542365, EPI_ISL_542372, EPI_ISL_542376, EPI_ISL_542378, EPI_ISL_542379, EPI_ISL_542380, EPI_ISL_542381, EPI_ISL_542382, EPI_ISL_542385, EPI_ISL_542387, EPI_ISL_542388, EPI_ISL_542389, EPI_ISL_542391, EPI_ISL_542394, EPI_ISL_542398 | | |
| Santa Casa de Misericórdia de Araçatuba | Instituto Adolfo Lutz, Interdiciplinary Procedures Center, Strategic Laboratory | Claudio Tavares Sacchi et al |
| EPI_ISL_547579 | | |
| Scripps Medical Laboratory | Andersen lab at Scripps Research | SEARCH Alliance San Diego with Michael Quigley et al |
| EPI_ISL_494599, EPI_ISL_494614 | | |
| SD Urban Indian Health Pierre | South Dakota Public Health Laboratory | Matt Plumb et al |
| EPI_ISL_569617 | | |

| | | |
|---|---|---|
| Seattle Flu Study | Seattle Flu Study | Chu et al et al |
| EPI_ISL_430128 | | |
| Seattle Flu Study | Seattle Flu Study | Deborah A. Nickerson et al |
| EPI_ISL_525709, EPI_ISL_525741, EPI_ISL_530134, EPI_ISL_530162 | | |
| Sentinelles network | National Reference Center for Viruses of Respiratory Infections, Institut Pasteur, Paris | Mélanie Albert et al |
| EPI_ISL_421514 | | |
| Service de Biologie clinique | National Reference Center for Viruses of Respiratory Infections, Institut Pasteur, Paris | Mélanie Albert et al |
| EPI_ISL_420042, EPI_ISL_421513 | | |
| Service de Biologie Médicale - BP 125 | National Reference Center for Viruses of Respiratory Infections, Institut Pasteur, Paris | Mélanie Albert et al |
| EPI_ISL_421501, EPI_ISL_420058, EPI_ISL_420060, EPI_ISL_420062, EPI_ISL_420064, EPI_ISL_421506, EPI_ISL_421512, EPI_ISL_428347 | | |
| Service de Virologie Hôpital Saint-Louis | Laboratory Cell Biology of Viral Infection-INSERM unit 944 | Laurent Meertens et al |
| EPI_ISL_469283 | | |
| Service de Virologie Hôpital Saint-Louis | Laboratory of Cell Biology of viral infection, Unit INSERM-U944 | Laurent Meertens et al |
| EPI_ISL_469282 | | |
| Servicio de Microbiología y Parasitología clínica. UCEIMP. Hospital Universitario Virgen del Rocío/IBIS/CSIC/US. | SeqCOVID-SPAIN consortium/IBV(CSIC) | Guillermo Martín Gutiérrez et al |
| EPI_ISL_452556 | | |
| Servicio de Microbiología, Hospital Universitario Son Espases | SeqCOVID-SPAIN consortium/IBV(CSIC) | Carla López-Causapé et al |
| EPI_ISL_469001, EPI_ISL_468969, EPI_ISL_541965 | | |

| | | |
|---|---|---|
| Servicio de Microbiología. Consorcio Hospital General Universitario de Valencia | Sequencing and Bioinformatics Service and Molecular Epidemiology Research Group. FISABIO-Public Health | Griselda De Marco et al |
| EPI_ISL_420114 | | |
| Servicio de Microbiologia. Hospital Clinico Universitario de Valencia | Sequencing and Bioinformatics Service and Molecular Epidemiology Research Group. FISABIO-Public Health | David Navarro et al |
| EPI_ISL_420130 | | |
| Servicio de Microbiologia. Hospital Clinico Universitario de Valencia | Sequencing and Bioinformatics Service and Molecular Epidemiology Research Group. FISABIO-Public Health | Inma Galán Vendrell et al |
| EPI_ISL_447531 | | |
| Servicio de Microbiologia. Hospital Clinico Universitario de Valencia | Sequencing and Bioinformatics Service and Molecular Epidemiology Research Group. FISABIO-Public Health | Marta Pla Diaz et al |
| EPI_ISL_436316, EPI_ISL_436322 | | |
| Servicio de Microbiologia. Hospital Clinico Universitario de Valencia | Sequencing and Bioinformatics Service and Molecular Epidemiology Research Group. FISABIO-Public Health | Vicente Soriano Chirona et al |
| EPI_ISL_436289 | | |
| Servicio de Microbiología. Hospital General Universitario de Castellón | SeqCOVID-SPAIN consortium/IBV(CSIC) | Rosario Moreno et al |
| EPI_ISL_537958, EPI_ISL_537960, EPI_ISL_538637, EPI_ISL_538651 | | |
| Servicio de Microbiologia. Hospital Universitario Doctor Peset | Sequencing and Bioinformatics Service and Molecular Epidemiology Research Group. FISABIO-Public Health | Juan Alberola Enguídanos et al |
| EPI_ISL_436251, EPI_ISL_436269 | | |
| Servicio de Microbiología. Hospital Universitario Donostia. OSI Donostialdea. Área de Enfermedades Infecciosas, Grupo de Infección Respiratoria y Resistencia Antimicrobiana. Instituto de Investigación Sanitaria Biodonostia | SeqCOVID-SPAIN consortium/Institute of Biomedicine of Valencia, IBV-CSIC | Gustavo Cilla et al |
| EPI_ISL_541103, EPI_ISL_541115, EPI_ISL_541119, EPI_ISL_541130, EPI_ISL_541131, EPI_ISL_541132, EPI_ISL_541133, EPI_ISL_541134, EPI_ISL_541135, EPI_ISL_541136, EPI_ISL_541137 | | |

| | | |
|---|---|---|
| Servicio de Microbiología. HRU de Málaga. Servicio Andaluz de Salud | SeqCOVID-SPAIN consortium/IBV(CSIC) | Inmaculada de Toro Peinado et al |
| EPI_ISL_452405, EPI_ISL_565911 | | |
| SERVIZIO DI IGIENE E SANITÀ PUBBLICA ASL Teramo | Istituto Zooprofilattico Sperimentale dell'Abruzzo e Molise "G.Caporale" | Lorusso A et al |
| EPI_ISL_435149, EPI_ISL_435154, EPI_ISL_435155, EPI_ISL_435153, EPI_ISL_436726, EPI_ISL_436727, EPI_ISL_436728, EPI_ISL_436729 | | |
| Servizio di igiene epidemiologia e sanità pubblica (SIESP)-Chieti | Istitituto Zooprofilattico Sperimentale dell'Abruzzo e Molise "G.Caporale" | Lorusso A et al |
| EPI_ISL_528994, EPI_ISL_529023, EPI_ISL_529024, EPI_ISL_528995, EPI_ISL_528996, EPI_ISL_528997, EPI_ISL_528998, EPI_ISL_528999, EPI_ISL_529000, EPI_ISL_529001, EPI_ISL_529002, EPI_ISL_529003, EPI_ISL_529025, EPI_ISL_529004, EPI_ISL_529005 | | |
| Servizio di Igiene, Epidemiologia e Sanità Pubblica (SIESP) Avezzano | Istituto Zooprofilattico Sperimentale dell'Abruzzo e Molise "G.Caporale" | Lorusso A et al |
| EPI_ISL_435152 | | |
| Servizio Igiene Epidemiologia e Sanità Pubblica (SIESP)-L'Aquila | Istitituto Zooprofilattico Sperimentale dell'Abruzzo e Molise "G.Caporale" | Lorusso A et al |
| EPI_ISL_529010, EPI_ISL_529011, EPI_ISL_529012 | | |
| Shanghai Public Health Clinical Center, Shanghai Medical College, Fudan University | National Research Center for Translational Medicine (Shanghai), Ruijin Hospital affiliated to Shanghai Jiao Tong University School of Medicine & Shanghai Public Health Clinical Center | Shengyue Wang et al |
| EPI_ISL_416347, EPI_ISL_416355, EPI_ISL_416405 | | |
| Skanes universitetssjukhus Lund | The Public Health Agency of Sweden | Anna-Malin Linde et al |
| EPI_ISL_560980 | | |
| Skovde/Unilabs | The Public Health Agency of Sweden | Anna-Malin Linde et al |
| EPI_ISL_455844, EPI_ISL_548246 | | |
| St.Olavs hospital/NTNU | Institute of Genomics Core Facility, University of Tartu | Aleksandr Ianevski et al |

| | | |
|---|---|---|
| EPI_ISL_450351, EPI_ISL_450349 | | |
| Stanford clinical virology lab | Chan-Zuckerberg Biohub | Benjamin Pinksy et al |
| EPI_ISL_450458, EPI_ISL_476778, EPI_ISL_476782, EPI_ISL_476790 | | |
| Suceava County Emergency Hospital | Stefan cel Mare University Metagenomics Lab | Lobiuc Andrei et al |
| EPI_ISL_491045 | | |
| Svardsjo VC | The Public Health Agency of Sweden | Tommy Janers et al |
| EPI_ISL_434645 | | |
| SVO Jundiaí | Instituto Adolfo Lutz, Interdiciplinary Procedures Center, Strategic Laboratory | Claudio Tavares Sacchi et al |
| EPI_ISL_547575 | | |
| Texas Department of State Health Services | Texas Department of State Health Services | Jenny Zhang et al |
| EPI_ISL_525771 | | |
| Texas Department of State Health Services | Texas Department of State Health Services | Rashmi Tuladhar et al |
| EPI_ISL_522965 | | |
| The First Affiliated Hospital of Guangzhou Medical University | BGI-shenzhen & The First Affiliated Hospital of Guangzhou Medical University | et al |
| EPI_ISL_429080 | | |
| The First Affiliated Hospital of Guangzhou Medical University | BGI-shenzhen & The First Affiliated Hospital of Guangzhou Medical University | Yanqun Wang et al |
| EPI_ISL_429089 | | |
| The National Institute of Public Health | State Veterinary Institute Prague | Nagy et al |
| EPI_ISL_541337, EPI_ISL_545957, EPI_ISL_546936, EPI_ISL_547966, EPI_ISL_547967, EPI_ISL_547968 | | |
| The National University Hospital of Iceland | deCODE genetics | Daniel F Gudbjartsson et al |

| | | |
|---|---|---|
| EPI_ISL_417727, EPI_ISL_417804, EPI_ISL_417837, EPI_ISL_417592, EPI_ISL_417622, EPI_ISL_424379, EPI_ISL_424442, EPI_ISL_424463, EPI_ISL_424471, EPI_ISL_417687, EPI_ISL_424572, EPI_ISL_424603 | | |
| TriCore Reference Laboratories | Center for Global Health, University of New Mexico Health Sciences Center | Daryl Domman et al |
| EPI_ISL_538237, EPI_ISL_542988 | | |
| TSGH-CP molecular lab | TSGH-CP molecular lab | Cherng-Lih Perng et al |
| EPI_ISL_436104 | | |
| TXDSHS | TXDSHS | Rashmi Tuladhar et al |
| EPI_ISL_560939 | | |
| UC San Diego Center for Advanced Laboratory Medicine | Andersen lab at Scripps Research | SEARCH Alliance San Diego with David Pride et al |
| EPI_ISL_483530, EPI_ISL_445116 | | |
| UCD National Virus Reference Laboratory | UCD National Virus Reference Laboratory | Michael Carr et al |
| EPI_ISL_418581, EPI_ISL_414587, EPI_ISL_418516, EPI_ISL_418584 | | |
| UCSF Clinical Microbiology Laboratory | Chan-Zuckerberg Biohub | CZB Cliahub Consortium et al |
| EPI_ISL_445177, EPI_ISL_445179, EPI_ISL_516734, EPI_ISL_516743, EPI_ISL_429050 | | |
| ULSS9 Distretto di Bussolengo | Istituto Zooprofilattico Sperimentale delle Venezie | Adelaide Milani et al |
| EPI_ISL_452181, EPI_ISL_522855, EPI_ISL_452184, EPI_ISL_422438, EPI_ISL_452185, EPI_ISL_452188, EPI_ISL_452189, EPI_ISL_522860, EPI_ISL_522861, EPI_ISL_522862, EPI_ISL_522863, EPI_ISL_522864, EPI_ISL_522865, EPI_ISL_522866, EPI_ISL_522867, EPI_ISL_522868 | | |

| | | |
|---|---|---|
| ULSS9 Distretto di San Bonifacio | Istituto Zooprofilattico Sperimentale delle Venezie | Adelaide Milani et al |
| EPI_ISL_452191, EPI_ISL_522856, EPI_ISL_452190, EPI_ISL_522858 | | |
| ULSS9 Scaligera | Istituto Zooprofilattico Sperimentale delle Venezie | Adelaide Milani et al |
| EPI_ISL_522857, EPI_ISL_522859 | | |
| UMMC-Health | WHO National Influenza Centre Russian Federation | Andrey Komissarov et al |
| EPI_ISL_560317 | | |
| Universitaetsklinik für Innere Medizin II Innsbruck | Bergthaler laboratory, CeMM Research Center for Molecular Medicine of the Austrian Academy of Sciences | Alexandra Popa et al |
| EPI_ISL_475765, EPI_ISL_437935, EPI_ISL_437950, EPI_ISL_437952, EPI_ISL_437964, EPI_ISL_437966, EPI_ISL_437969, EPI_ISL_475936 | | |
| University College London Hospital | COVID-19 Genomics UK (COG-UK) Consortium | Judith Heaney et al |
| EPI_ISL_507058, EPI_ISL_507064 | | |
| University College London, Great Ormond Street Hospital for Children NHS Foundation Trust, Imperial College Healthcare NHS Trust | COVID-19 Genomics UK (COG-UK) Consortium | Sergi Castellano et al |
| EPI_ISL_441043, EPI_ISL_449701, EPI_ISL_478431, EPI_ISL_478437 | | |
| University College London, Great Ormond Street Hospital for Children NHS Foundation Trust, Imperial College Healthcare NHS Trust | Wellcome Sanger Institute for the COVID-19 Genomics UK Consortium | Sergi Castellano et al |
| EPI_ISL_487606, EPI_ISL_492498 | | |
| University Hospital Basel, Clinical Virology | University Hospital Basel, Clinical Bacteriology | Hirsch et al |
| EPI_ISL_418275, EPI_ISL_418277, EPI_ISL_418278, EPI_ISL_418279, EPI_ISL_418280, EPI_ISL_418281, EPI_ISL_418282, EPI_ISL_418283, EPI_ISL_418284, EPI_ISL_418433, EPI_ISL_418434, EPI_ISL_418435, EPI_ISL_418439, EPI_ISL_418438, EPI_ISL_418440 | | |
| University Hospital Basel, Clinical Virology | University Hospital Basel, Clinical Bacteriology | Madlen Stange et al |

| |
|---|
| EPI_ISL_528030, EPI_ISL_528171, EPI_ISL_527917, EPI_ISL_527918, EPI_ISL_527919, EPI_ISL_527921, EPI_ISL_527926, EPI_ISL_527929, EPI_ISL_527936, EPI_ISL_527939, EPI_ISL_527940, EPI_ISL_527945, EPI_ISL_527946, EPI_ISL_527947, EPI_ISL_527948, EPI_ISL_527949, EPI_ISL_527950, EPI_ISL_527958, EPI_ISL_527960, EPI_ISL_527971, EPI_ISL_527972, EPI_ISL_527976, EPI_ISL_527984, EPI_ISL_527990, EPI_ISL_527993, EPI_ISL_527994, EPI_ISL_527999, EPI_ISL_528000, EPI_ISL_528002, EPI_ISL_528006, EPI_ISL_528011, EPI_ISL_528012, EPI_ISL_528013, EPI_ISL_528015, EPI_ISL_528023, EPI_ISL_528027, EPI_ISL_528032, EPI_ISL_528038, EPI_ISL_528040, EPI_ISL_528043, EPI_ISL_528045, EPI_ISL_528048, EPI_ISL_528049, EPI_ISL_528050, EPI_ISL_528051, EPI_ISL_528056, EPI_ISL_528062, EPI_ISL_528064, EPI_ISL_528065, EPI_ISL_528072, EPI_ISL_528073, EPI_ISL_528077, EPI_ISL_528079, EPI_ISL_528084, EPI_ISL_528086, EPI_ISL_528088, EPI_ISL_528090, EPI_ISL_528095, EPI_ISL_528099, EPI_ISL_528100, EPI_ISL_528103, EPI_ISL_528104, EPI_ISL_528106, EPI_ISL_528107, EPI_ISL_528108, EPI_ISL_528109, EPI_ISL_528110, EPI_ISL_528113, EPI_ISL_528117, EPI_ISL_528122, EPI_ISL_528125, EPI_ISL_528126, EPI_ISL_528130, EPI_ISL_528131, EPI_ISL_528133, EPI_ISL_528135, EPI_ISL_528137, EPI_ISL_528139, EPI_ISL_528144, EPI_ISL_528146, EPI_ISL_528147, EPI_ISL_528152, EPI_ISL_528154, EPI_ISL_528155, EPI_ISL_528157, EPI_ISL_528158, EPI_ISL_528161, EPI_ISL_528162, EPI_ISL_528169, EPI_ISL_528172, EPI_ISL_528175, EPI_ISL_528177, EPI_ISL_528179, EPI_ISL_528180, EPI_ISL_528184, EPI_ISL_528188, EPI_ISL_528192, EPI_ISL_528193, EPI_ISL_528197, EPI_ISL_528199, EPI_ISL_528201, EPI_ISL_528203, EPI_ISL_528210, EPI_ISL_528211, EPI_ISL_528212, EPI_ISL_528215, EPI_ISL_528219, EPI_ISL_528224, EPI_ISL_528225, EPI_ISL_528226, EPI_ISL_528231, EPI_ISL_528246, EPI_ISL_528265, EPI_ISL_528266, EPI_ISL_528267, EPI_ISL_528279, EPI_ISL_528289, EPI_ISL_528300, EPI_ISL_528301, EPI_ISL_528306, EPI_ISL_528308, EPI_ISL_528313, EPI_ISL_528321, EPI_ISL_528322, EPI_ISL_528328, EPI_ISL_528329, EPI_ISL_528352, EPI_ISL_528353, EPI_ISL_528368, EPI_ISL_528374, EPI_ISL_528379, EPI_ISL_528380, EPI_ISL_527916, EPI_ISL_527920, EPI_ISL_527922, EPI_ISL_527923, EPI_ISL_527924, EPI_ISL_527925, EPI_ISL_527927, EPI_ISL_527928, EPI_ISL_527930, EPI_ISL_527931, EPI_ISL_527932, EPI_ISL_527933, EPI_ISL_527934, EPI_ISL_527935, EPI_ISL_527938, EPI_ISL_527941, EPI_ISL_527942, EPI_ISL_527943, EPI_ISL_527944, EPI_ISL_527951, EPI_ISL_527952, EPI_ISL_527953, EPI_ISL_527954, EPI_ISL_527955, EPI_ISL_527956, EPI_ISL_527957, EPI_ISL_527959, EPI_ISL_527961, EPI_ISL_527962, EPI_ISL_527963, EPI_ISL_527964, EPI_ISL_527965, EPI_ISL_527966, EPI_ISL_527967, EPI_ISL_527968, EPI_ISL_527969, EPI_ISL_527970, EPI_ISL_527973, EPI_ISL_527974, EPI_ISL_527975, EPI_ISL_527977, EPI_ISL_527978, EPI_ISL_527979, EPI_ISL_527980, EPI_ISL_527981, EPI_ISL_527982, EPI_ISL_527983, EPI_ISL_527985, EPI_ISL_527986, EPI_ISL_527987, EPI_ISL_527988, EPI_ISL_527989, EPI_ISL_530122, EPI_ISL_527991, EPI_ISL_527992, EPI_ISL_527995, EPI_ISL_527996, EPI_ISL_527997, EPI_ISL_527998, EPI_ISL_528001, EPI_ISL_528003, EPI_ISL_528004, EPI_ISL_528005, EPI_ISL_528007, EPI_ISL_528008, EPI_ISL_528009, EPI_ISL_528010, EPI_ISL_528014, EPI_ISL_528016, EPI_ISL_528017, EPI_ISL_528018, EPI_ISL_528019, EPI_ISL_528020, EPI_ISL_528021, EPI_ISL_528022, EPI_ISL_528024, EPI_ISL_528025, EPI_ISL_528026, EPI_ISL_528028, EPI_ISL_528029, EPI_ISL_528031, EPI_ISL_528033, EPI_ISL_528034, EPI_ISL_528035, EPI_ISL_528036, EPI_ISL_528037, EPI_ISL_528039, EPI_ISL_528041, |

EPI_ISL_528042, EPI_ISL_528044, EPI_ISL_528046, EPI_ISL_528047, EPI_ISL_528052, EPI_ISL_528053, EPI_ISL_528054, EPI_ISL_528055, EPI_ISL_528057, EPI_ISL_528058, EPI_ISL_528059, EPI_ISL_528060, EPI_ISL_528061, EPI_ISL_528063, EPI_ISL_528066, EPI_ISL_528067, EPI_ISL_528068, EPI_ISL_528069, EPI_ISL_528070, EPI_ISL_528071, EPI_ISL_528074, EPI_ISL_528075, EPI_ISL_528076, EPI_ISL_528078, EPI_ISL_528080, EPI_ISL_528081, EPI_ISL_528082, EPI_ISL_528083, EPI_ISL_528085, EPI_ISL_528087, EPI_ISL_528089, EPI_ISL_528091, EPI_ISL_528092, EPI_ISL_528093, EPI_ISL_528094, EPI_ISL_528096, EPI_ISL_528097, EPI_ISL_528098, EPI_ISL_528101, EPI_ISL_528105, EPI_ISL_528111, EPI_ISL_528112, EPI_ISL_528114, EPI_ISL_528115, EPI_ISL_528116, EPI_ISL_528118, EPI_ISL_528119, EPI_ISL_528120, EPI_ISL_528121, EPI_ISL_528123, EPI_ISL_528124, EPI_ISL_528127, EPI_ISL_528128, EPI_ISL_528129, EPI_ISL_528132, EPI_ISL_528134, EPI_ISL_528136, EPI_ISL_528138, EPI_ISL_528141, EPI_ISL_528142, EPI_ISL_528143, EPI_ISL_528145, EPI_ISL_528148, EPI_ISL_528149, EPI_ISL_528150, EPI_ISL_528151, EPI_ISL_528153, EPI_ISL_528156, EPI_ISL_528160, EPI_ISL_528163, EPI_ISL_528164, EPI_ISL_528165, EPI_ISL_528166, EPI_ISL_528167, EPI_ISL_528168, EPI_ISL_528170, EPI_ISL_528174, EPI_ISL_528176, EPI_ISL_528178, EPI_ISL_528181, EPI_ISL_528182, EPI_ISL_528183, EPI_ISL_528185, EPI_ISL_528186, EPI_ISL_528187, EPI_ISL_528189, EPI_ISL_528190, EPI_ISL_528191, EPI_ISL_528194, EPI_ISL_528195, EPI_ISL_528196, EPI_ISL_528198, EPI_ISL_528200, EPI_ISL_528202, EPI_ISL_528204, EPI_ISL_528205, EPI_ISL_528206, EPI_ISL_528207, EPI_ISL_528209, EPI_ISL_528213, EPI_ISL_528214, EPI_ISL_528216, EPI_ISL_528217, EPI_ISL_528218, EPI_ISL_528220, EPI_ISL_528221, EPI_ISL_528222, EPI_ISL_528223, EPI_ISL_528227, EPI_ISL_528228, EPI_ISL_528229, EPI_ISL_528230, EPI_ISL_528232, EPI_ISL_528233, EPI_ISL_528234, EPI_ISL_528235, EPI_ISL_528236, EPI_ISL_528237, EPI_ISL_528238, EPI_ISL_528239, EPI_ISL_528240, EPI_ISL_528241, EPI_ISL_528242, EPI_ISL_528243, EPI_ISL_528244, EPI_ISL_528245, EPI_ISL_528247, EPI_ISL_528248, EPI_ISL_528249, EPI_ISL_528250, EPI_ISL_528251, EPI_ISL_528252, EPI_ISL_528253, EPI_ISL_528254, EPI_ISL_528255, EPI_ISL_528256, EPI_ISL_528257, EPI_ISL_528258, EPI_ISL_528259, EPI_ISL_528260, EPI_ISL_528261, EPI_ISL_528262, EPI_ISL_528263, EPI_ISL_528264, EPI_ISL_528268, EPI_ISL_528269, EPI_ISL_528270, EPI_ISL_528271, EPI_ISL_528272, EPI_ISL_528273, EPI_ISL_528274, EPI_ISL_528275, EPI_ISL_528276, EPI_ISL_528277, EPI_ISL_528278, EPI_ISL_528280, EPI_ISL_528281, EPI_ISL_528282, EPI_ISL_528283, EPI_ISL_528284, EPI_ISL_528285, EPI_ISL_528286, EPI_ISL_528287, EPI_ISL_528288, EPI_ISL_530123, EPI_ISL_528290, EPI_ISL_528291, EPI_ISL_528292, EPI_ISL_528293, EPI_ISL_528294, EPI_ISL_528295, EPI_ISL_528296, EPI_ISL_528297, EPI_ISL_528298, EPI_ISL_528299, EPI_ISL_528302, EPI_ISL_528303, EPI_ISL_528304, EPI_ISL_528305, EPI_ISL_528307, EPI_ISL_528309, EPI_ISL_528310, EPI_ISL_528311, EPI_ISL_528312, EPI_ISL_528314, EPI_ISL_528315, EPI_ISL_528316, EPI_ISL_528317, EPI_ISL_528318, EPI_ISL_528319, EPI_ISL_528320, EPI_ISL_528323, EPI_ISL_528324, EPI_ISL_528325, EPI_ISL_528326, EPI_ISL_528327, EPI_ISL_528330, EPI_ISL_528331, EPI_ISL_528332, EPI_ISL_528333, EPI_ISL_528334, EPI_ISL_528335, EPI_ISL_528336, EPI_ISL_528337, EPI_ISL_528338, EPI_ISL_528339, EPI_ISL_528340, EPI_ISL_528341, EPI_ISL_528342, EPI_ISL_528343, EPI_ISL_528344, EPI_ISL_528345, EPI_ISL_528346, EPI_ISL_528347, EPI_ISL_528348, EPI_ISL_528349, EPI_ISL_528350, EPI_ISL_528351, EPI_ISL_528354, EPI_ISL_528355,

| | | |
|---|---|---|
| EPI_ISL_528356, EPI_ISL_528357, EPI_ISL_528358, EPI_ISL_528359, EPI_ISL_528360, EPI_ISL_528361, EPI_ISL_528362, EPI_ISL_528363, EPI_ISL_528364, EPI_ISL_528365, EPI_ISL_528366, EPI_ISL_528367, EPI_ISL_528369, EPI_ISL_528370, EPI_ISL_528371, EPI_ISL_528372, EPI_ISL_528373, EPI_ISL_528375, EPI_ISL_528376, EPI_ISL_528377, EPI_ISL_528378, EPI_ISL_528381, EPI_ISL_527937, EPI_ISL_528102, EPI_ISL_528173, EPI_ISL_528208, EPI_ISL_528159, EPI_ISL_528140 | | |
| University Hospital Basel, Clinical Virology | University Hospital Basel, Clinical Virology | Hirsch et al |
| EPI_ISL_418436 | | |
| University Hospital Basel, Clinical Virology | University Hospital Basel, Labormedizin | Hirsch et al |
| EPI_ISL_418273 | | |
| University Hospital for Infectious Diseases "Dr. Fran Mihaljević", Research Unit | University of Zagreb, Centre for research and knowledge transfer in biotechnology | Ivan-Christian Kurolt et al |
| EPI_ISL_454581 | | |
| University Hospital of Northern Norway, Department for Microbiology and Infectious Disease Control | Norwegian Institute of Public Health, Department of Virology | Kathrine Stene-Johansen et al |
| EPI_ISL_420152 | | |
| University Hospital Zurich | Department of Biosystems Science and Engineering, ETH Zürich | Christian Beisel et al |
| EPI_ISL_483677, EPI_ISL_483682, EPI_ISL_483685, EPI_ISL_483668, EPI_ISL_483671, EPI_ISL_483674, EPI_ISL_483678, EPI_ISL_483680, EPI_ISL_483683, EPI_ISL_483669, EPI_ISL_483672, EPI_ISL_483675, EPI_ISL_483676, EPI_ISL_483679, EPI_ISL_483681, EPI_ISL_483684, EPI_ISL_483670, EPI_ISL_483673 | | |
| University Hospital Zürich | Institute of Medical Virology, University of Zurich | Stefan Schmutz et al |
| EPI_ISL_524782 | | |
| University Hospital Zürich | Institute of Medical Virology, University of Zurich | Verena Kufner et al |
| EPI_ISL_524482 | | |
| University Hospitals of Geneva Laboratory of Virology | University Hospitals of Geneva Laboratory of Virology | Laubscher F. et al |
| EPI_ISL_415457, EPI_ISL_415704, EPI_ISL_415456, EPI_ISL_415701, EPI_ISL_429212, EPI_ISL_429197, EPI_ISL_415455, EPI_ISL_429218, EPI_ISL_415706, EPI_ISL_429199, EPI_ISL_429207, EPI_ISL_415700, EPI_ISL_415454, EPI_ISL_429203, EPI_ISL_429208, EPI_ISL_429201, EPI_ISL_429219, EPI_ISL_429214, EPI_ISL_429196, EPI_ISL_429209, | | |

| | | |
|---|---|---|
| EPI_ISL_429222, EPI_ISL_415705, EPI_ISL_429223, EPI_ISL_429215, EPI_ISL_415708, EPI_ISL_429206, EPI_ISL_429211, EPI_ISL_429200, EPI_ISL_429220, EPI_ISL_429221, EPI_ISL_429217, EPI_ISL_415707, EPI_ISL_429204, EPI_ISL_429213, EPI_ISL_415458, EPI_ISL_429210, EPI_ISL_415459, EPI_ISL_415698, EPI_ISL_415699, EPI_ISL_415702, EPI_ISL_415703 | | |
| University of Birmingham | COVID-19 Genomics UK (COG-UK) Consortium | Institute of Microbiology et al |
| EPI_ISL_526314, EPI_ISL_526321, EPI_ISL_529605, EPI_ISL_529606 | | |
| University of California, Davis | Chan-Zuckerberg Biohub | CZB Cliahub Consortium et al |
| EPI_ISL_548477, EPI_ISL_548374 | | |
| University of Debrecen, Department of Medical Microbiology | National Laboratory of Virology, Szentágothai Research Centre | Endre Gábor Tóth et al |
| EPI_ISL_476074, EPI_ISL_476075, EPI_ISL_476077 | | |
| University of Exeter | COVID-19 Genomics UK (COG-UK) Consortium | Ben Temperton et al |
| EPI_ISL_457081, EPI_ISL_457177, EPI_ISL_457200 | | |
| University of Liège COVID-19 testing center | GIGA Medical Genomics | Keith Durkin et al |
| EPI_ISL_540443, EPI_ISL_540444, EPI_ISL_540445, EPI_ISL_540446, EPI_ISL_540447, EPI_ISL_540455, EPI_ISL_540457, EPI_ISL_540462, EPI_ISL_540463, EPI_ISL_540464, EPI_ISL_540466, EPI_ISL_540467 | | |
| University of Miami Immunology and Histocompatibility Laboratory | University of Miami Immunology and Histocompatibility Laboratory | Emilio Margolles-Clark et al |
| EPI_ISL_513290, EPI_ISL_527385, EPI_ISL_527396 | | |
| University of Szeged, Institute of Clinical Microbiology | National Laboratory of Virology, Szentágothai Research Centre | Endre Gábor Tóth et al |
| EPI_ISL_477620, EPI_ISL_477622, EPI_ISL_476078 | | |

| | | |
|---|---|---|
| University of Washington Virology Lab | University of Washington Virology Lab | Pavitra Roychoudhury et al |
| EPI_ISL_501090, EPI_ISL_501129 | | |
| University of Wisconsin-Madison AIDS Vaccine Research Laboratories | University of Wisconsin-Madison AIDS Vaccine Research Laboratories | Gage Moreno et al |
| EPI_ISL_516501, EPI_ISL_480365, EPI_ISL_516448, EPI_ISL_516451, EPI_ISL_516460, EPI_ISL_516478, EPI_ISL_516484, EPI_ISL_516496, EPI_ISL_425166, EPI_ISL_522907, EPI_ISL_522930, EPI_ISL_536587, EPI_ISL_536625, EPI_ISL_536626, EPI_ISL_547581, EPI_ISL_536640, EPI_ISL_428261, EPI_ISL_428304, EPI_ISL_491374, EPI_ISL_495484, EPI_ISL_495504, EPI_ISL_421341, EPI_ISL_509858, EPI_ISL_509898, EPI_ISL_509905, EPI_ISL_509912 | | |
| unknown | Department of Medicine | Kassela et al |
| EPI_ISL_447647 | | |
| unknown | Department of Microbiology | Gao et al |
| EPI_ISL_455693 | | |
| unknown | Faculty of Medicine | Rodpan et al |
| EPI_ISL_437623, EPI_ISL_437621 | | |
| unknown | Instituto Nacional de Saude (INSA) | Borges et al et al |
| EPI_ISL_453915, EPI_ISL_453944, EPI_ISL_453970, EPI_ISL_454019, EPI_ISL_454033, EPI_ISL_454039, EPI_ISL_454300, EPI_ISL_454323 | | |
| unknown | National Reference Center for Viruses of Respiratory Infections, Institut Pasteur, Paris | Mélnie Albert et al |
| EPI_ISL_414626 | | |
| Utah Public Health Laboratory | Utah Public Health Laboratory | Erin L. Young et al |
| EPI_ISL_524274 | | |
| Utah Public Health Laboratory | Utah Public Health Laboratory | Erin Young et al |
| EPI_ISL_430037, EPI_ISL_560892 | | |
| UW Virology Lab | UW Virology Lab | Pavitra Roychoudhury et al |

| | | |
|---|---|---|
| EPI_ISL_477677, EPI_ISL_430945, EPI_ISL_476912, EPI_ISL_570614, EPI_ISL_427173, EPI_ISL_423031, EPI_ISL_477689, EPI_ISL_424236, EPI_ISL_424245, EPI_ISL_424310, EPI_ISL_570845, EPI_ISL_570990, EPI_ISL_570991, EPI_ISL_570993, EPI_ISL_570827, EPI_ISL_571013, EPI_ISL_570562, EPI_ISL_570630, EPI_ISL_570634, EPI_ISL_570637, EPI_ISL_570638, EPI_ISL_570642, EPI_ISL_570647, EPI_ISL_570667, EPI_ISL_570670, EPI_ISL_570748, EPI_ISL_570769, EPI_ISL_570771, EPI_ISL_491018, EPI_ISL_429652, EPI_ISL_430873, EPI_ISL_416719, EPI_ISL_418032, EPI_ISL_418033 | | |
| VI-US Virgin Islands Department of Health | Pathogen Discovery, Respiratory Viruses Branch, Division of Viral Diseases, Centers for Disease Control and Prevention | Yan Li et al |
| EPI_ISL_450804 | | |
| Victorian Infectious Diseases Reference Laboratory (VIDRL) | Microbiological Diagnostic Unit Public Health Laboratory and Victorian Infectious Diseases Reference Laboratory, Doherty Institute | Caly L. et al |
| EPI_ISL_426650, EPI_ISL_426745, EPI_ISL_426806, EPI_ISL_426821 | | |
| Victorian Infectious Diseases Reference Laboratory (VIDRL) | Microbiological Diagnostic Unit Public Health Laboratory and Victorian Infectious Diseases Reference Laboratory, The Peter Doherty Institute for Infection and Immunity | Caly L. et al |
| EPI_ISL_430473, EPI_ISL_430493 | | |
| Victorian Infectious Diseases Reference Laboratory (VIDRL) | Victorian Infectious Diseases Reference Laboratory and Microbiological Diagnostic Unit Public Health Laboratory, Doherty Institute | Caly L. et al |
| EPI_ISL_419905, EPI_ISL_419949, EPI_ISL_419950, EPI_ISL_419974, EPI_ISL_419748, EPI_ISL_419809 | | |
| Vigilância em Saúde de Cajamar | Instituto Adolfo Lutz, Interdiciplinary Procedures Center, Strategic Laboratory | Claudio Tavares Sacchi et al |
| EPI_ISL_547573 | | |
| Villa Serena del Dr. Leonardo Petruzzi | Istituto Zooprofilattico Sperimentale dell'Abruzzo e Molise "G.Caporale" | Lorusso A et al |
| EPI_ISL_435146, EPI_ISL_435147 | | |
| Viral Respiratory Lab, National Institute for Biomedical Research (INRB) | Pathogen Sequencing Lab, National Institute for Biomedical Research (INRB) | Placide Mbala-Kingebeni et al |

| | | |
|---|---|---|
| EPI_ISL_420032, EPI_ISL_417434, EPI_ISL_417437 | | |
| Virginia DCLS | Virginia DCLS | Virginia DCLS et al |
| EPI_ISL_467943, EPI_ISL_485849, EPI_ISL_522823, EPI_ISL_528640, EPI_ISL_528660, EPI_ISL_529937, EPI_ISL_529946, EPI_ISL_529954, EPI_ISL_572300, EPI_ISL_572253, EPI_ISL_572221 | | |
| Virology Department, Royal Infirmary of Edinburgh, NHS Lothian | Virology Department, Royal Infirmary of Edinburgh, NHS Lothian | McHugh M et al |
| EPI_ISL_415640 | | |
| Virology Department, Royal Infirmary of Edinburgh, NHS Lothian / School of Biological Sciences, University of Edinburgh | Wellcome Sanger Institute for the COVID-19 Genomics UK Consortium | McHugh M et al |
| EPI_ISL_491569, EPI_ISL_487684, EPI_ISL_488931, EPI_ISL_488917, EPI_ISL_487876, EPI_ISL_487908 | | |
| Virology Department, Royal Infirmary of Edinburgh, NHS Lothian / School of Biological Sciences, University of Edinburgh / Institute of Genetics and Molecular Medicine, University of Edinburgh | COVID-19 Genomics UK (COG-UK) Consortium | McHugh M et al |
| EPI_ISL_425819, EPI_ISL_425825, EPI_ISL_425833, EPI_ISL_425839, EPI_ISL_425844, EPI_ISL_425856, EPI_ISL_425881, EPI_ISL_439144, EPI_ISL_439145, EPI_ISL_433070, EPI_ISL_433192, EPI_ISL_439213, EPI_ISL_425971, EPI_ISL_439337, EPI_ISL_425993, EPI_ISL_453106, EPI_ISL_449319, EPI_ISL_449329, EPI_ISL_439671, EPI_ISL_432868, EPI_ISL_432873 | | |
| Virology Department, Sheffield Teaching Hospitals NHS Foundation Trust | Department of Infection, Immunity and Cardiovascular Disease, The Florey Institute, The Medical School, University of Sheffield | Thushan de Silva et al |
| EPI_ISL_416740, EPI_ISL_420238, EPI_ISL_414500 | | |
| Virology Department, Sheffield Teaching Hospitals NHS Foundation Trust/Department of Infection, Immunity and Cardiovascular Disease, The Medical School, University of Sheffield | COVID-19 Genomics UK (COG-UK) Consortium | Thushan de Silva et al |
| EPI_ISL_475379, EPI_ISL_441984, EPI_ISL_453752 | | |

| | | |
|---|---|---|
| Wadsworth Center, New York State Department.of Health | Wadsworth Center, New York State Department.of Health | Kirsten St. George et al |
| EPI_ISL_426045 | | |
| Wales Specialist Virology Centre | Public Health Wales Microbiology Cardiff | Catherine Moore et al |
| EPI_ISL_419438, EPI_ISL_419497, EPI_ISL_432188, EPI_ISL_431966, EPI_ISL_432302, EPI_ISL_432086, EPI_ISL_432196, EPI_ISL_445822, EPI_ISL_446692 | | |
| Wales Specialist Virology Centre Sequencing lab: Pathogen Genomics Unit | COVID-19 Genomics UK (COG-UK) Consortium | Catherine Moore et al |
| EPI_ISL_472442, EPI_ISL_472816, EPI_ISL_479417 | | |
| Washington State Department of Health | Seattle Flu Study | Chu et al et al |
| EPI_ISL_463625, EPI_ISL_430179, EPI_ISL_430294, EPI_ISL_449885, EPI_ISL_449948 | | |
| Washington State Department of Health | Seattle Flu Study | Deborah A. Nickerson et al |
| EPI_ISL_495722, EPI_ISL_495994, EPI_ISL_496038, EPI_ISL_496174, EPI_ISL_497353, EPI_ISL_497471, EPI_ISL_497304 | | |
| West China Hospital of Sichuan University | State Key Laboratory of Biotherapy of Sichuan University | Baowen Du et al |
| EPI_ISL_451345 | | |
| West of Scotland Specialist Virology Centre, NHSGGC / MRC-University of Glasgow Centre for Virus Research | COVID-19 Genomics UK (COG-UK) Consortium | Ana da Silva Filipe et al |
| EPI_ISL_425647, EPI_ISL_438762, EPI_ISL_433557, EPI_ISL_425690, EPI_ISL_433658, EPI_ISL_473562, EPI_ISL_439057, EPI_ISL_529686, EPI_ISL_540781, EPI_ISL_535108, EPI_ISL_535139, EPI_ISL_425779, EPI_ISL_425790, EPI_ISL_425799, EPI_ISL_425803, EPI_ISL_425805 | | |
| WHO National Influenza Centre Russian Federation | WHO National Influenza Centre Russian Federation | Andrey Komissarov et al |
| EPI_ISL_507287 | | |

| | | |
|---|---|---|
| Wisconsin State Laboratory of Hygiene Communicable Disease Division | Wisconsin State Laboratory of Hygiene Communicable Disease Division | Kelsey R. Florek et al |
| EPI_ISL_471180 | | |
| Wuhan Chain Medical Labs (CMLabs) | State Key Laboratory of Biotherapy of Sichuan University | Baowen Du et al |
| EPI_ISL_454928, EPI_ISL_454956 | | |
| Wyoming Public Health Laboratory | Center for Global Health, University of New Mexico Health Sciences Center | Daryl Domman et al |
| EPI_ISL_462970 | | |
| Zentralinstitut für medizinische und chemische Labordiagnostik, Universitätskliniken Insbruck | Bergthaler laboratory, CeMM Research Center for Molecular Medicine of the Austrian Academy of Sciences | Alexandra Popa et al |
| EPI_ISL_475887, EPI_ISL_475899, EPI_ISL_475904, EPI_ISL_475906, EPI_ISL_475908 | | |
| ZOTZ KLIMAS MVZ Düsseldorf-Centrum GbR ÜBAG für Labormedizin, Genetik, Zytologie, Pathologie | Center of Medical Microbiology, Virology, and Hospital Hygiene, University of Duesseldorf | Maximilian Damagnez et al |
| EPI_ISL_539577, EPI_ISL_539578, EPI_ISL_539579, EPI_ISL_539580, EPI_ISL_539581, EPI_ISL_539582, EPI_ISL_539583, EPI_ISL_539584, EPI_ISL_539585, EPI_ISL_539586, EPI_ISL_539587, EPI_ISL_539589, EPI_ISL_539590, EPI_ISL_539591, EPI_ISL_539592, EPI_ISL_539593, EPI_ISL_539594, EPI_ISL_539595, EPI_ISL_539596, EPI_ISL_539597, EPI_ISL_539598, EPI_ISL_539599, EPI_ISL_539600, EPI_ISL_539601, EPI_ISL_539602, EPI_ISL_539603, EPI_ISL_539604, EPI_ISL_539605, EPI_ISL_539606, EPI_ISL_539607, EPI_ISL_539608, EPI_ISL_539609, EPI_ISL_539610, EPI_ISL_539611, EPI_ISL_539612, EPI_ISL_539614, EPI_ISL_539615 | | |
